# Immune Correlates Analysis of the PREVENT-19 COVID-19 Vaccine Efficacy Clinical Trial

**DOI:** 10.1101/2022.06.22.22276362

**Authors:** Youyi Fong, Yunda Huang, David Benkeser, Lindsay N. Carpp, Germán Áñez, Wayne Woo, Alice McGarry, Lisa M. Dunkle, Iksung Cho, Christopher R. Houchens, Karen Martins, Lakshmi Jayashankar, Flora Castellino, Christos J. Petropoulos, Andrew Leith, Deanne Haugaard, Bill Webb, Yiwen Lu, Chenchen Yu, Bhavesh Borate, Lars W. P. van der Laan, Nima S. Hejazi, April K. Randhawa, Michele P. Andrasik, James G. Kublin, Julia Hutter, Maryam Keshtkar-Jahromi, Tatiana H. Beresnev, Lawrence Corey, Kathleen M. Neuzil, Dean Follmann, Julie A. Ake, Cynthia L. Gay, Karen L. Kotloff, Richard A. Koup, Ruben O. Donis, Peter B. Gilbert, the Coronavirus Vaccine Prevention Network (CoVPN)/2019nCoV-301 Principal Investigators and Study Team, the United States Government (USG)/CoVPN Biostatistics Team

**Affiliations:** Vaccine and Infectious Disease Division, Fred Hutchinson Cancer Center, Seattle, WA, USA.; Public Health Sciences Division, Fred Hutchinson Cancer Center, Seattle, WA, USA.; Department of Global Health, University of Washington; Seattle, WA 98195, USA.; Department of Biostatistics and Bioinformatics, Rollins School of Public Health, Emory University, Atlanta, GA, USA.; Novavax, Inc., Gaithersburg, MD, USA.; Biomedical Advanced Research and Development Authority, Washington, DC, USA.; LabCorp-Monogram Biosciences, South San Francisco, CA, USA.; Nexelis, Seattle, WA, USA.; Department of Statistics, University of Washington, Seattle, WA, USA.; Department of Biostatistics, T.H. Chan School of Public Health, Harvard University, Boston, MA, USA.; Division of Microbiology and Infectious Diseases, National Institute of Allergy and Infectious Diseases, NIH, Rockville, MD, USA; Department of Laboratory Medicine and Pathology, University of Washington, Seattle, WA, USA.; Center for Vaccine Development and Global Health, University of Maryland School of Medicine, Baltimore, MD, USA.; Biostatistics Research Branch, National Institute of Allergy and Infectious Diseases, National Institutes of Health, Bethesda, MD, USA.; U.S. Military HIV Research Program, Walter Reed Army Institute of Research, Silver Spring, MD, USA.; University of North Carolina School of Medicine, Chapel Hill, NC, USA.; Vaccine Research Center, National Institute of Allergy and Infectious Diseases, National Institutes of Health, Bethesda, MD, USA.; Department of Biostatistics, School of Public Health, University of Washington, Seattle, WA, USA.

## Abstract

In the randomized, placebo-controlled PREVENT-19 phase 3 trial conducted in the U.S. and Mexico of the NVX-CoV2373 adjuvanted, recombinant spike protein nanoparticle vaccine, anti-spike binding IgG concentration (spike IgG) and pseudovirus 50% neutralizing antibody titer (nAb ID50) measured two weeks after two doses were assessed as correlates of risk and as correlates of protection against PCR-confirmed symptomatic SARS-CoV-2 infection (COVID- 19). These immune correlates analyses were conducted in the U.S. cohort of baseline SARS-CoV-2 negative per-protocol participants using a case-cohort design that measured the antibody markers from all 12 vaccine recipient breakthrough COVID-19 cases starting 7 days post antibody measurement and from 639 vaccine recipient non-cases (Mexico was excluded due to zero breakthrough cases with the efficacy data cut-off date April 19, 2021). In vaccine recipients, the baseline risk factor-adjusted hazard ratio of COVID-19 was 0.36 (95% CI: 0.20, 0.63), p<0.001 (adjusted p-0.005) per 10-fold increase in IgG spike concentration and 0.39 (0.19, 0.82), p=0.013 (adjusted p=0.030) per 10-fold increase in nAb ID50 titer. At spike IgG concentration 100, 1000, and 6934 binding antibody units/ml (100 is the 3^rd^ percentile, 6934 is the 97.5^th^ percentile), vaccine efficacy to reduce the probability of acquiring COVID-19 at 59 days post marker measurement was 65.5% (95% CI: 23.0%, 90.8%), 87.7% (77.7%, 94.4%), and 94.8% (88.0%, 97.9%), respectively. At nAb ID50 titers of 50, 100, 1000, and 7230 IU50/ml (50 is the 5^th^ percentile, 7230 the 97.5^th^ percentile), these estimates were 75.7% (49.8%, 93.2%), 81.7% (66.3%, 93.2%), 92.8% (85.1%, 97.4%) and 96.8% (88.3%, 99.3%). The same two antibody markers were assessed as immune correlates via the same study design and statistical analysis in the mRNA-1273 phase 3 COVE trial (except in COVE the markers were measured four weeks post dose two). Spike IgG levels were slightly lower and nAb ID50 titers slightly higher after NVX-CoV2373 than after mRNA-1273 vaccination. The strength of the nAb ID50 correlate was similar between the trials, whereas the spike IgG antibodies appeared to correlate more strongly with NVX-CoV2373 in PREVENT-19, as quantified by the hazard ratio and the degree of change in vaccine efficacy across antibody levels. However, the relatively few breakthrough cases in PREVENT-19 limited the ability to infer a stronger correlate. The conclusion is that both markers were consistent correlates of protection for the two vaccines, supporting potential cross-vaccine platform applications of these markers for guiding decisions about vaccine approval and use.

## Introduction

The NVX-CoV2373 (Novavax) SARS-CoV-2 vaccine is composed of full-length, stabilized, prefusion, recombinant spike protein trimers produced from the Wuhan-Hu-1 sequence, assembled into nanoparticles coformulated with a saponin-based adjuvant (Matrix-M^TM^). NVX- CoV2373 was found to be safe and immunogenic in adults^1, 2^ and to provide high vaccine efficacy against COVID-19 of any severity caused by the B.1.1.7 (alpha) variant in a phase 3 randomized, placebo-controlled trial in the United Kingdom.^4^ Furthermore, the PREVENT-19 (Prefusion Protein Subunit Vaccine Efficacy Novavax Trial–COVID-19) (NCT04611802) phase 3 randomized, placebo-controlled trial of NVX-CoV2373 in adults 18 years of age or older in the United States (US) and Mexico conducted during a period in which the circulating variants were predominantly B.1.1.7 (alpha), B.1.351 (beta), P.1 (gamma), B.1.427/B.1.429 (epsilon), and B.1.526 (iota)^5, 6^ showed adequate safety and high vaccine efficacy against PCR-confirmed symptomatic COVID-19.^7^

PREVENT-19 randomized 29,949 participants in a 2:1 ratio to receive 2 doses of NVX-CoV2373 or placebo between December 27, 2020, and February 18, 2021. Based on occurrence of 77 COVID-19 primary endpoints over 3 months of follow-up post first vaccination (14 among vaccine recipients and 63 among placebo recipients at least 7 days after the second vaccine dose), vaccine efficacy was 90.4%; 95% confidence interval [CI], 82.9 to 94.6; P<0.001. As reported in Dunkle et al.^6^, of 61 COVID-19 primary endpoints with SARS-CoV-2 genomes sequenced from nasal swabs, 35 were classified as variants of concern and 13 were classified as variants of interest, based on the Centers for Disease Control and Prevention classification in June of 2021, and vaccine efficacy against any variant of concern or interest was 92.6% (95% CI, 83.6 to 96.7). These variants (different from Wuhan-Hu-1) are no longer considered to be variants of concern or of interest. The predominant variant was B.1.1.7 (alpha) (31 of 61 genomes) with small COVID-19 endpoint counts from more than a dozen other variants (see “*SARS-CoV-2 lineages causing COVID-19 endpoints”* for additional information). The NVX- CoV2373 vaccine has been issued an Emergency Use Listing by the World Health Organization,^7^ conditional authorization by the European Commission,^8^ and approval or authorization in nearly 40 countries.^9^

Validation of an immune biomarker as a correlate of protection (CoP)^11–13^ against COVID-19 would aid decisions for approval and use of COVID-19 vaccines, for example by allowing approval for populations not represented in phase 3 trial cohorts (e.g. pediatrics) or facilitating approval of alternative formulations or schedules Two specific interrelated applications are estimation of the durability of vaccine protection and estimation of the breadth of vaccine protection against a set of circulating and emerging strains. Moreover, a validated immune biomarker guides research to develop next-generation vaccines by providing an immunogenicity study endpoint for ranking and down-selection of candidate vaccine regimens.

For many licensed vaccines against viral diseases, either binding antibodies (bAbs) or neutralizing antibodies (nAbs) have been validated as CoPs for certain applications,^11^ and the body of evidence supporting these markers as CoPs for COVID-19 vaccines has been steadily growing.^14–22^ The US Government (USG) COVID-19 Response Team and the vaccine developers partnered to design and implement five harmonized phase 3 COVID-19 vaccine efficacy trials, with one of the major objectives to develop a CoP based on an IgG bAb or nAb assay.^22^ The first correlates analysis in this program evaluated the mRNA-1273 COVID-19 vaccine in the COVE phase 3 trial,^24^ and showed that both IgG bAb and nAb markers measured four weeks post first dose and post second dose were associated with vaccine efficacy against symptomatic COVID-19, with nAb titer mediating about two-thirds of the vaccine efficacy.^25^

These markers were IgG bAbs against SARS-CoV-2 spike protein (“spike IgG”), IgG bAbs against the spike protein receptor binding domain (“RBD IgG”), and neutralizing antibodies measured by a pseudovirus neutralization assay (50% inhibitory dilution, “nAb ID50”), with all results reported in the World Health Organization (WHO) International Units. The second correlates analysis in this program evaluated the Ad26.COV2.S COVID-19 vaccine in the international ENSEMBLE phase 3 trial, studying the same antibody markers as correlates, and showed that nAb ID50 measured four weeks after a single vaccine dose was associated with single-dose vaccine efficacy against symptomatic COVID-19, and spike IgG and RBD IgG showed non-significant trends toward associating with vaccine efficacy against symptomatic COVID-19.^26^ External to the USG-supported program, the phase 3 trial of the AZD12222 (ChAdOx1 nCoV-19) vaccine in the United Kingdom showed that the same three markers, measured 4 weeks post second vaccination, all associated with vaccine efficacy against symptomatic COVID-19.^27^ In the current article, we present the correlates analysis for the PREVENT-19 study. The same spike IgG and nAb ID50 markers are assessed as correlates (the RBD IgG marker cannot yet be assessed because the assay is still undergoing validation). The study design and statistical methods for correlates is very similar to that applied to the COVE and ENSEMBLE studies, although because the number of vaccine breakthrough cases evaluable for correlates was small in PREVENT-19 (12, compared to 36 in COVE and 92 in ENSEMBLE), it was only possible to apply a subset of the correlates statistical methods specified in the harmonized Statistical Analysis Plan (SAP).^27^ Because all 12 breakthrough cases occurred in the US, the entire correlates analysis is restricted to the US participants.

## Results

### Immunogenicity subcohort and case-cohort set

The assessment of immune correlates was based on measurement of spike IgG and nAb ID50 at D35 (hereafter, “D35” denotes the Day 35 study visit, with an allowable visit window of + 7 days post Day 35) in the case-cohort set, comprised of a stratified random sample of the study cohort (the “immunogenicity subcohort”) plus all vaccine recipients experiencing the COVID-19 primary endpoint after D35 (“breakthrough cases”) (Supplementary Figure 1A). Detailed information on the sampling design is in the SAP, provided as Supplementary Material. D35 spike IgG antibody data were available from 12 vaccine recipient breakthrough cases and 639 non-cases. D35 nAb ID50 data were available from 11 of the 12 vaccine recipient breakthrough cases and from a subset (628) of the 639 non-cases; the missing nAb ID50 values from vaccine cases and non-cases were imputed from their spike IgG values using predictive mean matching (default for imputing a quantitative variable with the R package *mice*). All analyses of D35 antibody markers restricted to baseline SARS-CoV-2 negative per-protocol participants in the case-cohort set (Supplementary Table 1, Supplementary Figure 2).

### Participant demographics

Participant demographics and clinical characteristics of participants selected for the U.S. immunogenicity subcohort (N=669 in the vaccine group, N=76 in the placebo group) are shown in Supplementary Table 2. The demographics description includes a larger number of vaccine recipients in the immunogenicity subcohort (669 vs. 639) because 30 participants did not qualify for the immunogenicity subcohort due to not having D0 and D35 antibody data or due to evidence of SARS-CoV-2 infection by 6 days post D35. Of all participants selected, 46.7% were ≥ 65 years old, 49.7% had co-existing conditions associated with high risk of severe COVID-19 (as listed in Dunkle et al.^7^, these were obesity, chronic lung disease, diabetes mellitus type 2, cardiovascular disease, and/or chronic kidney disease), and 46.7% were female. Forty-two and a half percent had minority status (defined as other than White Non-Hispanic), with 19.6% of the participants being Black or African American, 20.8% Hispanic or Latino, and 2.6% American Indian or Alaska Native. Of note, the balanced distribution of these participants according to these key factors is not reflective of the study cohort but of the sampling design of the immunogenicity cohort; the sampling design was accounted for in the statistical inferences of the antibody data.

### COVID-19 study endpoint

Correlates analyses were performed based on adjudicated COVID-19 primary endpoints, the same COVID-19 endpoint definition studied in Dunkle et al.^7^ However, endpoint onset was required to be at least 7 days post-D35 visit, differing from Dunkle et al.^7^ where onset was required to be at least 7 days post-D21 (second dose). The correlates analyses excluded COVID-19 endpoints between 1 and 6 days post-D35 visit because some of these endpoints likely had SARS-CoV-2 infection before D35, which could possibly generate anamnestic responses that would impact D35 antibody levels. In both the correlates analyses and Dunkle et al. COVID-19 endpoints were included through to April 19, 2021, the data cut date of the primary analysis. Vaccine recipient non-cases were defined as baseline SARS-CoV-2 negative per-protocol participants sampled into the immunogenicity subcohort with D0 and D35 antibody data measured with no evidence of SARS-CoV-2 infection (i.e., never tested RT-PCR positive) up to the end of the correlates study period (April 19, 2021). Correlates analyses based on the cumulative probability of COVID-19 through a fixed time point post D35 selected 59 days as the time point for analysis, where 59 days was selected as the latest day post-D35 among the 12 vaccine breakthrough COVID-19 endpoint cases with antibody data, which was 73 days post dose two.

### SARS-CoV-2 lineages causing COVID-19 endpoints

Dunkle et al.^7^ reported the variants of concern and variants of interest of the COVID-19 primary endpoints in PREVENT-19. Of the s77 primary COVID-19 endpoints, lineage information was available from 61 endpoints, and of the 56 COVID-19 endpoints at least 7 days post-D35 visit and hence included in the immune correlates analyses, lineage information was available from 44 endpoints. Supplementary Table 3 shows the distribution of SARS-CoV-2 variants that caused these 44 COVID-19 endpoints by randomization arm. Eleven of these variants were of the Wuhan Ancestral lineage (genetically close to the vaccine strain), and 27 and 6 of these variants were classified by the Center for Disease Control and Prevention (June 2021) as variants of concern and as variants of interest, respectively. Approximately 50% of the 44 sequences were alpha (B.1.1.7), with 13 other variants each causing between 1 and 4 COVID- 19 endpoints. All 44 sequences pre-dated the emergence of B.1.617/AY delta and BA.1-5 omicron variants of concern.

### Vaccine recipient non-cases had higher D35 spike IgG concentrations and neutralization ID50 titers than vaccine breakthrough cases

At D35, 99.6% (95% CI: 99.2%, 99.8%) of vaccine recipient non-cases had a positive spike IgG response [defined by IgG > 10.8424 binding antibody units (BAU)/ml] and 98.8% (95% CI: 97.5%, 99.4%) had a detectable nAb ID50 titer (defined by nAb ID50 > 2.612 IU50/ml, the assay detection limit) (Figure 1, Table 1). For both D35 markers, the frequency of vaccine recipients with positive/detectable response was lower in cases than in non-cases: For Spike IgG, 91.7% (52.5%, 99.1%) of cases had a positive response, with a difference in frequencies for non-cases minus cases of 8.0% (0.5%, 47.2%). For nAb ID50, 83.4% (47.6%, 96.5%) of cases had a detectable nAb ID50 response, with a difference in frequencies for non-cases minus cases of 15.4% (2.2%, 51.2%)] (Table 1). For both D35 markers, the geometric mean value was about 3 times higher for non-cases than for cases [spike IgG geometric mean 1552 BAU/ml (1407, 1713) in non-cases vs. 528 BAU/ml (184, 1513) in cases, ratio = 2.9 (1.0, 8.3); nAb ID50 geometric mean 461 IU50/ml (404, 526) in non-cases vs. 135 IU50/ml (35, 519) in cases, ratio = 3.4 (0.9, 12.5)].

**Figure 1.**
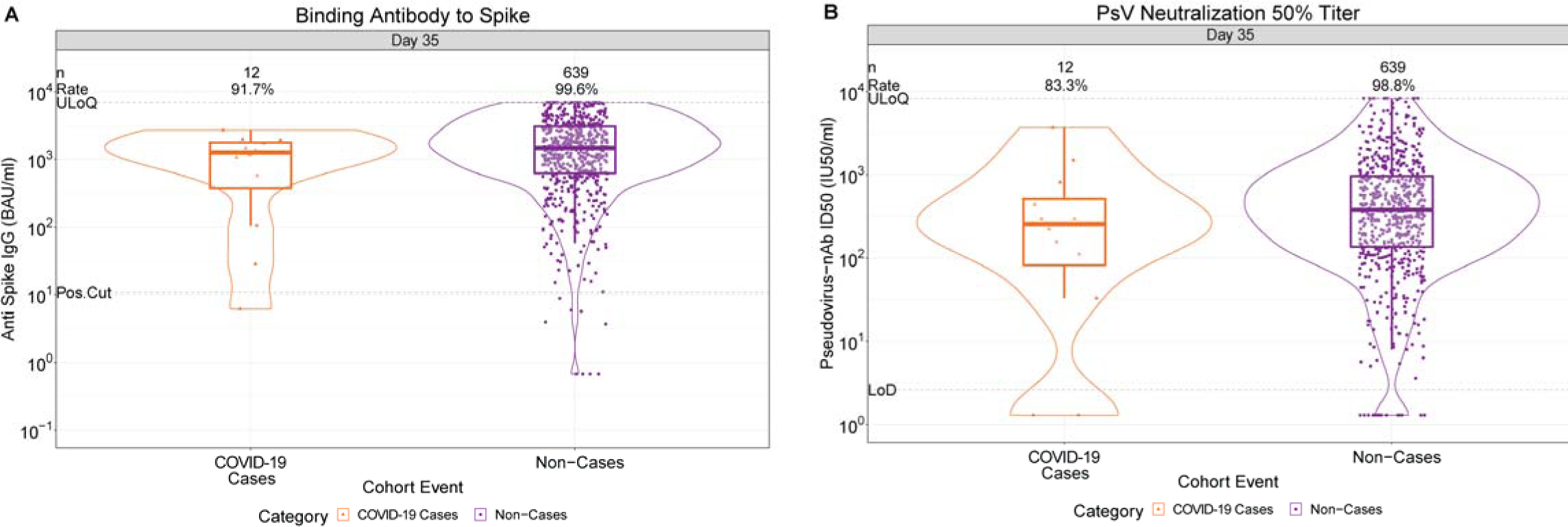
D35 antibody marker level by COVID-19 outcome status in baseline SARS-CoV- 2 negative per-protocol vaccine recipients (U.S. study sites). (A) Anti-spike IgG concentration and (B) pseudovirus (PsV) neutralization ID50 titer. The violin plots contain interior box plots with upper and lower horizontal edges the 25^th^ and 75^th^ percentiles of antibody level and middle line the 50^th^ percentile, and vertical bars the distance from the 25^th^ (or 75^th^) percentile of antibody level and the minimum (or maximum) antibody level within the 25^th^ (or 75^th^) percentile of antibody level minus (or plus) 1.5 times the interquartile range. At both sides of the box, a rotated probability density curve estimated by a kernel density estimator with a default Gaussian kernel is plotted. Frequencies of participants with positive spike IgG/detectable nAb ID50 responses were computed with inverse probability of sampling weighting (reported at the top of the plots as “Rate”). Pos.Cut, Positivity cut-off for spike IgG defined by IgG > 10.8424 BAU/ml, the assay positivity cut-off. ULoQ = 6934 BAU/ml for spike IgG. Seroresponse for ID50 was defined by a detectable value > limit of detection (LOD) (2.612 IU50/ml). ULoQ = 8319.938 IU50/ml. Cases experienced the primary COVID-19 endpoint starting 7 days post D35 visit through to the data cut (April 19, 2021). Non-cases are sampled into the immunogenicity subcohort with no evidence of SARS-CoV-2 infection (i.e., never tested RT-PCR positive) up to the end of the correlates study period (the data cut-off date April 19, 2021).

**Table 1.** D35 antibody marker SARS-CoV-2 seroresponse rates and geometric means in the U.S. cohort by COVID-19 outcome status. Analysis based on baseline SARS-CoV-2 negative per-protocol vaccine recipients in the case-cohort set. Median (interquartile range) days from vaccination to D35 was 38 (6).

|  |  | COVID-19 Cases<sup>1</sup> |  | Non-Cases in Immunogenicity Subcohort<sup>2</sup> |  |  | Comparison |  |
| --- | --- | --- | --- | --- | --- | --- | --- | --- |
| D35 Marker | N | Proportion with Antibody Response<sup>3</sup><br>(95% CI) | Geometric Mean (GM)<br>(95% CI) | N | Proportion with Antibody Response<sup>3</sup><br>(95% CI) | Geometric Mean (GM) (95 % CI) | Response Rate<br>Difference (Non-Cases – Cases) | Ratio of GM<br>(Non-Cases/<br>Cases) |
| Anti Spike IgG<br>(BAU/ml) | 12 | 91.7%<br>(52.5%, 99.1%) | 528<br>(184, 1513) | 639 | 99.6%<br>(99.2%, 99.8%) | 1552<br>(1407, 1713) | 8.0% (0.5%, 47.2%) | 2.9 (1.0, 8.3) |
| Pseudovirus-nAb<br>ID50 (IU50/ml) | 12 | 83.3%<br>(47.6%, 96.5%) | 135<br>(35, 519) | 639 | 98.8%<br>(97.5%, 99.4%) | 461<br>(404, 526) | 15.4% (2.2%, 51.2%) | 3.4 (0.9, 12.5) |

<sup>1</sup>Cases are baseline SARS-CoV-2 seronegative per-protocol vaccine recipients with the primary COVID-19 endpoint (symptomatic RT-PCR-confirmed COVID-19) starting 7 days post D35 visit through to the data cut (April 19, 2021).
<sup>2</sup>Non-cases are baseline negative per-protocol vaccine recipients sampled into the immunogenicity subcohort with no evidence of SARS-CoV-2 infection (i.e., never tested RT-PCR positive) up to the end of the correlates study period (the data cut-off date April 19, 2021).
<sup>3</sup>Antibody response defined by IgG concentration above the assay positivity cut-off (10.8424 BAU/ml) or by detectable ID50 > limit of detection (LOD) = 2.612 IU50/ml.

The two D35 markers were moderately-to-highly correlated (Spearman rank r = 0.80) (Figure 2). For each D35 marker, the reverse cumulative distribution function curve in the context of the overall vaccine efficacy estimate is shown in Supplementary Figure 3. As expected, because the analyzed cohort is baseline SARS-CoV-2 negative, frequencies of placebo recipients with positive or detectable responses at D35 were near zero (among non-cases 0.9% for spike IgG and 0.2% for nAb ID50) (Supplementary Table 4).

**Figure 2.**
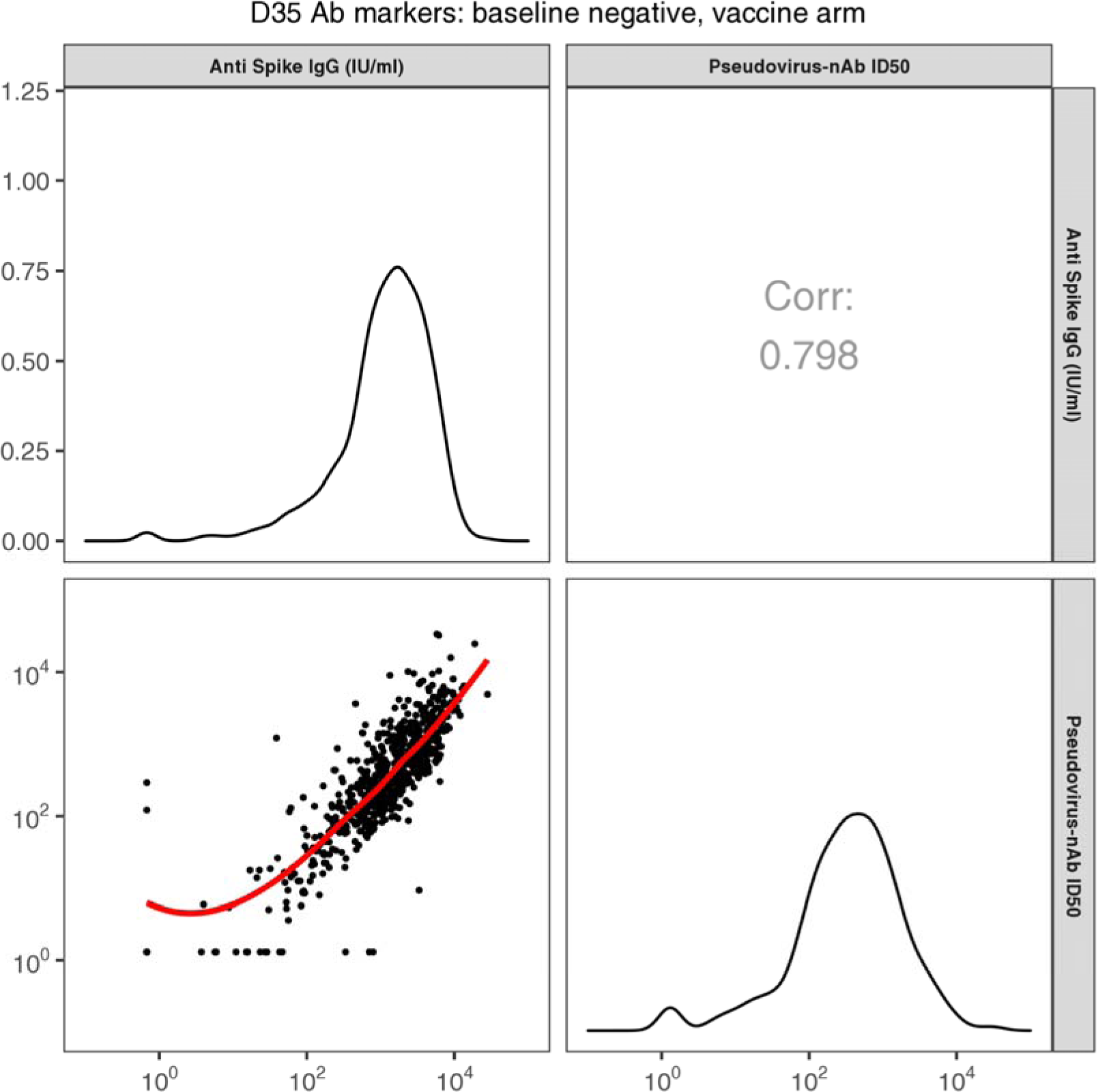
Scatterplot of Day 35 spike IgG vs. pseudovirus (PsV)-nAb ID50 values for baseline SARS-CoV-2 negative per-protocol vaccine recipients in the immunogenicity subcohort (U.S. study sites). Corr, Spearman rank correlation.

### D35 spike IgG concentration and neutralization ID50 titer are inversely correlated with risk of COVID-19 in vaccine recipients

The cumulative incidence of COVID-19 for vaccine recipient subgroups defined by D35 antibody marker tertiles suggest that COVID-19 risk decreased with increasing tertiles for both antibody markers (Figure 3), although given the small number of breakthrough cases the SAP specified not conducting hypothesis tests for tertile correlates. There were 5, 7, and 0 breakthrough cases in the Low, Medium, and High D35 spike IgG antibody subgroups, with point estimates of marginalized hazard ratios 1.48 (95% CI: 0.43, 5.10) for Medium vs. Low and 0.0 (0.0, Not Calculated) for High vs. Low. There were 8, 2, and 2 breakthrough cases in the Low, Medium, and High D35 nAb ID50 antibody groups, with point estimates of marginalized hazard ratios 0.26 (0.05, 1.41) for Medium vs. Low and 0.25 (0.05, 1.33) for High vs. Low; thus point estimates (with wide confidence intervals) indicate about 4-fold lower risk for vaccine recipients with nAb ID50 above the 33^rd^ percentile compared to those with the lowest tertile nAb ID50 titers.

**Figure 3.**
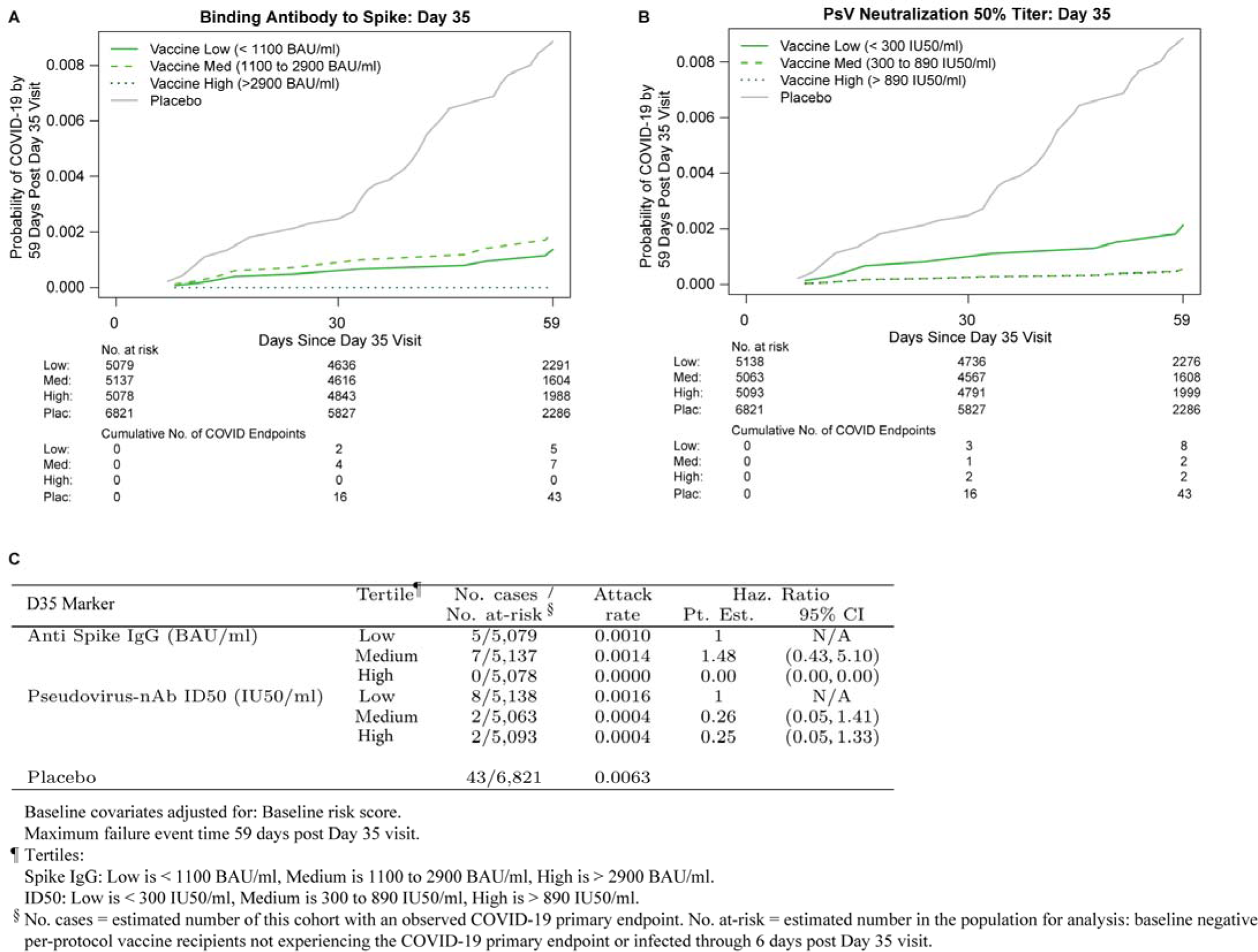
COVID-19 risk by D35 antibody marker level in baseline SARS-CoV-2 negative per-protocol vaccine recipients (U.S. study sites). Baseline risk score-adjusted cumulative incidence of COVID-19 by Low, Medium, High tertile of D35 antibody marker level. (A, C) Anti-spike IgG concentration; (B, C) pseudovirus (PsV) neutralization ID50 titer. No p-values for hypothesis tests are used because of the small number of COVID-19 breakthrough cases, as specified in the Statistical Analysis Plan.

The SAP specified hypothesis tests for whether the quantitative D35 markers correlated with risk of COVID-19. Both markers significantly inversely correlated with risk [estimated hazard ratio per 10-fold increase in IgG spike concentration of 0.36 (0.20, 0.63), p<0.001; estimated hazard ratio per 10-fold increase in nAb ID50 titer of 0.39 (0.19, 0.82), p=0.013] (Table 2A). Both markers passed the specified multiple testing correction [family-wise error rate (FWER)- adjusted p=0.005 and p=0.030 for spike IgG and nAb ID50, respectively]. When the results are placed on the hazard ratio scale, per standard deviation increase in marker value to aid comparability of the correlates, the correlate strengths were similar: hazard ratio 0.52 (0.36, 0.75) for spike IgG and 0.49 (0.28, 0.86) for nAb ID50 (Table 2B). The narrower confidence interval for spike IgG may reflect lower technical measurement variability for the assay of this marker.

**Table 2.**
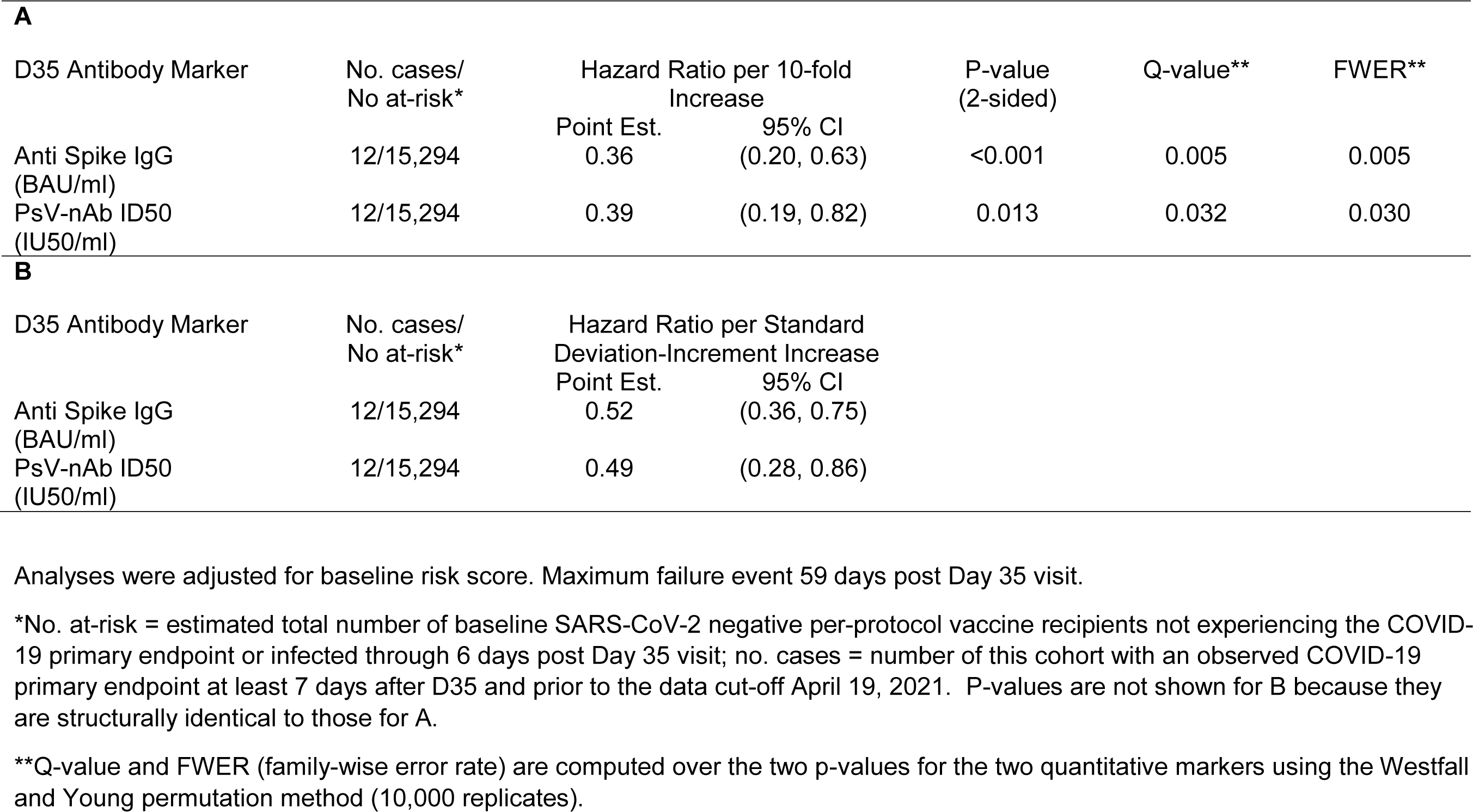
Hazard ratios of COVID-19 (A) per 10-fold increase or (B) per standard deviation increase in each D35 marker in baseline negative per-protocol vaccine recipients (U.S. study sites).

| **A** |  |  |  |  |  |  |
| --- | --- | --- | --- | --- | --- | --- |
| D35 Antibody Marker | No. cases/<br>No at-risk* | Hazard Ratio per 10-fold<br>Increase |  | P-value<br>(2-sided) | Q-value** | FWER** |
|  |  | Point Est. | 95% CI |  |  |  |
| Anti Spike IgG<br>(BAU/ml) | 12/15,294 | 0.36 | (0.20, 0.63) | <0.001 | 0.005 | 0.005 |
| PsV-nAb ID50<br>(IU50/ml) | 12/15,294 | 0.39 | (0.19, 0.82) | 0.013 | 0.032 | 0.030 |
| **B** |  |  |  |  |  |  |
| D35 Antibody Marker | No. cases/<br>No at-risk* | Hazard Ratio per Standard<br>Deviation-Increment Increase |  |  |  |  |
|  |  | Point Est. | 95% CI |  |  |  |
| Anti Spike IgG<br>(BAU/ml) | 12/15,294 | 0.52 | (0.36, 0.75) |  |  |  |
| PsV-nAb ID50<br>(IU50/ml) | 12/15,294 | 0.49 | (0.28, 0.86) |  |  |  |

Analyses were adjusted for baseline risk score. Maximum failure event 59 days post Day 35 visit.
\*No. at-risk = estimated total number of baseline SARS-CoV-2 negative per-protocol vaccine recipients not experiencing the COVID-19 primary endpoint or infected through 6 days post Day 35 visit; no. cases = number of this cohort with an observed COVID-19 primary endpoint at least 7 days after D35 and prior to the data cut-off April 19, 2021. P-values are not shown for B because they are structurally identical to those for A.
\*\*Q-value and FWER (family-wise error rate) are computed over the two p-values for the two quantitative markers using the Westfall and Young permutation method (10,000 replicates).

Panels A and B in Figure 4 show the marginalized Cox modeling results in terms of estimated cumulative incidence of COVID-19 (from 7 to 59 days post-D35) across D35 marker levels. For each antibody marker, COVID-19 cumulative incidence/risk decreased as antibody marker level increased. Across the range of D35 spike IgG concentrations, estimated risk decreased from 0.0030 (0.0007, 0.00063) at low concentration spike IgG = 100 BAU/ml (3rd percentile) to 0.0005 (0.0002, 0.0009) at 6934 BAU/ml, a 6-fold change in risk level (Figure 4A). For D35 nAb ID50, estimated risk decreased from 0.0022 (0.0006, 0.0040) at low nAb ID50 titer = 50 IU50/ml (3rd percentile) to 0.0003 (0.0001, 0.0010) at the highest value evaluated 7230 IU50/ml, a 7.3- fold change in risk level (Figure 4B).

**Figure 4.**
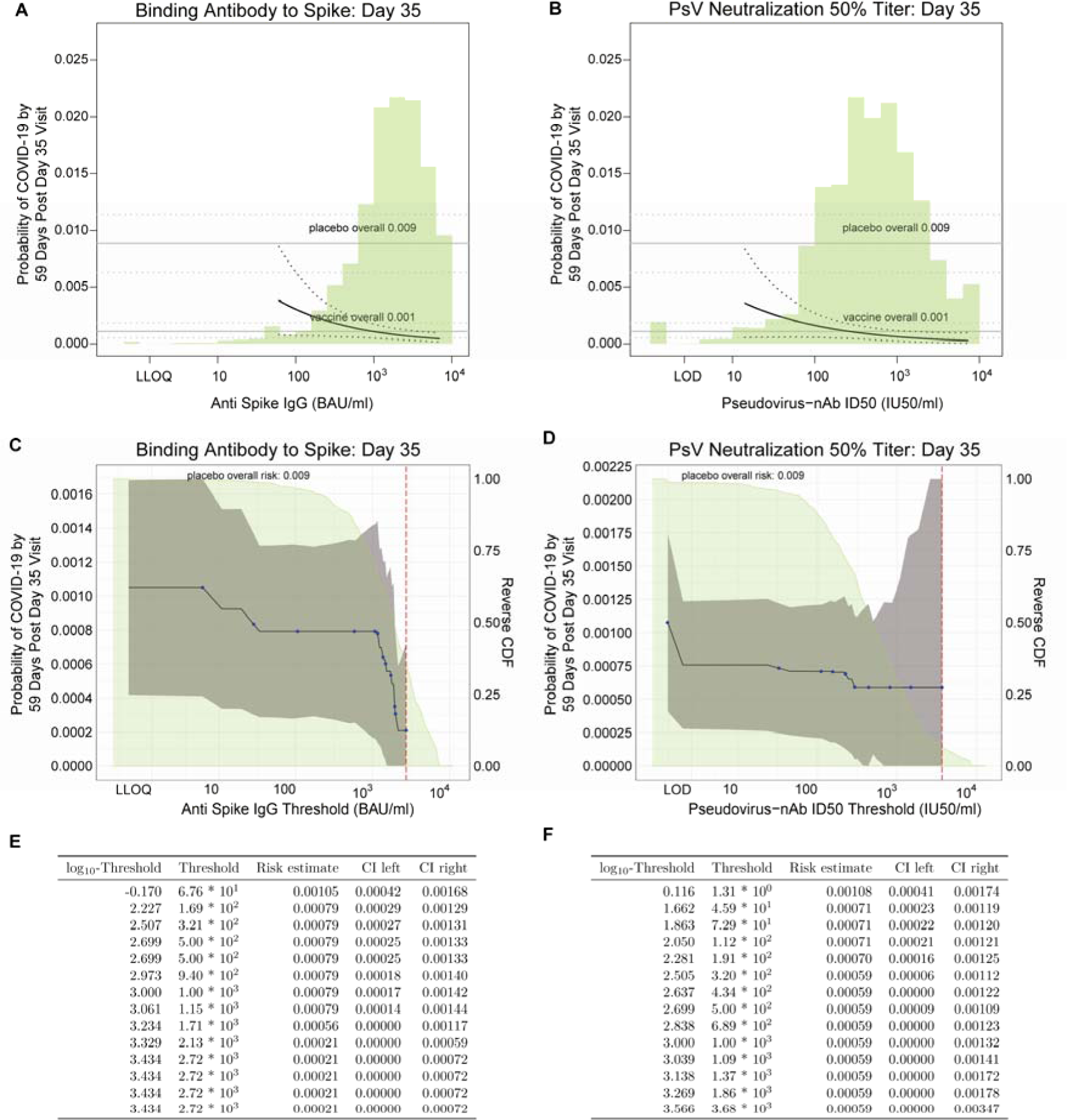
Analyses of D35 antibody markers as a correlate of risk in baseline SARS-CoV-2 negative per-protocol vaccine recipients (US study sites). (A, B) Baseline risk score- adjusted cumulative incidence of COVID-19 by 59 days post D35 by D35 (A) anti-spike IgG or (B) pseudovirus (PsV)-nAb ID50 titer, estimated using a marginalized Cox model. The dotted black lines indicate bootstrap pointwise 95% CIs. The upper and lower horizontal gray lines are the overall cumulative incidence of COVID-19 from 7 to 59 days post D35 in placebo and vaccine recipients, respectively. Curves are plotted over the antibody marker range from the 2.5^th^ percentile to the 97.5^th^ percentile: 60.9 to 6934 BAU/ml for spike IgG (where 6934 is the ULOQ) and 14.2 to 7230 IU50/ml for PsV-nAb ID50. (C-F) Baseline risk score-adjusted cumulative incidence of COVID-19 by 59 days post D35 by D35 (C, E) anti-spike IgG or (D, F) PsV-nAb ID50 titer above a threshold. The blue dots are point estimates at each COVID-19 primary endpoint linearly interpolated by solid black lines; the gray shaded area is pointwise 95% confidence intervals (CIs). The estimates and CIs assume a non-increasing threshold-response function. The upper boundary of the green shaded area is the estimate of the reverse cumulative distribution function (CDF) of D35 antibody marker level. The vertical red dashed line is the D35 antibody marker threshold above which no COVID-19 endpoints occurred (in the time frame of 7 days post D35 through to the data cut-off date April 19, 2021). (E, F) D35 antibody marker thresholds, risk estimates, and 95% confidence intervals corresponding to the blue dots in panels C and D, respectively. PsV, pseudovirus.

When vaccine recipients were divided into subgroups defined by their D35 antibody marker level above a specific threshold and varying the threshold over the range of values, nonparametric regression showed that the cumulative incidence of COVID-19 (from 7 to 59 days post-D35) decreased with each of the D35 markers (panels C and D in Figure 4). Risk decreased from 0.00105 (0.00042, 0.00168) in all vaccine recipients to 0.00021 (0, 0.00059) for vaccine recipients with spike IgG concentration > 2130 BAU/ml, and to 0.00059 for vaccine recipients with nAb ID50 > 320 IU50/ml. Additional nAb ID50 threshold increases did not reduce risk further. These results suggest that for NVX-CoV2373, high spike IgG appears to be a better marker of very low risk than high nAb ID50. Panels E and F in Figure 4 provide tables of risk estimates corresponding to the plots in panels C and D, respectively.

### Vaccine efficacy increases with D35 spike IgG concentration and neutralization ID50 titer

Figure 5 shows estimated vaccine efficacy against COVID-19 (from 7 to 59 days post-D35) across a range of levels of each D35 antibody marker. [The causal parameter being estimated is one minus the probability of COVID-19 by 59 days for the vaccine group supposing the D35 marker is set to a given level or all vaccine recipients, divided by this probability for the placebo arm; see the SAP (Section 11) for details.] Estimated vaccine efficacy increased with the level of each D35 marker. At three selected D35 IgG concentration values covering the span of values (100, 1000, and 6934 BAU/ml), estimated VE was 65.5% (23.0%, 90.8%), 87.7% (77.7%, 94.4%), and 94.8% (88.0%, 97.9%). At four selected nAb ID50 titers covering the span of values (50, 100, 1000, and 7230 IU50/ml), estimated VE was 75.7% (49.8%, 93.2%), 81.7% (66.3%, 93.2%), 92.8% (85.1%, 97.4%) and 96.8% (88.3%, 99.3%). A causal sensitivity analysis using the same methodology and implementation as used in the correlates analyses of the COVE and ENSEMBLE trials supported that vaccine efficacy increased with each marker after accounting for potential unmeasured confounding of the effect of the D35 antibody marker on occurrence of COVID-19 (Supplementary Figure 4).

**Figure 5.**
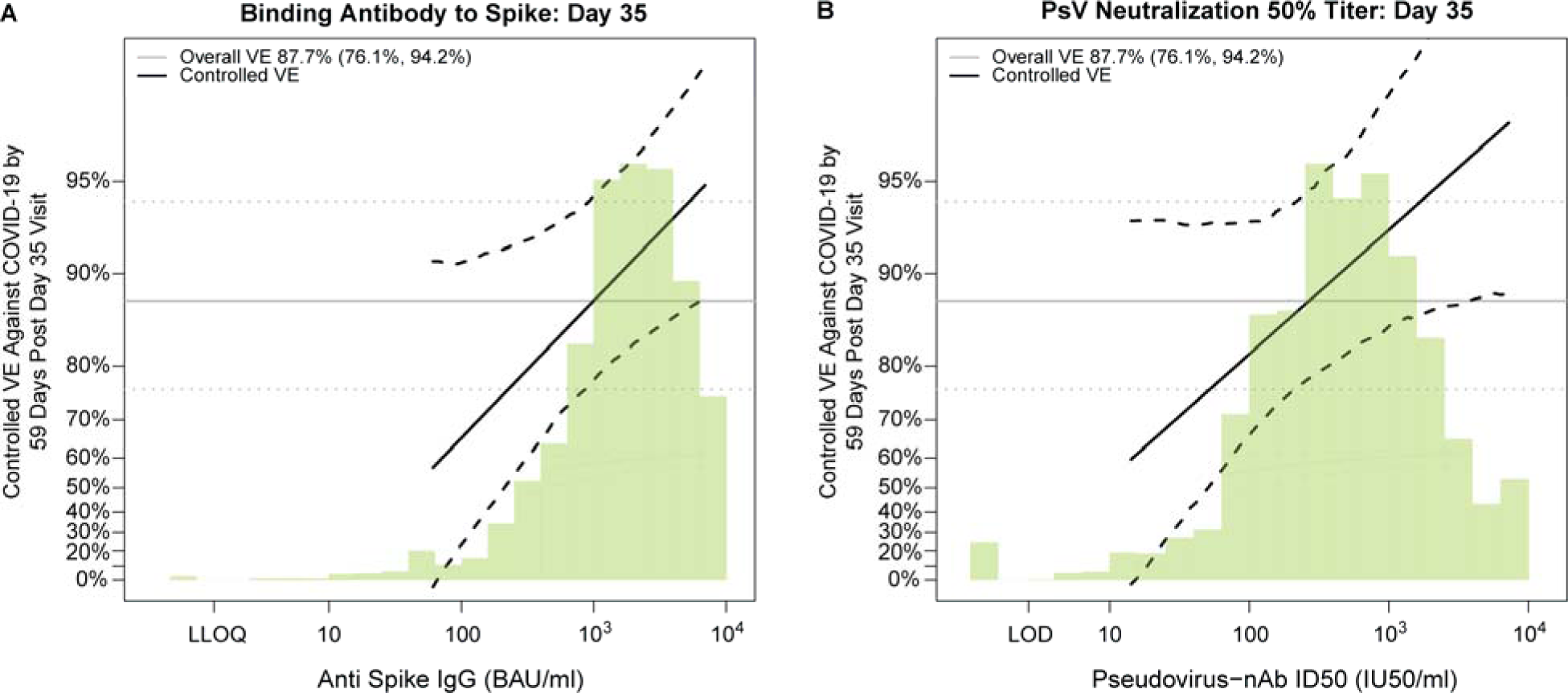
Vaccine efficacy by D35 antibody marker level in baseline SARS-CoV-2 negative per-protocol vaccine recipients (U.S. study sites). Curves shown are for D35 (A) anti-spike IgG concentration or (B) pseudovirus (PsV)-nAb ID50 titer. The dotted black lines indicate bootstrap pointwise 95% confidence intervals. The green histogram is an estimate of the density of D35 antibody marker level and the horizontal gray line is the overall vaccine efficacy from 7 to 59 days post D35, with the dotted gray lines indicating the 95% confidence intervals. Analyses adjusted for baseline risk score. Curves are plotted over the antibody marker range from the 2.5th percentile to the 97.5^th^ percentile: 60.9 to 6934 BAU/ml for anti-spike IgG (where 6934 is the ULOQ) and 14.2 to 7230 IU50/ml for PsV-nAb ID50.

### Comparing the antibody markers as correlates of risk and protection across phase 3 trials/vaccine platforms

We next compared the correlates of risk and the vaccine efficacy-by-antibody marker curves from PREVENT-19 to those estimated from three other randomized, placebo-controlled COVID- 19 vaccine efficacy trials: COVE (two doses of Moderna mRNA-1273 at D1 and D29),^24^ ENSEMBLE (one dose of Janssen Ad26.COV2.S at D1),^30^ and COV002 in the United Kingdom^31^ (two doses of AstraZeneca AZD1222/ChAdOx1 nCoV-19 at D0 and D28). In these three studies the antibody markers were measured 4 weeks post-vaccination, compared to 2 weeks post-vaccination in PREVENT-19. In this comparison for ENSEMBLE we restricted to data from the U.S. study sites (ENSEMBLE-US) in order to match the fact that the correlates analyses of PREVENT-19 and COVE both restricted to the U.S. Direct comparison of correlates of risk and vaccine efficacy at a given spike IgG concentration values across the four trials is possible because the Meso Scale Discovery (MSD) assay at Nexelis that was used in PREVENT-19, the MSD assay at VRC that was used in COVE^25^ and ENSEMBLE,^26^ and the MSD assay at PPD that was used in COV002,^27^ all had the original assay readout (in Arbitrary Units/ml) transformed to WHO International Standard 20/136 international units (BAU/ml scale).^32, 33^ Direct comparison of correlates of risk and vaccine efficacy at a given nAb ID50 titer in PREVENT-19 to results at the same nAb ID50 titer in COVE is possible because the Duke assay (used in COVE) and the Monogram assay (used in PREVENT-19) underwent concordance testing^25, 34^ and were calibrated to the WHO IS 20/136 (described in refs.^25, 34^) to be expressed in IU50/ml. A similar comparison can be performed vs. the ENSEMBLE and COV002 nAb ID50 values because the same pseudovirus neutralization assay (Monogram) was used to assay samples from the PREVENT-19, ENSEMBLE, and COV002 trials.^27^

Table 3 compares the inverse correlates of risk results for spike IgG and for nAb ID50 across the three USG-supported trials. The estimated strength of the IgG spike correlate of risk is strongest in PREVENT-19 (hazard ratio 0.36) compared to COVE (hazard ratio 0.66) and ENSEMBLE-U.S. (hazard ratio 0.62. The estimated strength of the nAb ID50 correlate of risk is comparable in the three trials (hazard ratios 0.39, 0.42, 0.38, respectively). The IgG spike marker passes FWER-correction for being a significant correlate in 2 of the 3 trials (exception ENSEMBLE-US) and the nAb ID50 marker passes FWER-correction for being a significant correlate in all 3 trials. Estimated vaccine efficacy increased with increasing spike IgG concentration in each trial (Figure 6A). The figure shows that the distribution of spike IgG was similar for the NVX-CoV2373 and mRNA-1273 vaccines, slightly lower for the former (geometric mean concentration 1552 vs. 2652 BAU/ml). The dynamic range of this marker among NVX- CoV2373 recipients is wider than in mRNA-1273 recipients, which may explain the strong statistical significance of the PREVENT-19 results (p < 0.001), stronger than COVE (p=0.005). The apparently stronger correlate of protection for NVX-CoV2373 than for mRNA-1273 is indicated by the steeper estimated vaccine efficacy curve. This figure also supports that spike IgG at 1000 BAU/ml or higher is associated with high vaccine efficacy (at least 85-90%) for both NVX-CoV2373 and mRNA-1273. In addition, the figure supports similar vaccine efficacy curves for NVX-CoV2373 and AZD1222 over the range of spike IgG concentrations that overlap and hence can be compared (IgG concentration about 50 to 800 BAU/ml). The AD26.COV2.S vaccine induced spike IgG levels an order of magnitude lower than those induced by NVX- CoV2373, limiting the ability to compare the vaccine efficacy curves between these vaccines.

**Figure 6.**
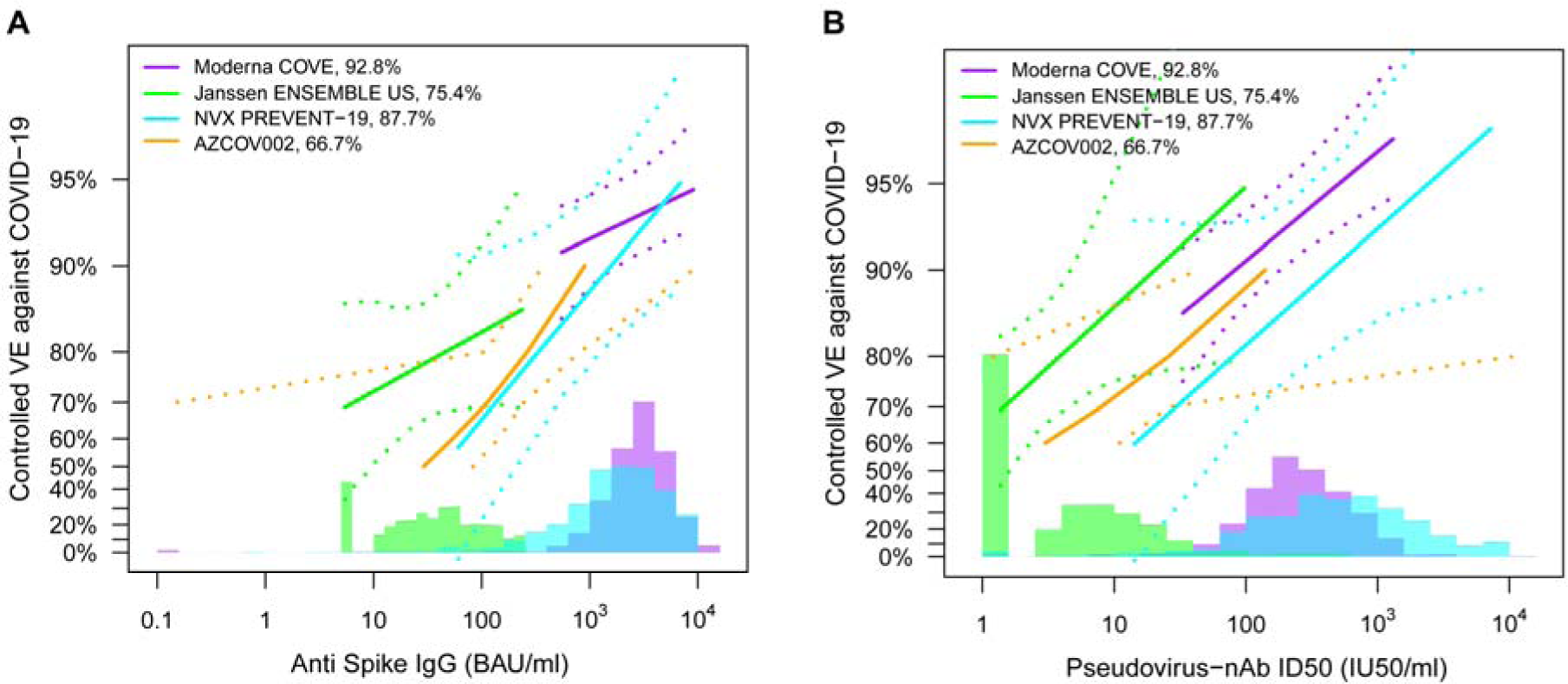
Vaccine efficacy (solid lines) in baseline SARS-CoV-2 negative per-protocol participants by post-vaccination antibody marker level in four randomized, placebo- controlled COVID-19 vaccine efficacy trials. Vaccine efficacy was estimated using the marginalized Cox proportional hazards implementation of Gilbert et al.^42^ Vaccine efficacy (VE) estimates are shown by A) anti-spike IgG concentration [D57 in COVE, D29 in ENSEMBLE- United States sites (ENSEMBLE-US), D35 in PREVENT-19, D56 in COV002] or by B) pseudovirus (PsV)-nAb ID50 titer (D57 in COVE, D29 in ENSEMBLE-US,D35 in PREVENT-19, D56 in COV002). The dashed lines indicate bootstrap point-wise 95% confidence intervals. The follow-up periods for the VE assessment were: COVE (doses D1, D29), 7 to 100 days post D57; ENSEMBLE-US (one dose, D1), 1 to 53 days post D29; PREVENT-19 (doses D0, D21), 7 to 59 days post D35; COV002 (doses D0, D28; 28 days post D28 until the end of the study period). The histograms are an estimate of antibody marker density in baseline SARS-CoV-2 negative per-protocol vaccine recipients. Curves are plotted over the following antibody marker ranges: A) COVE: 2.5^th^ percentile to 97.5th percentile of marker, ENSEMBLE-US: 2.5^th^ percentile to 97.5^th^ percentile, PREVENT-19: 2.5^th^ percentile to 97.5^th^ percentile, COV002: 29 to 899 BAU/ml; B) COVE: 10 IU50/ml to 97.5th percentile of marker, ENSEMBLE-US: 2.5^th^ percentile to 97.5^th^ percentile, PREVENT-19: 2.5^th^ percentile to 97.5^th^ percentile, COV002: 3 to 140 IU50/ml. Baseline covariates adjusted for were: COVE: baseline risk score, comorbidity status, and Community of color status; ENSEMBLE-US, baseline risk score; PREVENT-19: baseline risk score; COV002: baseline risk score.

**Table 3.** Comparison of correlate of risk results for spike IgG and PsV-nAb ID50 across three randomized, placebo- controlled COVID-19 vaccine efficacy trials (U.S. study sites).

| Vaccine Platform* | Trial | Ab Marker 4 Wks Post-Vaccination | Follow-Up Post Vaccination | Estimated Hazard Ratio per 10-fold Increase in the Marker (95% CI) | P-Value | Q-Value | FWER-Adjusted P-Value |
| --- | --- | --- | --- | --- | --- | --- | --- |
| mRNA | COVE | Spike IgG | 126 days | 0.66 (0.50, 0.88) | 0.005 | 0.014 | 0.010 |
| Ad26 | ENSEMBLE-US | Spike IgG | 83 days | 0.62 (0.28, 1.37) | 0.24 | 0.35 | 0.36 |
| Recombinant Protein | PREVENT-19 | Spike IgG | 73 days | 0.36 (0.20, 0.64) | < 0.001 | 0.005 | 0.005 |
| mRNA | COVE | PsV-nAb ID50 | 126 days | 0.42 (0.27, 0.65) | < 0.001 | 0.002 | 0.003 |
| Ad26 | ENSEMBLE-US | PsV-nAb ID50 | 83 days | 0.38 (0.13, 1.12) | 0.078 | 0.22 | 0.20 |
| Recombinant Protein | PREVENT-19 | PsV-nAb ID50 | 73 days | 0.39 (0.19, 0.82) | 0.013 | 0.032 | 0.030 |

\* COVE: Moderna mRNA-1273 spike vaccine; ENSEMBLE: Janssen Ad26 vector spike vaccine Ad26.CoV2.S; PREVENT-19: Novavax recombinant spike protein vaccine NVX-CoV2373.

Estimated vaccine efficacy also increased with increasing nAb ID50 titer in each trial (Figure 6B). The distribution of nAb ID50 titer was similar for the NVX-CoV2373 and mRNA-1273 vaccines, slightly higher for the former (geometric mean concentration 461 vs. 247 IU50/ml). Again the dynamic range of this marker among NVX-CoV2373 recipients is wider than that among mRNA-1273 recipients. However, the results suggest comparable-strength correlates of protection for NVX-CoV2373 and mRNA-1273 as indicated by comparably-steep estimated vaccine efficacy curves and the similar hazard ratios (0.39 and 0.42, respectively) per 10-fold nAb ID50 increase. The vaccine efficacy point estimates suggest that higher nAb ID50 titers may be needed to achieve the same level of high vaccine efficacy for NVX-CoV2373 than for mRNA-1273, but given the wide 95% confidence intervals this cannot be inferred statistically. Over the regions of overlapped titers, Figure 6B also tentatively suggests that higher nAb ID50 titers for NVX-CoV2373 may be needed to achieve the same level of high vaccine efficacy as seen for the AD26.COV2.S and AZD1222 vaccines; the fact that the NVX-CoV2373 nAb ID50 titers are considerably higher than those for the viral vector vaccines may explain why the overall vaccine efficacy is higher for NVX-CoV2373. Overall, the comparative results are consistent with a correlate of protection model that attributes differences in overall vaccine efficacy observed across the four regimens to differences in the distribution of spike IgG or of nAb ID50 among the four vaccine regimens, consistent with the results of meta-analyses.^35, 36^

## Discussion

RNAThe immune correlates analysis of baseline SARS-CoV-2 per-protocol participants enrolled at U.S. study sites of the PREVENT-19 trial of the NVX-CoV2373 vaccine vs. placebo showed that both anti-spike IgG concentration and pseudovirus neutralization ID50 titer measured two weeks post second dose at the D35 study visit were significant inverse correlate of risk of symptomatic COVID-19 occurrence over the subsequent 7 to 59 days. In addition, vaccine efficacy increased with higher D35 antibody marker levels, such that vaccine efficacy was estimated to be 66% for vaccine recipients with low spike IgG value of 100 BAU/ml (3^rd^ percentile), increasing to 95% at the highest quantifiable antibody level of 6934 BAU/ml (the assay ULOQ). Similarly, vaccine efficacy was estimated at 76% for vaccine recipients with low nAb ID50 value of 50 IU50/ml (3^rd^ percentile) and increased to 97% at highest antibody levels of 7230 IU50/ml (97.5^th^ percentile). With only 12 vaccine breakthrough cases in the PREVENT-19 trial, it is remarkable that both markers passed the multiple hypothesis testing correction for being inverse correlates of risk, indicating that the estimated correlations were quite strong. The limited precision in statistical inferences (i.e., wide confidence intervals) resulting from only 12 cases illustrates the challenges in deriving precise estimates of vaccine efficacy to specific values of the antibody markers. Rather, we propose that the results be interpreted more qualitatively as providing strong support that both markers are immune correlates, and additional studies with more breakthrough cases are needed to more precisely define the quantitative relationship between the levels of each antibody marker and vaccine efficacy.

Along the same vein, the level of precision of the data limits the ability to uncover differences in the performance of the two antibody markers, and in terms of point estimates the results are similar, with an indication from the nonparametric threshold analysis (Figure 4 Panels C and D) that spike IgG may be a more discriminating correlate. The apparent similarity of results is in part explained by the fairly high correlation between the two markers (Spearman rank correlation 0.80). The spike IgG marker had greater precision in the correlates analyses in terms of narrower confidence intervals around the hazard ratios, marker-specific probabilities of COVID-19, and marker-specific vaccine efficacy; for example 95% confidence intervals about hazard ratios per SD increase in the marker were 0.36 to 0.75 for spike IgG and 0.28 to 0.86 for nAb ID50. Based on the ratio of squared standard errors to estimated log hazard ratio coefficient, this difference in confidence interval width constitutes an approximately 2.3-fold efficiency advantage for spike IgG, which suggests that 2.3 times more vaccine breakthrough cases being needed for a correlates analysis of nAb ID50 to achieve the same confidence interval width about the hazard ratio as a correlates analysis of spike IgG.^35^ The narrower confidence intervals for spike IgG likely reflects the lower technical measurement variability of this marker (as reflected by lower %CV in assay validation studies). This lower measurement variability is a known advantage of an ELISA-type assay, with successful track record of use as a correlate or protection/surrogate endpoint for many licensed vaccines.^11^ However, our analysis could only evaluate correlates for COVID-19 with SARS-CoV-2 strains that were circulating during the trial, which amounted to strains that were genetically close to the ancestral Wuhan-Hu-1 vaccine strain (non-variant strains) and variants of concern and variants of interest that started to differ phylogenetically from Wuhan-Hu-1-like strains. Additional research is needed to understand which assay performs best as a correlate of risk and protection against SARS-CoV-2 strains that are more genetically divergent from the vaccine strain, such as strains from the delta and omicron variants of concern.

Because the NVX-CoV2373 vaccine induced antibody responses by D35 in almost all vaccine recipients (positive spike IgG for 99.6% of vaccine recipients and detectable nAb ID50 titers for 98.8% of vaccine recipients), it was not possible to assess the proportion of vaccine efficacy mediated through either of these markers. Nonetheless, the fact that vaccine efficacy was estimated at 66-76% at the lowest evaluable antibody values generates the hypothesis that these markers did not fully mediate the vaccine’s efficacy: other immune responses or immune markers at other time points (e.g. anamnestic responses) or not quantifiable in serum likely contributed to vaccine efficacy (including memory B cells, Fc effector functions, and T cell responses^20^).

Strengths of the study include the pre-specification of analyses that makes p-values and confidence intervals valid and increases confidence in the conclusions; and the fact that the phase 3 trial was randomized and double-blinded throughout the period of follow-up. Moreover, as in other phase 3 trials, the restriction to participants who were SARS-CoV-2 negative at enrollment aided interpretability by ensuring that the antibody markers measured only vaccine-elicited antibodies, as opposed to a mixture of vaccine-elicited antibodies and pre-existing infection-induced antibodies. The USG effort to develop COVID-19 vaccine correlates of protection has prioritized planning and execution of harmonized design and analysis,^22^ to enable comparisons of correlates across trials and to combine data for meta-analysis correlates assessment. The PREVENT-19, COVE and ENSEMBLE correlates studies share harmonized trial protocols and study populations, restriction of the analysis to the randomized, placebo-controlled double-blind follow-up period, use of a two-phase case-cohort antibody marker sampling design, application of the same open-source, reproducibly-implemented statistical methods, and use of validated immunoassays with the analyzed readouts placed on a common WHO International Units scale to enable comparability of results.^22^

The PREVENT-19 correlates results supported that both spike IgG and nAb ID50 performed similarly as correlates as previously found in the mRNA-1273 COVE correlates study,^24^ encouraging consideration of the use of a common immune correlate for applications to both recombinant protein and mRNA COVID-19 vaccines. Comparisons of PREVENT-19 correlates results to the ENSEMBLE and COV002 correlates results are more limited, given the lower degree of overlap of antibody levels between the NVX-CoV2373 vaccine compared to the Ad26.COV2.S and AZD1222 vaccines. While estimates of vaccine efficacy at the same marker values were lower for NVX-CoV2373 than for the adenovirus vector vaccines, the wide confidence intervals precludes statistical inferences about whether higher antibody levels are needed to achieve similarly high levels of vaccine efficacy. The fact that PREVENT-19 measured the antibody markers 2 weeks post-vaccination whereas COVE, ENSEMBLE, and COV002 measured the antibody markers 4 weeks post-vaccination is a limitation of the comparisons. In addition, comparability is limited by the fact that the studies had somewhat different periods of follow-up during which different distribution of variants circulated, which could have created differences in distributions of antigenic distances of SARS-CoV-2 circulating strains to the vaccine strain.

We next discuss some additional limitations of this correlates study and their implications on future work. First, other NVX-CoV2373-induced immune responses of interest (e.g. spike-specific T-cell responses,^36^ Fc effector antibody functions^38^) were not assessed. A second limitation is the relatively short follow-up (two and a half months post dose two), such that correlates for longer-term COVID-19 could not be assessed. This short follow-up resulted from the need to institute a blinded cross-over phase to enable high-risk individuals to be vaccinated once other COVID-19 vaccines were shown to be efficacious and granted emergency use authorization by the Food and Drug Administration. Future work is being planned to assess, in the USG-supported trials, binding and pseudovirus neutralizing antibodies over time as exposure-proximal correlates of risk, which could generate new insights about mechanistic correlates of protection. In addition, amended PREVENT-19 protocols provided NVX-CoV2373 vaccination to willing placebo recipients and offered a third dose of NVX-CoV2373 to all trial participants, and the trial continues to monitor participants for the COVID-19 primary endpoint; however, placebo comparisons are no longer possible. Future work can assess levels of post dose three anti-omicron spike IgG and nAb ID50 titers against omicron spike-pseudotyped virus as correlates of risk and protection against COVID-19 caused by strains from the omicron variant. These analyses can also assess post dose three anti-Wuhan-Hu-1 spike IgG and D614G nAbs as correlates of risk and protection against COVID-19 caused by strains from the omicron variant, to understand whether and how much the immune correlate can be improved based on homologous titers instead of vaccine-strain titers.

None of the 12 vaccine breakthrough endpoints were severe, precluding the study of severe COVID-19 correlates, a topic of substantial interest. It was also not possible to study whether immune correlates varied among subgroups (e.g., defined by race, ethnicity, age, and co- existing condition). In addition, given the high global prevalence of SARS-CoV-2 infection, it will be important to understand correlates of protection specifically in previously infected individuals, and to understand how the correlates depend on details of previous infection history such as timing, frequency, symptomatology, geographical factors, and SARS-CoV-2 infecting strain.

Similarly, many persons who will receive the NVX-CoV2373 vaccine will have previously received another SARS-CoV-2 vaccine such as an mRNA vaccine, and understanding whether the correlates are influenced by the type of vaccine that elicited the immune responses or are invariant to vaccine type will help determine whether and how immune correlates can be applied across vaccine platforms (the ‘ideal correlate’ would be independent of both vaccine history and infection history, depending only on the measured immune marker level).

Overall, this work provides the first direct correlates analysis of a phase 3 trial of a recombinant protein COVID-19 vaccine, and supports that both binding and pseudovirus neutralizing antibody titers are a correlate of protection for the NVX-CoV2373 vaccine, a step forward toward defining a surrogate endpoint for this vaccine. Moreover, this work lays groundwork for future research on whether such a surrogate endpoint may also be applied for other recombinant protein COVID-19 vaccines, and possibly also for other COVID-19 vaccine platforms.

## Methods

### Trial design, study cohort, COVID primary endpoints, and case/non-case definitions

Enrollment for the PREVENT-19 trial began on December 27, 2020. A total of 29,949 participants were randomized (2:1 ratio) to receive two doses of NVX-CoV2373 or placebo, one each on Days 0 and 21. Of the participants who were randomized, 28,181 were randomized at U.S. study sites; all analyses are restricted to U.S. study sites. Serum samples were taken on D0 and on D35 for antibody measurements in subset analyses. D35 antibody measurements were evaluated as correlates against the symptomatic COVID-19 endpoint defined in the main text.

The correlates analysis included COVID-19 primary endpoints up to the data cut-off date of April 19^th^, 2021 (the same data cut-off date as that of the primary efficacy analysis^7^). Correlates analyses were performed in baseline SARS-CoV-2 negative participants in the per-protocol cohort, with the same definition of “per-protocol” as in Dunkle et al.,^7^ except that the endpoint timeframe in that publication started 7 days after dose 2. Within this correlates analysis cohort, cases were COVID-19 primary endpoints in vaccine recipients starting 7 days post D35 through to the data cut-off and non-cases/controls were vaccine recipients sampled into the immunogenicity subcohort with no evidence of SARS-CoV-2 infection (i.e., never tested RT-PCR positive) up to the end of the correlates study period (data cut-off date). The rationale for requiring 7 days post D35 is given in Results (“COVID-19 study endpoint”).

### Solid-phase electrochemiluminescence S-binding IgG immunoassay (ECLIA)

Serum IgG binding antibodies against spike (homologus vaccine strain antigen, i.e. Wuhan-Hu- 1) were quantitated by Nexelis using a validated solid-phase electrochemiluminescence S-binding IgG immunoassay as previously described.^25^ Within an assay run, each human serum test sample was added to the precoated wells in duplicates in an 8-point dilution series.

Conversion of arbitrary units/ml (AU/ml) readouts to bAb units/ml (BAU/ml) based on the World Health Organization 20/136 anti SARS-CoV-2 immunoglobulin International Standard^30^ was performed as previously described.^25^ Antibody seroresponse was defined as IgG concentration above the positivity cut-off 10.8424 BAU/ml. Values below the lower limit of quantitation (LLOQ = 1.35 BAU/ml) were assigned the value LLOQ/2. Supplementary Table 5 provides the assay limits.

### Pseudovirus neutralization assay

Neutralizing antibody activity was measured at Monogram in a validated assay^34^ utilizing lentiviral particles pseudotyped with full-length SARS-CoV-2 spike (homologous vaccine strain, i.e. Wuhan-Hu-1, with the D614G mutation). The lentiviral particles also contained a firefly luciferase (Luc) reporter gene, enabling quantitative measurement of infection via relative luminescence units (RLU). Supplementary Table 5 provides the assay limits. The limit of detection (LOD) was not formally defined; we denote the value corresponding to the starting dilution level of the assay as the LOD. Neutralizing antibody response was defined by detectable ID50 > LOD = 2.612 IU50/ml. Values below the LOD were assigned the value LOD/2. ID50 is reported in units calibrated to the 20/136 anti SARS-CoV-2 immunoglobulin International Standard.

### Ethics

The following Institutional Review Boards (IRBs)/Independent Ethics committees reviewed and approved the study: Western Copernicus Group IRB, US; Great Plains IRB, US; Comite de etica en investigacion del Instituto Nacional de Ciencias Medicas y Nutricion, Salvador Zubiran, Mexico; Comite de etica en investigacion de la Unidad de Atencion Medica e Investigacion en Salud S.C., Mexico; Comite de etica en investigacion del Instituto Nacional de Salud Publica, Mexico; Comite de etica en investigacion de Medica Rio Mayo S.C., Mexico; Comite de etica en investigacion del Hospital La Mision S.A. de C.V., Mexico. All necessary patient/participant consent has been obtained and the appropriate institutional forms have been archived.

### Statistical methods

All data analyses were performed as pre-specified in the SAP (Supplementary Material).

#### Case-cohort set included in the correlates analyses

A case-cohort^38^ sampling design was used to randomly sample participants for D0, D35 antibody marker measurements. This random sample was stratified by the following baseline covariates: randomization arm, baseline SARS-CoV-2 status, and 8 US baseline demographic covariate strata defined by all combinations of: underrepresented minority (URM) vs. non-URM/unknown, age 18-64 vs. age ≥ 65, and heightened risk for COVID-19 severe complications (see the SAP for details). There were also two Mexico baseline demographic covariate strata (age 18-64 vs. age ≥ 65); however, the analysis here restricts to the U.S. study sites only.

#### Covariate adjustment

All correlates analyses were adjusted for a baseline risk score defined as the logit of predicted COVID-19 risk built from machine learning of data from baseline SARS-CoV-2 negative per-protocol placebo recipients, where the predicted outcome was occurrence of the primary COVID-19 endpoint starting 7 days after the D35 visit. Ensemble learning was used to build this risk score, using age, sex, ethnicity, race, co-existing conditions, height, weight, and BMI as input variables. Risk score development was restricted to US participants. The baseline risk score had weak ability to predict COVID-19, with cross-validated area under the ROC curve (CV-AUC) 0.583 for the placebo arm and AUC 0.540 for the vaccine arm (Supplementary Figure 5).

#### Correlates of risk in vaccine recipients

All correlates of risk and protection analyses were performed in baseline SARS-CoV-2 negative per-protocol participants with no evidence of SARS-CoV-2 infection through 6 days post D35 visit and not right-censored by D35. For each D35 marker, the baseline risk score-adjusted hazard ratio of COVID-19 (across marker tertiles, per 10-fold increase in the quantitative marker, or per standard deviation-increment increase in the quantitative marker) was estimated using inverse probability sampling weighted Cox regression models with 95% CIs and Wald-based p-values (p-values were only computed for the quantitative markers given the small number of breakthrough cases). These Cox model fits were also used to estimate marker-conditional cumulative incidence of COVID-19 through 59 days post-D35 in baseline negative per-protocol vaccine recipients, with 95% CIs computed using the percentile bootstrap. The Cox models were fit using the survey package^39^ for the R language and environment for statistical computing.^40^ Point and 95% CI estimates about marker-threshold-conditional cumulative incidence were computed by nonparametric targeted minimum loss-based regression.^41^

### Correlates of protection

#### Controlled vaccine efficacy

For each marker, vaccine efficacy by D35 marker level was estimated by a causal inference approach using baseline risk score-marginalized Cox proportional hazards regression.^42^ A sensitivity analysis of the robustness of results to potential unmeasured confounders of the impact of antibody markers on COVID-19 risk was also conducted. The analysis specified a certain amount of confounding that made it harder to infer a correlate of protection and estimated how much vaccine efficacy increases with quantitative D35 antibody marker despite the specified unmeasured confounder.

#### Hypothesis testing

For hypothesis tests for D35 marker correlates of risk, Westfall-Young multiplicity adjustment^43^ was applied to obtain false-discovery rate adjusted p-values and family-wise error rate (FWER) adjusted p-values. Permutation-based multiple-testing adjustment was performed over the quantitative marker CoR analyses. The SAP specified not conducting hypothesis tests for tertile correlates, due to the small number of breakthrough cases. All p-values were two-sided.

#### Cross-trial comparisons

Calibration of ID50 nAb titers between the Duke University neutralization assay (COVE trial samples) and the Monogram PhenoSense neutralization assay (COV002 and ENSEMBLE trial samples) was performed using the WHO Anti-SARS CoV-2 Immunoglobulin International Standard (20/136) and Approach 1 of Huang et al.^34^ (with arithmetic mean as the calibration factor), as described in the supplementary material of Gilbert, Montefiori, McDermott et al.^25^

#### Software and data quality assurance

The analysis was implemented in R version 4.0.3^40^; code was verified using mock data.

## Data Availability Statement

Data are available from https://clinicaltrials.gov/ct2/show/NCT04611802.

## Code Availability Statement

All analyses were done reproducibly based on publicly available R scripts hosted on the GitHub collaborative programming platform (https://github.com/CoVPN/correlates_reporting2).

## Supporting information

Statistical Analysis Plan

## Data Availability

Data are available from https://clinicaltrials.gov/ct2/show/NCT04611802.

## Acknowledgments

Supported by Novavax; the Office of the Assistant Secretary for Preparedness and Response, Biomedical Advanced Research and Development Authority (BARDA) (contracts Operation Warp Speed: Novavax Project Agreement number 1 under Medical CBRN [Chemical, Biological, Radiological, and Nuclear] Defense Consortium base agreement no. 2020-530, Department of Defense no. W911QY20C0077 and Government Contract No. 75A50122C00008 with Labcorp – Monogram Biosciences); and the National Institute of Allergy and Infectious Diseases (NIAID), National Institutes of Health. The NIAID provides grant funding to the HIV Vaccine Trials Network (HVTN) Leadership and Operations Center (UM1 AI68614), the HVTN Statistics and Data Management Center (UM1 AI68635), the HVTN Laboratory Center (UM1 AI68618), the HIV Prevention Trials Network Leadership and Operations Center (UM1 AI68619), the AIDS Clinical Trials Group Leadership and Operations Center (UM1 AI68636), and the Infectious Diseases Clinical Research Consortium leadership group (UM1 AI148684).

This work was also partially supported by NIAID through award no. R37AI054165 and by the Intramural Research Program of the NIAID Scientific Computing Infrastructure at Fred Hutch, under ORIP grant S10OD028685.

The content is solely the responsibility of the authors and does not necessarily represent the official views of the National Institutes of Health. The findings and conclusions in this report are those of the author(s) and do not necessarily represent the views of the Department of Health and Human Services or its components.

## Author Contributions

Conceptualization: Y.F., D.B., D.F., P.B.G., R.A.K., and R.O.D. Data curation: Y.F., Y.H., D.B., G.A., W.W., A.M.G., L.M.D., I.C., Y.L., C.Y., B.B., L.W.P.v.d.L., N.S.H., and P.B.G. Formal analysis: Y.F., D.B., Y.L., C.Y., B.B., L.W.P.v.d.L., N.S.H., and P.B.G. Funding acquisition: P.B.G., Y.H., R.A.K., and R.O.D. Investigation: G.A., W.W., A.M.G., L.M.D., I.C., K.M., L.J., F.C., C.J.P., A.L., D.H., and B.W. Methodology: Y.F., D.B., L.W.P.v.d.L., N.S.H., D.F., and P.B.G. Project administration: C.R.H., J.A.A., R.O.D., and R.A.K. Resources: C.J.P., C.L.G., K.L.K., A.K.R., M.P.A., J.G.K., J.H., M.K.-J., T.H.B., L.C., K.M.N., R.A.K., and R.O.D. Software: Y.F., D.B., Y.L., C.Y., B.B., L.W.P.v.d.L., and N.S.H. Supervision: Y.F., D.B., D.F., P.B.G., R.A.K., and R.O.D. Validation: Y.F., Y.H., D.B., N.S.H., and P.B.G. Visualization: Y.F., D.B., L.N.C., Y.L., C.Y., B.B., L.W.P.v.d.L., N.S.H., and P.B.G. Writing – original draft: P.B.G. Writing – review and editing: All coauthors.

