## Supplementary Material for "Immune Correlates Analysis of the PREVENT-19 COVID-19 Vaccine Efficacy Clinical Trial"

**Immune Assays Team**

| Affiliation | Team Members |
|---|---|
| Biomedical Advanced Research and Development Authority (BARDA), Washington, DC | Oleg Borisov, Flora Castellino, Brett Chromy, Mark Delvecchio, Ruben O. Donis, Tremel Faison, Corey Hoffman, Christopher Houchens, Tom Hu, Pennie Hylton, Lakshmi Jayashankar, Aparna Kolhekar, James Little, Karen Martins, Jeanne Novak, Azhar Ravji, Carol Sabourin, Evan Sturtevant, Kimberly Taylor, Xiaomi Tong, John Treanor, Danielle Turley, Leah Watson, Daniel Wolfe |
| Boston Consulting Group, Boston, MA | Gian King, Andrew Li, Najaf Shah, Smruthi Suryaprakash, Jue Xiang Wang |
| Division of AIDS, NIAID, NIH, Bethesda, MD | Patricia D'Souza |
| Division of MID (Microbiology and Infectious Diseases), NIAID, NIH, Bethesda, MD | Janie Russell |
| Duke University, Durham, NC | David Beaumont, Kendall Bradley, Jiayu Chen, Xiaoju Daniell, Thomas Denny, Elizabeth Domin, Amanda Eaton, Kelsey Engel, Wenhong Feng, Juanfei Gao, Hongmei Gao, Kelli Greene, Sarah Hiles, Leihua Liu, Kristy Long, Kellen Lund, Charlene McDanal, David C. Montefiori, Marcella Sarzotti-Kelsoe, Francesca Suman, Haili Tang, Jin Tong, Olivia Widman |
| LabCorp-Monogram Biosciences, South San Francisco, CA, USA | Christos J. Petropoulos, Terri Wrin |
| The Tauri Group, an LMI company - Contract Support for U.S. Department of Defense (DOD) Joint Program Executive Office for Chemical, Biological, Radiological and Nuclear Defense (JPEO-CBRND) Joint Project Manager for Chemical, Biological, Radiological, and Nuclear Medical (JPM CBRN Medical), Fort Detrick, Maryland, USA | Christopher S. Badorrek, Gregory E. Rutkowski |
| Vaccine Research Center, NIAID, NIH, Bethesda, MD | Obrimpong Amoa-Awua, Manjula Basappa, Robin Carroll, Britta Flach, Suprabhath Gajjala, Nazaire Jean-Baptiste, Richard A. Koup, Bob C. Lin, Adrian McDermott, Christopher Moore, Mursal Naisan, Muhammed Naqvi, Sandeep Narpala, Sarah O'Connell, Clare Whittaker, Weiwei Wu, Allen Mueller, Martin Apgar, Tommy Bruington, Joe Stashick, Leo Serebryannyy, Mike Castro, Jennifer Wang |

### 2019nCoV-301 Study Group (Pubmed listed, in alphabetical order of institution affiliation)

| Affiliation/Funding* | Study Group | Location |
|---|---|---|
| **México** | | |
| Centro de Atención e Investigación Médica (CAIMED) | Jorge F. Méndez Galván, MD, Monica B. Carrascal, Adriana Sordo Duran, Laura Ruy Sanchez Guerrero, Martha Cecilia Gómora Madrid | Mexico City, Mexico |
| FAICIC Clinical Research | Alejandro Quintín Barrat Hernández, MD, Sharzhaad Molina Guizar, Denisse Alejandra González Estrada, Silvano Omar Martínez Pérez, MD, Zindy Yazmín Zárate Hinojosa, MD | Veracruz, Mexico |
| Instituto Nacional de Ciencias Médicas y Nutrición Salvador Zubirán | Guillermo Miguel Ruiz-Palacios, MD | Mexico City, Mexico |
| Instituto Nacional de Salud Pública | Aurelio Cruz-Valdez, PhD, Janeth, Pacheco-Flores, MD, Anyela Lara, MD, Secia Díaz-Miralrio | Cuernavaca, Mexico |
| PanAmerican Clinical Research México | María José Reyes Fentanes, MD, Jocelyn Zuleica Olmos Vega, MD, Daniela Pineda Méndez, MD, Karina Cano Martínez, MD, Winniberg Stephany Alvarez León | Querétaro, Mexico |
| PanAmerican Clinical Research México | Vida Veronica Ruiz Herrera, MD, Eduardo Gabriel Vázquez Saldaña, Laura Julia Camacho Choza, Karen Sofia Vega Orozco, Sandra Janeth Ortega Domínguez | Guadalajara, Mexico |
| Unidad de Atención Médica e Investigación en Salud (UNAMIS) | Jorge A. Chacón, MD, Juan J. Rivera, MD, Erika A. Cutz, MD, Maricruz E. Ortegón, MD, María I. Rivera, MD | Mérida, Mexico |
| **United States and Puerto Rico** | | |
| Accellacare | David Browder, MD, Cortney Burch, Terri Moye, Paul Bondy, MD, Lesley Browder, MD | Rocky Mount, NC |
| Accellacare | Rickey D. Manning, MD, James Wilson Hurst, MD, Rodney E. Sturgeon, MD, Paul H. Wakefield, MD, John A. Kirby, MD | Knoxville, TN |
| Accel Research Sites | James Andersen, MD, Szheckera Fearon, MSN, FNP-C, Rosa Negron, MD, Amy Medina, ADN, BS | Lakeland, FL |
| Accel Research Sites | Bruce Rankin, DO, John M. Hill, MD, Steven Shinn, MD, Vivek Rajasekhar, DO, Marshall Nash, MD | DeLand, FL |
| Achieve Clinical Research | Hayes Williams, MD, PhD, LaShondra Cade, Rhodna Fouts, Connie Moya | Birmingham, AL |
| Alliance for Multispecialty Research | Corey G. Anderson, MD, Naomi Devine, NP-C, James Ramsey, NP-C, Ashley Perez, David Tatelbaum | Tempe, AZ |
| Alliance for Multispecialty Research | Michael Jacobs, MD, Kathleen Menasche, LPN, Vincent Mirkil, MD | Las Vegas, NV |
| Anaheim Clinical Trials | Peter J. Winkle, MD, Amina Z. Haggag, MD, Michelle Haynes, Marysol Villegas, Sabina Raja | Anaheim, CA |
| Atlanta Center for Medical Research | Robert Riesenberg, MD, Stanford Plavin, MD, Mark Lerman, MD, Leana Woodside, DNP, NP-C, Maria Johnson, MD | Atlanta, GA |
| Baylor College of Medicine / NIAID (UM1AI148575) | C. Mary Healy, MD, Jennifer A. Whitaker, MD, Hana El Sahly, MD, Christine Akamine, MD, Wendy A. Keitel, MD, Robert L. Atmar, MD | Houston, TX |
| Biomedical Advanced Research and Development Authority (BARDA) | Richard Gorman, MD, Gary Horwith, MD, Robin Mason, MS, MBA | Washington, DC |
| Benchmark Research | Laurence Chu, MD, Michelle Chouteau, MD, Lisa Johnson, FNP, Tambra Dora | Austin, TX |
| Benchmark Research | Greg Hachigian, MD, Deborah Murray, FNP, Michael Cancilla, PA, Logan Ledbetter, PA, Masaru Oshita, MD | Sacramento, CA |
| Benchmark Research | William Seger, MD, Beverly Ewing, APRN, DNP, FNP-BC | Fort Worth, TX |
| Beth Israel Deaconess Medical Center / NIAID (UM1AI068614) | Kathryn E. Stephenson, MD, MPH, Chen Sabrina Tan, MD, Rebecca Zash, MD, Jessica L. Ansel, MSN, Kate Jaegle, MSN, Caitlin J. Guiney, MSN | Boston, MA |
| Black Hills Center for American Indian Health / Missouri Breaks Industries Research Inc / NIAID (UM1AI068614) | Jeffrey A. Henderson, MD, MPH, Marcia O'Leary, RN, Kendra Enright, RN, Jill Kessler, MS, Pete Ducheneaux, LPN, Asha Inniss, MS, APRN | Eagle Butte, SD |
| California Research Foundation | Donald M. Brandon, MD, William B. Davis, MD, Daniel T. Lawler, MD | San Diego, CA |
| Carolina Institute for Clinical Research | Yaa D. Oppong, MD, Ryan P. Starr, DO, Scott N. Syndergaard, DO, Rozeli Shelly, MD, Mashrur Islam Majumder | Fayetteville, NC |
| Cedar Crosse Research Center | Danny Sugimoto, MD, Jeffrey Dugas Sr., MD, Dolores Rijos, Sandra Shelton, Stephan Hong, MD | Chicago, IL |
| Cenexel RCA | Howard Schwartz, MD, Nelia Sanchez-Crespo, MD, Jennifer Schwartz, APRN, Terry Piedra, BS, Barbara Corral, APRN | Hollywood, FL |
| Centex Studies | Joel Solis, MD, Carmen Medina, PA, Westley Keating, PA | McAllen, TX |
| Clinical Neuroscience Solutions | Michael E. Dever, MD, Mitul Shah, MD, Michael Delgado, MD, Tameika Scott, DrPH | Orlando, FL |
| Clinical Neuroscience Solutions | Lisa S. Usdan, MD, Lora J. McGill, MD, Valerie K. Arnold, MD, Carolyn Scatamacchia, MSN, NP-C, Codi M. Anthony, DNP, APRN, PMHNP-BC | Memphis, TN |
| CommonSpirit Health Research Institute | Rajan Merchant, MD, Anelgine Crans Yoon, MD, Janet Hill, PA-C, Lucy Ng-Price, MA, Teri Thompson-Seim | Woodland, CA |
| Comprehensive Clinical Research | Ronald Ackerman, MD, Jamie Ackerman, Florida Aristy, APRN | West Palm Beach, FL |
| Covid-19 Prevention Network (CoVPN) | Lawrence Corey, MD, Kathleen M Neuzil, MD, MPH, Huub G Gelderblom, MD, PhD, Nzeera Ketter, Carrie Sopher | Seattle, WA |
| CRA Headlands | Jon Finley, MD, Nathan Segall, MD, Mildred Stull, APRN, FNP-C | Stockbridge, GA |
| DM Clinical Research | Vicki E. Miller, MD, MPH, Monica Murray, Blanca Gomez, Zainab Rizvi, Sonia Guerrero | Tomball, TX |
| Empire Clinical Research | Yogesh K. Paliwal, MD, Amit Paliwal, MD, Sarah Gordon, MS, Bryan Gordon, Cynthia Montano-Pereira | Pomona, CA |
| Headlands Research | Christopher Galloway, MD, Candice Montros, Lily Aleman, Samira Shairi, RN, Wesley Van Ever | Orlando, FL |
| Health Research of Hampton Roads | George H. Freeman, MD, Esther Laverne Harmon, ANP, Marshall A. Cross, MD, Kacie Sales, BSN, RN, Catherine Q. Gular, PharmD | Newport News, VA |
| HHS-DoD Countermeasures Acceleration Group | Matthew Hepburn, MD | Washington, DC |

| | | |
|---|---|---|
| HOPE Research Institute | Matthew Doust, MD, Nathan Alderson, PhD, Shana Harshell | Phoenix, AZ |
| Howard University Hospital / Howard University College of Medicine / NIAID (UM1AI068614) | Siham Mahgoub, MD, Celia Maxwell, MD, Thomas Mellman, MD, Karl M Thompson, PhD, Glenn Wortman, MD | Washington, DC |
| IACT Health | Jeff Kingsley, DO, April Pixler, LaKondria Curry, Sarah Afework, Austin Swanson | Columbus, GA |
| Jacksonville Center for Clinical Research | Jeffry Jacqmein, MD, Maggie Bowers, PA-C, Dawn Robison, APRN-C, Victoria Mosteller, MD, Janet Garvey, DNP | Jacksonville, FL |
| Johnson County Clin-Trials | Carlos Fierro, MD, Mary Easley, BSN, RN | Lenexa, KS |
| Joint Program Executive Office for Chemical, Biological, Radiological and Nuclear Defense's, US Department of Defense | Rebecca J. Kurnat | Washington, DC |
| Lynn Health Science Institute | Carl P. Griffin, MD, Raymond Cornelison, MD, Shanda Gower, APRN, CNP, William Schnitz, MD, Destiny S. Heinzig-Cartwright, BA | Oklahoma City, OK |
| Lynn Institute of the Ozarks | Derek Lewis, MD, Fred E. Newton, MD, Aeiress Duhart, Breana Watkins, Brandy Ball | Little Rock, AR |
| Lynn Institute of the Rockies | Ripley Hollister, MD, Jeremy Brown, DO, Melody Ronk, PA-C, Jill York, Shelby Pickle | Colorado Springs, CO |
| M3-Emerging Medical Research | David B. Musante, MD, William P. Silver, MD, Linda R. Belhorn, MD, Nicholas A. Viens, MD, David Dellaero, MD | Durham, NC |
| M3-Wake Research | Matthew Hong, MD, Wayne Harper, MD, Lisa Cohen, DO, Priti Patel, NP, Kendra Lisec, PA | Raleigh, NC |
| MD Clinical | Beth Safirstein, MD, Luz Zapata, MD, Lazaro Gonzalez, APRN, Evelyn Quevedo, APRN, Farah Irani, PhD | Hallandale Beach, FL |
| Medical Research International | Joseph Grillo, MD, Amy Potts, PA-C, MPH, Julie White, MBA | Oklahoma City, OK |
| Medical University of South Carolina | Patrick Flume, MD, Gary Headden, MD, Brandie Taylor, NP, Ashley Warden, Amy Chamberlain | Charleston, SC |
| MedPharmics | Robert Jeanfreau MD, Susan Jeanfreau MD | Metairie, LA |
| MedPharmics | Paul G. Matherne, MD, Amy Caldwell, RN, Jessica Stahl, Mandy Vowell, Lauren Newhouse | Gulfport, MS |
| Meharry Medical College / NIAID (UM1AI068614) | Vladimir Berthaud MD, MPH, Zudi-Mwak Takizala MD, MPH, MBA, Genevieve Beninati, FNP, Kimberly Snell, PharmD, Sherrie Baker, BS, James Walker, RN | Nashville, TN |
| Meridian Clinical Research | David Ensz, MD, Tavane Harrison, CNP, Meagan Miller, Janet Otto | Sioux City, IA |
| Meridian Clinical Research | Brandon Essink, MD, Roni Gray, APRN, Christine Wilson, Tiffany Nemecek, Hannah Harrington, MPH | Omaha, NE |
| Meridian Clinical Research | Charles Harper, MD, Keith Vrbicky, MD, Chelsie Nutsch, NP, Sally Eppenbach, NP, Wendell Lewis, NP | Norfolk, NE |
| Meridian Clinical Research | Jordan Whatley, MD, Christopher Dedon, APRN, FNP-C, Tana Bourgeois, RN, Lyndsea Folsom, Crystal Rowell, APRN, FNP-C | Baton Rouge, LA |
| Miami Veterans Affairs Medical Center / NIAID (UM1AI068614) | Gregory Holt, MD, Mehdi Mirsaeidi, MD, Rafael Calderon, MD, Paola Lichtenberger, MD, Jalima Quintero, RN, Becky Martinez, RN | Miami, FL |
| Morehouse School of Medicine / NIAID (UM1AI068614) | Lilly Immergluck, MD, Erica Johnson, PhD, Austin Chan, MD, Norberto Fas, MD, LaTeshia Thomas-Seaton, MS, APRN, Saadia Khizer, MD, MPH | Atlanta, GA |
| MultiCare Institute for Research and Innovation | Jonathan Staben, MD | Cheney, WA |
| National Institute of Allergy and Infectious Diseases (NIAID) / National Institutes of Health (NIH) | Tatiana Beresnev, MD, Maryam Jahromi, MD, Mary A. Marovich, MD, Julia Hutter, MD, Martha Nason, PhD, Julie Ledgerwood, DO, John Mascola, MD | Bethesda, MD |
| National Research Institute | Mark Leibowitz, MD, Fernanda Morales, Mike Delgado, Rosario Sanchez, Norma Vega | Los Angeles, CA |
| Novavax, Inc. | Lisa M. Dunkle, MD, Germán Áñez, MD, Gary Albert, Erin Coston, Chinar Desai, Haoua Dunbar, Mark Eickhoff, Jenina Garcia, Margaret Kautz, Angela Lee, Maggie Lewis, Alice McGarry, Irene McKnight, Joy Nelson, Patrick Newingham, Patty Price-Abbott, Patty Reed, Diana Vegas, Bethanie Wilkinson, PhD, Katherine Smith, MD, Wayne Woo, MS, Iksung Cho, MS, Gregory M. Glenn, MD, Filip Dubovsky, MD, MPH | Gaithersburg, MD |
| Omega Medical Research | David L. Fried, MD, Lynne A. Haughey, MSN, FNP, Ariana C. Stanton, PA-C, Lisa Stevens Rameaka, MD | Warwick, RI |
| Pharmacology Research Institute | David Rosenberg, MD, Lee Tomatsu, Viviana Gonzalez, Millie Manalo | Los Alamitos, CA |
| PMG Research of Bristol | Bernard Grunstra, MD, Donald Quinn, MD, Phillip Claybrook, MD, Shelby Olds, MD, Amy Dye | Bristol, TN |
| PMG Research of Wilmington | Kevin D. Cannon, MD, Mesha M. Chadwick, MD, Bailey Jordan, Morgan Hussey, Hannah Nevarez | Wilmington, NC |
| Ponce de Leon Center / NIAID (UM1AI068614) | Colleen F. Kelley, MD, MPH, Valeria D. Cantos MD, Michael Chung MD, Caitlin Moran, MD, MSc, Paulina Rebolledo, MD, Christina Bacher, PAC | Atlanta, GA |
| Ponce School of Medicine / NIAID (UM1AI148685) | Elizabeth Barranco-Santana, MD, Jessica Rodriguez, MD, Rafael Mendoza, MD, Karen Ruperto, MD, Odette Olivieri, MD, Enrique Ocaña, MD | Ponce, Puerto Rico |
| Preferred Research Partners | Paul E. Wylie, MD, Renea Henderson, DO, Natasa Jenson, MD, Fan Yang, MD, Amy Kelley, BSN, RN | Little Rock, AR |
| Providea Health Partners Elligo Health Research | Kenneth Finkelstein, DO, David Beckmann, MD, Tanya Hutchins, FNP, Sebastian Garcia Escallon, BA, Kristen Johnson | Evergreen Park, IL |
| Providence Clinical Research | Teresa S. Sligh, MD, Parul Desai, NP, Vincent Huynh, BSc, Carlos Lopez, MD, Erika Mendoza, BA | North Hollywood, CA |
| Research Your Health | Jeffrey Adelglass, MD, Jerome (Jerry) G. Naifeh, MD, Kristine Jane Kucera, PA-C, MPAS, DHS, Waseem Chughtai, BS, MBBS, Shireen Hasham Jaffer | Plano, TX |
| Rochester Clinical Research | Matthew G. Davis, MD, Jennifer Foley, Michelle Lyn Burgett, RN, Tammi Louise Shlotzhauer, MD, Sarah Michelle Ingalsbe-Geno, RPA-C | Rochester, NY |
| SIMEDHealth / SIMEDResearch | Daniel Duncanson, MD, Kelly Kush, Lori Nesbitt, Cora Sonnier, Jennifer McCarter | Gainesville, FL |

| | | |
|---|---|---|
| Sterling Research Group | Michael B. Butcher, MD, James Fry, PA-C, Donna Percy, RN, BSN, Karen Freudemann | Cincinnati, OH |
| Sterling Research Group | Bruce C. Gebhardt, MD, Padma N. Mangu, MD, Debra Beck Schroeck, MS, PA-C, Rajesh Kumar Davit, MD, Gayle D. Hennekes, PA-C, MPAS | Cincinnati, OH |
| Stony Brook University - Stony Brook Medicine / NIAID (UM1AI068614) | Benjamin J. Luft, MD, Melissa Carr, BA, Sharon Nachman, MD, Alison Pellecchia, BA, Candace Smith, PharmD, Bruno Valenti, NP | Commack, NY |
| Suncoast Research Associates | Maria I. Bermudez, MD, Noris Peraita, ARNP, Ernesto Delgado, ARNP, Alicia Arrazcaeta, Natalie Ramirez | Miami, FL |
| Suncoast Research Group | Mark E. Kutner, MD, Jorge Caso, MD, Janet Mendez, ARNP, Marianela Carvajal, ARNP, Carmen Amador, ARNP | Miami, FL |
| Sundance Clinical Research | Larkin Tyler Wadsworth III, MD, Horacio Marafioti, MD, Lyly Dang, DNP-BC, Lauren Clement, NP-C, Jennifer Berry, FNP-BC | St. Louis, MO |
| Synexus Clinical Research | Mohammed Allaw, MD, Georgettea Geuss, Chelsea Miles, NP, Zachary Bittner, Melody Werne | Evansville, IN |
| Synexus Clinical Research | Cornell Calinescu, MD, Shannon Rodman, Joshua Rindt | Henderson, NV |
| Synexus Clinical Research | Erin Cooksey, MD, Kristina Harrison, Deanna Cooper, Manisha Horton Amanda Philyaw | Anderson, SC |
| Synexus Clinical Research | William Jennings, MD, Hilario Alvarado, MD, Michele Baka, MD, Malina Regalado, NP | San Antonio, TX |
| Synexus Clinical Research | Linda Murray, DO | Pinellas Park, FL |
| Synexus Clinical Research | Sherif Naguib, MD, Justin Singletary, Sha-Wanda Richmond, Sarah Omodele, Emily Oppenheim | Atlanta, GA |
| Synexus Clinical Research | Joseph Newberg, MD, Laura Pearlman, MD, Reuben Martinez, Victoria Andriulis | Chicago, IL |
| Synexus Clinical Research | Paul J. Nugent, DO, Leonard Singer, MD, Jeanne Blevins, Meagan Thomas, Christine Hull | Cincinnati, OH |
| Synexus Clinical Research | Isabel Pereira, MD, Gina Rivero, Tracy Okonya, Frances Downing, Paulina Miller | Vista, CA |
| Synexus Clinical Research | Margaret Rhee, MD, Katherine Stapleton, Jeffrey Klein, Rosamond Hong, MD | Akron, OH |
| Synexus Clinical Research | Suzanne Swan, MD, Tami Wahlin, MD, Elizabeth Bennett, PA, Amy Salzl Sharine Phan | Richfield, MN |
| Synexus Clinical Research | Jewel Johnny White, MD, Amanda Occhino, Ruth Paiano APRN, Morgan McLaughlin APRN, Elisa Swieboda APRN | The Villages, FL |
| Texas Center for Drug Development | Veronica Garcia-Fragoso, MD, Maria Gabriela Becerra, MD, Cecilia Mckeown, Lisa Holloway, Toni White | Houston, TX |
| The Charlotte-Mecklenburg Hospital Authority d/b/a Atrium Health / NIAID (UM1AI068614) | Christine B. Turley, MD, Andrew McWilliams, MD, Tiffany Esinhart, PA-C, Natasha Montoya, APRN, Shamika Huskey, FNP, Leena Paul, FNP | Charlotte, NC |
| The Miriam Hospital / NIAID (UM1AI068636) | Karen Tashima, MD, Jennie Johnson, MD, Marguerite Neill, MD, Martha Sanchez, MD, Natasha Rybak, MD, Maria Mileno, MD | Providence, RI |
| UC Davis Health / NIAID (UM1AI068614) | Stuart H. Cohen, MD, Monica Ruiz, Dean M. Boswell, BS, Elizabeth E. Robison, BS, Trina L. Reynolds, BS, Sonja Neumeister, MPH | Sacramento, CA |
| Universidad de Puerto Rico - Recinto de Ciencias Médicas - Maternal Infant Studies Center (CEMI) / NIAID (UM1AI068636) | Carmen D. Zorrilla, MD, Juana Rivera, MD, MPH, Jessica Ibarra, MD, Iris García, BSN, RN, Dianca Sierra, BA, Wanda Ramon, BSPh | San Juan, Puerto Rico |
| University of Colorado Hospital CRS / NIAID (UM1AI068636) / NCATS (UL1TR002535, UM1AI069432) | Thomas B. Campbell, MD, Suzanne Fiorillo, MSPH, Rebecca Pitotti, RNP, Victoria Riedel Anderson, MS, Jose Castillo Mancilla, MD, Nga Le, PharmD | Aurora, CO |
| University of Iowa Medical Center / NIAID (UM1AI068614) / NCATS (UL1TR002537) | Patricia L. Winokur, MD, Dilek Ince, MD, Theresa Hegmann, PA, Jeffrey Meier, MD, Jack Stapleton, MD, Laura Stulken, PA | Iowa City, IA |
| University of Maryland School of Medicine / NIAID (UM1AI148689) | Monica McArthur, MD, PhD, Karen L. Kotloff, MD, Kathleen Neuzil, MD, Andrea Berry, MD, Milagritos Tapia, MD, Elizabeth Hammershaimb, MD, MS, Toni Robinson, RN, Rosa MacBryde, RN | Baltimore, MD |
| University of Minnesota / NIAID (UM1AI068614) | Susan Kline, MD, MPH, Joanne L. Billings, MD, MPH, Winston Cavert, MD, Les B. Forgosh. MD, Timothy W. Schacker, MD, Tyler D. Bold, MD, PhD | Minneapolis, MN |
| University of Missouri Health Care / NIAID (UM1AI148685) | Dima Dandachi, MD, MPH, Taylor Nelson, DO, Andres Bran, MD, Grant Geiger, S. Hasan Naqvi, MD | Columbia, MO |
| University of Nebraska Medical Center / NIAID (UM1AI068614) | Diana F Florescu, MD, Richard Starlin, MD, David Kline, MD, Andrea Zimmer, MD, Anum Abbas, MD, Natasha Wilson, APRN | Omaha, NE |
| University of North Carolina / NIAID (UM1AI068619) / University of North Carolina at Chapel Hill Center for AIDS Research (P30AI050410) / NC TraCS Institute (UL1TR002489) | Cynthia L. Gay, MD, MPH, Joseph J Eron, MD, Michael Sciaudone, MD, MPH, A. Lina Rosengren, MD, MPH, MS, John S Kizer, MD, Sarah E Rutstein, MD, PhD | Chapel Hill, NC |
| University of South Florida, Morsani College of Medine / NIAID (UM1AI068614) | Carina A. Rodriguez, MD, Elizabeth Bruce, MD, Claudia Espinosa, MD, Lisa J Sanders, MD, Kami Kim, MD, Denise Casey, RN | Tampa, FL |
| University of Texas Health Science Center San Antonio / NIAID (UM1AI068614) | Barbara S. Taylor, MD, MS, Thomas Patterson, MD, Ruth Serrano Pinilla, MD, Delia Bullock, MD, Philip Ponce, MD, Jan Patterson, MD | San Antonio, TX |
| University of Washington / Lummi Tribal Health Center / NIAID (UM1AI148573) | R. Scott McClelland, MD, MPH, Dakotah C. Lane, MD, Anna Wald, MD, MPH, Frank James, MD, Elizabeth Duke, MD, Kirsten Hauge, MPH, Jessica Heimonen, MPH | Seattle, WA |
| University of Washington | Robert W. Coombs, MD, PhD, Alex Greninger, MD, PhD, MS, MPhil, Pavitra Roychoudhury, PhD, Erin A. Goecker, MS, Yunda Huang, PhD, Youyi Fong, PhD | Seattle, WA |
| VA Ann Arbor Healthcare System / NIAID (UM1AI068614) | Carol Kauffman, MD, Kathleen Linder, MD, Kimberly Nofz, BSN, Andrew McConnell, BS | Ann Arbor, MI |

| | | |
|---|---|---|
| Velocity Clinical Research | Robert J. Buynak, MD, Angella Webb, APRN, Taryn Petty, FNP, Stephanie Andree, FNP | Valparaiso, IN |
| Velocity Clinical Research | Judith Kirstein, MD, Marcia Bernard, Erica Sanchez, Nolan Mackey, Clarisse Baudelaire | Banning, CA |
| Velocity Clinical Research | Gregg Lucksinger, MD, Jaleh Ostovar, NP | Medford, OR |
| Velocity Clinical Research | Mary Beth Manning, MD, Joan Rothenberg, MD, Toby Briskin, MD, Denise Roadman, PAC, Sarah Dzigiel | Cleveland, OH |
| Velocity Clinical Research | J. Scott Overcash, MD, Adrienna Marquez, Hanh Chu, Kia Lee, Kim Quillin | La Mesa, CA |
| Velocity Clinical Research | Barbara Rizzardi, MD, Michelle King, NP, Vanessa Abad, NP, Jennifer Knowles, BS | West Jordan, UT |
| Velocity Clinical Research | Michael Waters, MD, Karla Zepeda, NP, Scott Overcash, MD, Jordan Coslet, NP, Dalia Tovar, MA | Chula Vista, CA |
| Velocity Clinical Research | Marian E. Shaw, MD, Mark A. Turner, MD, Cory J. Huffine, FNP-C, Esther S. Huffine, FNP-C | Meridian, ID |
| Walter Reed Army Institute of Research | Julie A. Ake, MD, MSc | Silver Spring, MD |
| Wayne State University / NIAID (UM1AI068614) | Elizabeth Secord, MD, Eric McGrath, MD, Phillip Levy, MD, Brittany Stewart, RD, PharmD, Charnell Cromer, RN, MSN, Ayanna Walters, RN, BSN | Detroit, MI |
| Weill Cornell Chelsea CRS / NIAID (UM1AI068619) | Kristen Marks, MS, MD, Grant Ellsworth, MD, MS, Caroline Greene, ANP-BC, Sarah Galloway, BA, Shashi Kapadia, MD, MS, Elliot DeHaan, MD | New York, NY |
| Willis-Knighton Health System / WKB Family Medicine Associates | Clint Wilson, MD, Jason Milligan, MD, Danielle Raley, MD, Joseph Bocchini, MD | Bossier City, LA |
| Womack Army Medical Center | Bruce McClenathan, MD, Mary Hussain, BS, Evelyn Lomasney, MD, Evelyn Hall, MMS, PA-C, Sherry Lamberth, PharmD | Fort Bragg, NC |
| WR Clinsearch | Mark McKenzie, MD, Teresa Deese, Christy Schmeck, Vickie Leathers, Christy Sweet | Chattanooga, TN |

* Funding of institutions by the National Institute of Allergy and Infectious Diseases (NIAID) and/or research support by the National Center for Advancing Translational Science (NCATS), as indicated. All other institutions were funded by Office of the Assistant Secretary for Preparedness and Response, Biomedical Advanced Research and Development Authority. The content of this publication is solely the responsibility of the authors and does not necessarily represent the official views of the funding sources.

### 2019nCoV-301 Principal Investigators and Study Team (in alphabetical order)

| Principal Investigator | Study Team | Institution | Location |
|---|---|---|---|
| Ronald Ackerman, MD | Jamie Ackerman, Florida Aristy, Tomeko Heard, Diana Mann, Maureen Stewart, Cheryl Demczyk, Rohan Barron, Ashley Torres, Jennifer Gomez, Tiffany Potter | Comprehensive Clinical Research | West Palm Beach, FL |
| Jeffrey Adelglass, MD | Jerome (Jerry) G. Naifeh, Kristine Jane Kucera, Waseem Chughtai, Shireen Hasham Jaffer, Anuja Sathe, Cameron Galownia, Cheryl Hill, Ramiro Lopez, Erica Parker-Martinez, Helene Harrison, Chiedza Mutindori, Sabrina Flowers, Tamara Betters, Carolyn Ackley, Pamela Fox, Noelia Tejada James, Dorothy Saylor, Hallen Dao, Jon Etta Randolph, Jason Tentativa, Malaika Chughtai, Shanzae Chughtai, Maheen Shah, Hayyan Chughtai, Tyler Love, Ti'arah Love | Research Your Health | Plano, TX |
| Mohammed Allaw, MD | Georgettea Geuss, Chelsea Miles, Zachary Bittner, Melody Werne, Lyndsey Morrison, Stephanie Albin, Linda Frazier, Jacque Nalley, Christie Borin, Jacque Nalley | Synexus Clinical Research | Evansville, IN |
| James Andersen, MD | Szheckera Fearon, Rosa Negron, Amy Medina, Diana Holmes, Colleen Figueroa, Cristal Ruiz, Nancy Masseus Tare Floyd, Kenta Oliver, Candice Gerber, Mae Ann Francisco, Gilbert de la Cruz, Ginny McClanahan, Veronica Walker, David Irwin, Gloria Adejobi | Accel Research Sites | Lakeland, FL |
| Corey G. Anderson, MD | Naomi Devine, James Ramsey, Tyanna Montijo, Ashley Perez, David Tatelbaum, Lisa M. Dean, Angela D. Ledezma, Anthony Padilla, Cecilia M. Tanori, Georgina Lopez-Wood, Tasha C. Marriott, Ronald Hawkins, Hannah Spinks | Alliance for Multispecialty Research | Tempe, AZ |
| Elizabeth Barranco-Santana, MD | Michele Irizarry, Alice Grace Rodriguez, Irmaris Arroyo, Sara Cancel, Alejandra Román, Juan D. Lugo, Armando X. Torres, Marianne Hernandez, Brenda Garcia, Nancy Jiménez, Orlando Torres | Ponce School of Medicine | Ponce, Puerto Rico |
| Alejandro Quintín Barrat Hernández, MD | Sharzhaad Molina Guizar, Denisse Alejandra González Estrada, Silvano Omar Martínez Pérez, Zindy Yazmín Zárate Hinojosa, Norberto Daniel Vázquez Tinajero, Yessica Olivo Domínguez, Daniel Hernández León, Gloria Norma Ambrosio Lara, José Carlos Mateos Castro, Irving Nerí Leyva Ferrer, María Fernanda Hernández García, Heidy Jazmín Maldonado Pavón, Evelyn Monserrat Bravo Serralta, Edgar Iván Muñoz López, Karina Esmeralda García Mateo, Lorena Cruz Cruz, José Javier Zárate Hinojosa, Javier Torres Cole, Yareth Jiménez Barcenas, Andrea Anaid Rangel Huerta, Erika Guillén González, María de la Luz Rufina Martínez Lugo, Angélica Liliana Muñoz Solano, David Sena Gómez, Berenice Valera Montalvo, Moisés Miguel Ruíz Nogueira, Yoshira Montero Díaz, Francisco Javier Martínez Osorio, Alejandra Morales Arias, Sandra Itzel Solis Rivera, Alejandro Esteban Cortina, Aldo Miguel López Domínguez, María Fernanda Cortés Ruíz, Marilyn Yulissa Ramírez Domínguez, Lucero Moctezuma Juan, Francisco Barrales Arcos | FAICIC Clinical Research | Veracruz, Mexico |
| Maria I. Bermudez, MD | Noris Peraita, Ernesto Delgado, Alicia Arrazcaeta, Natalie Ramirez, Giovanna Salcedo, Aliana Amador, Elizabeth Martinez, Arleen Aspuru, Gabriella Gonzalez, Gabriella Alabaci, Livan Sanchez, Raul Tejeda, Adriana Bello, Barbara Vega-Aguera, Kassandra Martinez, Grettel Obregon, Oscar Alejandro Gutierrez Luna, Magela C. Dominguez, Lauren Pena | Suncoast Research Associates | Miami, FL |
| Vladimir Berthaud, MD, MPH | Toni Hall, Livette Johnson, Sylvia Eluhu, Ana Tomescu, Katharina Whitbeck, Rajbir Singh | Meharry Medical College | Nashville, TN |
| Donald M. Brandon, MD | William B. Davis, Daniel T. Lawler, Maria Aceves, Kathleen B. Anderson, Hana Berry, Janice E. Brandon, Jeffrey C. Brandon, Patricia A. Brandon, Lorraine Boggs, Charlene Cruz, Mairead Hawkins, Clarice Hranicky, Andrew J. McCrea, Karen G. McCrea, Kimberly Najera, Tierney J. O'Connor, Michelle L. Rios, Cindy F. Stevens, Hannah J. Zapata | California Research Foundation | San Diego, CA |
| David Browder, MD | Cortney Burch, Terri Moye, Michael Wright, Paul Bondy, Lesley Browder | Accellacare | Rocky Mount, NC |
| Michael B. Butcher, MD | James Fry, Julia Froschauer, Allison Deuel, Jeanne Piccola, Donna Percy, Karen Freudemann, Lois Rawe, Megan Bryant, Kurt Percy, Jon Marvin, Luann Corcoran | Sterling Research Group | Cincinnati, OH |
| Robert J. Buynak, MD | Mark Yarosz, Rachel McNeal, Megan Smith, Patricia Volom, Nicholas Hanna, Erica Lewis, Miranda Lee, Goldie Luna, Marilyn Idowu, Destiny Williams, Jessica Johnson, Consuelita Perez, Priscilla Dodson | Velocity Clinical Research | Valparaiso, IN |
| Cornell Calinescu, MD | Shannon Rodman, Joshua Rindt, Krystal Tyner, Lovelyn Vincente, Alejandro Osuna-Meda, Charmaine Brown, Matthew Derrick, Melodee Morrison, Marissa Washington | Synexus | Henderson, NV |
| Thomas B. Campbell, MD | Donna McGregor, Laurel Ware, Myron Levin, Steven Johnson, Sophia Quesada, Martin Krsak, Kristine Erlandson, Nicholas Sarchet, Vanessa Sutton, Lawrence Moran, Tracey Stevenson, Alaina Dougherty, Julianne Randlemon | University of Colorado Hospital CRS | Aurora, CO |
| Kevin D. Cannon, MD | Mesha M. Chadwick, Bailey Jordan, Taylor Fedorcha, Kathryn Zweier, Brettany Holt, Emily Johnson, Karen Ruggiero, Olivia Houghton, Courtney Christie, Allison Dunn, Courtney Boyce, Sasha Saint-Lot, Ashley Andrades, Ashley Miller, LaShaya Dunston, Russell Larkins, Brittany Savoca, Hannah Nevarez, Hannah Nevarez, Larkin Collins, Morgan Cyrus, Morgan Hussey, Christina MacNaughton, Heidi Kaufman, Sheila Gard, Alyssa Gaylor, Bethany Donelan-Wilson, Taylor Bayless, Anna McManus, Tracie Marlowe Bryant, Ben Manuel, Laura McMillan, Nicole Stigers, Prerana Zanke | PMG Research of Wilmington | Wilmington, NC |
| Jorge A. Chacon, MD | Juan J. Rivera, Erika A. Cutz, Maricruz E. Ortegon, María I. Rivera, Ricardo Cervera, Felipe Rivera, Daniela Pat, Daniela Cruz, Alberto Chacon, Kattia Borges, Aldo Borraz, Rebeca Ortegon, Karla Ic, Carmen Ojeda, Irvin Ortega, Mayra Jimenez, Cindy Novelo, Pharmacist, Mónica Pérez, Adriana Hernandez, Laura Martinez | Unidad de Atención Medica e Investigación en Salud (UNAMIS) | Merida, Mexico |
| Laurence Chu, MD | Michelle Chouteau, Lisa Johnson, Tambra Dora, Lamar Box, Michelle Listz, Katherine Davis, Jennifer Montes, Jessica Ruff, Jennifer Leyva, Pamela Fidler, Ruth Fitch, Sean Turnbow, Francesca Vigil, Maria Barrientes, Isaiah Knight, Cindy Duran, Lauren Christal, Breana Wade Liaison, Brooke Harris, Dean Skiles, Marisol Ramos, Brandon Newsom, Candace Gaitan, David Pereira | Benchmark Research | Austin, TX |

| | | | |
|---|---|---|---|
| Stuart H. Cohen, MD | Curtis Blankenship, Katelyn Trigg, Courtney Lymuel, Gursimran Mann, Zayan Musa, Hana Minsky, Eliseo Vasquez, Nicole Garza, Kaitlyn Low, Mehrab Hussain, William Li, Rahul Araza, Monique Conover, George Thompson, Hien Nguyen, Scott Crabtree, Bennett Penn, Minh-Vu Nguyen, Archana Reddy, Derek Bays, Kaitlyn Hardin, Matthew Boutros, Alan Koff, Natascha Tuznik, Angel Desai, Naomi Hauser, Sarah Waldman, Gauri Barlingay, Dean Blumberg | UC Davis Health | Sacramento, CA |
| Erin Cooksey, MD | Kristina Harrison, Deanna Cooper, Manisha Horton, Amanda Philyaw | Synexus Clinical Research | Anderson, SC |
| Aurelio Cruz-Valdez, PhD | Janeth, Pacheco-Flores, Anyela Lara, Secia Diaz-Miralrio | Instituto Nacional de Salud Pública | Cuernavaca, México |
| Dima Dandachi, MD, MPH | Tami Day, Britlyn Brown, Taylor Mathews | University of Missouri Health Care | Columbia, MO |
| Matthew G. Davis, MD | Therese Dayton, Joseph I. Mann, Patricia S. Larrabee, Jean C. Kelly, Tia. L Albro, Zerina Zornic, Susan J. Willer, Donna M. Willome, Kathleen K. Ebeling, Jaclyn P. Zona, Julie A. Mooney, Katherine A. Pagenkemper, Victoria F. Fink, Christine N. Hall, Chelsea Bork, Abigail Miller, Mackay Kanaley, Chelsey LoMonaco, Marie Musolino, Jessica Fisher, Katilyn Bergen, Rachel Bordonaro, Cassidy Glod, Liam Sullivan, Brandi Douglass, Ann Casey, Philip LaSpino, Maurice Holmes | Rochester Clinical Research | Rochester, NY |
| Michael E. Dever, MD | Michael Delgado, Tameika Scott, Laverne Denise Davila, Nelisa Frias, Anissa Hilton, Patricia Brown, Shana Caldwell, Martha Hendrix, Edmund Delgado, Mitul Shah, Gracemarie Rosario, Kaneitra Williamson, Taylor Lucier, Jaime Hawat, Matthew Stephens, Monica Cooper, Dante Canidate, Denise Pagan, Sierra Robinson, Pascal Nelson-Quiles, Anthony Perez, Chanel Adams, Keisha Foster, Scott Salmon, Andrew Lockwood, Priya Moorhouse, Paul Yi | Clinical Neuroscience Solutions | Orlando, FL |
| Matthew Doust, MD | Stephanie Catanzaro, Shana Harshell, Madison Mikulak, Bettie, D'Nise Corcoran, Susan DeCraene, Jasmin Redden, Brian DeCraene, Karen Wakefield, Adrian Aljeo, Denise Sample, Clarissa Lara, Stephanie Junker, Nathan Alderson, Kimberly Joshlin, Mia Munoz, Michele Aguirre, Dina Reyes Cordova, Neil Pearson | HOPE Research Institute | Phoenix, AZ |
| Daniel Duncanson, MD | Kelly Kush, Lori Nesbitt, Cora Sonnier, Jennifer McCarter, Thomas Buschbacher, Evie Zavala, Brittany Cooper, Abbey Mannings, Melissa Berrio, Erin Juhl, William Douglas, Timothy Elder, Linda Grover, Colleen Crabbe, Rachel Francis, Jesse Lipnick, Seldon Longley, Michael Rozboril, Madison Duncanson, Jakob Vaes, Michael Costa, Dhruv Panchal, Michelle Hendricks, Sergio Montalvo, Angel Dubois | SIMEDHealth / SIMEDResearch | Gainesville, FL |
| David Ensz, MD | Bruce Rankin, Tavane Harrison, Meagan Miller, Kayla Sturgeon, Jessica Knight, Janet Otto, Monica Salazar, Megan Howard, Carly Deges, Joseph Harris, Rylea Gulick, Melissa Wiseman, Sue Doty | Meridian Clinical Research | Sioux City, IA |
| Brandon Essink, MD | Roni Gray, Christine Wilson, Fritz Raiser, Akossiwa "Essi" Yovogan, Jessica Satorie Tiffany Nemecek, Hannah Harrington, Amy Lett-Brown,Chelsea Steinmetz, Tabitha Campbell, Carrie Essink, Jamie Meyer, Riley Brockman, Melissa Monarrez, Troy Humphries, Wynter Huffman, Brooke Dworak, Raquel Davis, Samantha Nocita, Heidi Smith, Carissa Schejbal, Kayla Flege, Joe Genoways, Jessa Swanson, Avery Dunn, Kevin Grimes, Phillip Astorino, Ashtynn Jarosz, Hailey Harper, Amy Nichols, Azra Bauman, Jessica Fellows, Courtney Heisey, Ginny McNew | Meridian Clinical Research | Omaha, NE |
| Carlos Fierro, MD | Natalia Leistner, Amy Thompson, Celia Gonzalez, Nathan Arthur, Mazen Zari, Mary Easley, Heather Barker, Manyvohn Rinehart, Monica Atwood, Natalya Amrine, Kelly Moen, Kaley Miller, Angela Eichler, Ann Geier, Christa Estrada, Amber Wolf, Denise Essix, Latoria Rios, Kasie Hickert, Kenny Nguyen, Karol Moore, Stefanie Uwah, Kaelyn Howell, Miranda Dean | Johnson County Clin-Trials | Lenexa, KS |
| Kenneth Finkelstein, DO | David Beckmann, Tanya Hutchins, Sebastian Garcia Escallon, Kristen Johnson, Athena Rivera, David Otuada, Jessica Bartlett, Lauren Wade, Tyler Will, Gina Nielsen-Grewe, Anita Suri | Providea Health Partners Elligo Health Research | Evergreen Park, IL |
| Jon Finley, MD | Nathan Segall, Mildred Stull, Michelle Sowell, Michelle Binns, Kiara Tyner, Karen Yangapatty, Elizabeth West, Cynthia Steele, Kwannda Whatley, Hannah Smith, Pamela Talbott, Kimberly Cobb, Donna Toepfer, Jennifer LeBrun, Susan Jones, Patrizia Greene, Cynthia Pinckney, Kim Banaski, Karen Hickson | CRA Headlands | Stockbridge, GA |
| Diana F Florescu, MD | Mark Rupp, Daniel Brailita, Adia Sikyta, Erica Stohs, Sara Hurtado Bares, Nada Fadul, Matthew Lunning, Elizabeth Schnaubelt, Molly Ferris, Andrew Buettner, Matthew Palmer, Bailee Lichter, Alison Lewis, Chase Kimberling, Jonathan Beck, Erin Iselin, Kimmai McClain, Andrew Schnaubelt | University of Nebraska Medical Center | Omaha, NE |
| Patrick Flume, MD | Gary Headden, Brandie Taylor, Ashley Warden, Amy Chamberlain, Kim Spencer, April Rasberry, Angela Millare, Angel Darrow, Abbey Grady, Max Lento, Allison Patterson, Caitlan LeMatty, Jhonatan Diaz, Andrew Stephens, Emalee Wood, Destri Eichman, Annie Cribb, Annelise Kauffman<br>Charnele Handy, Elizabeth Poindexter, Moira Chance, Anna Miller, Elizabeth Dickinson, Andrea Boan, Erin Klintworth | Medical University of South Carolina | Charleston, SC |
| Veronica Garcia-Fragoso, MD | Maria Gabriela Becerra, Cecilia Mckeown, Lisa Holloway, Toni White, Bonnie Colville, Frederic Santiago, Teresa Becker, Shakira Barr, Chen Ho Yang, Tracy Kowalski, Danitra Glasper, Diana Chehab Nazanin Zarinkamar, Joanna Quezon, Maryam Rabbani, Sadaf Batla, Ayla Perez, Berenice Ferrero, Dean Jang, Biman Goswami, Dustin McFadden, Elton Oliveira, Enya Rentas-Sherman, Julian Edmonson, Laura Plaza-Grisanty, Olga Konshina, Rachely Araujo-Gutierrez, Scott Ward, Teodoro Seminario, Patricia Matute, Sauleha Husain, Akram Assaf, Elisa Moralez, Frances Saubon, Jenny Torres, William Fernandez, Ashraf Jafri, Amy Anderson, Saji Mathew Perinjelil, Waheeda Sureshbabu, Kara Sikes, Joel Cano, Kendra Rogers, Quiana Wilson, Karina Sainz, Abdeali Dalal, Leena Mir, Misbah Baloch, Shammarrian Hampton, Crystal Reese, Lucia Almaguer,<br>Felicia Ardoin, Deep Patel, Bernardo Martinez Leal, Faryal Mahmood, Ana Rueda, Norma Gonzalez, Stacey Montero, Chandra Tobin, Abyssinia Moges, Ari Amirkhosravi, Herman | Texas Center for Drug Development | Houston, TX |

| | | | |
|---|---|---|---|
| | Ortiz, Matthew Joseph, Parul Mehta, Zain Rizvi, Diego Carrington, Blessing Feliz-Okoroji, Moez Talpur, Robert Krbashyan, Simeen Khan, Mary Rogers | | |
| George H. Freeman, MD | Esther Laverne Harmon, Marshall A. Cross, Kacie Sales, Catherine Q. Gular, Amanda Fronzaglio, Timothy O'Malley, Zaahin Huq, Jenna Johnson, Jessica Fuggett, Danielle Merian, Rita Quinn | Health Research of Hampton Roads | Newport News, VA |
| David L. Fried, MD | Lynne A. Haughey, Ariana C. Stanton, Lisa Stevens Rameaka | Omega Medical Research | Warwick, RI |
| Christopher Galloway, MD | Candice Montros, Lily Aleman, Samira Shairi, Robert Duran, Wesley Van Ever, Wasilah Suid, Sandra Torres, Taylor Rice, Wanda Estrada, Julie Castillo,Stephanie Cassidy, Ashleigh Ford, Thai Marie, Colon Maldonado, Amedaris Cordero, Zahra Somji, Rachel Morris | Headlands Research | Orlando, FL |
| Cynthia L. Gay, MD, MPH | David Wohl, Michelle Floris-Moore, Michael Herce, Danielle Clement, Arianna Morrison, Jan Busby-Whitehead, Michelle Hernandez, Zachary Willis, Allison, Burbank, Peyton Thompson, Chris Evans, Susan Pedersen, Becky Straub, Samantha Earnhardt, Erin Hoffman, Jonathan Oakes, Tevnan Keller, Victoria Rucinski, Camille O'Reilly, Kelsey Vollmer, Jennifer Rees, April Welch, Patti Vasquez, Joy Wannamaker, Tanailly Giralt Smith, India Pitts, Amanda Beaten, Ebony Harrington, Alex Bradley, Chidinma Okafor, Miriam Chicurel-Bayard, Kristina Shoffner, Polly Tsai, Chelsea Taylor, Susanne Hendersen, Emily Padgett, Debbie Pence, Jane Salm, Matt Campbell, Kirsten Haigler, Ekatherina Diadiuk, Mariam Ramzan, Pamela Miller, Julie Nelson, Nicole Maponga, Carmen Garcia, Charlie McGehee, Gloria Oyediran, Paul Alabanza, William Wolf, Hannah Munro, Rachael Turner, Dana Lapple, Grace Tillotson, Andrew Powell, Mandy Tipton, Catherine Kronk, Oesa Vinesette, Arti Malik, Kirby Caraballo, Maria Stetson, Charles West, Erin Cardot, Andy Thorne, Maria Bullis, William Zhao, Jennifer Thompson, Kristen Gray, Sarah Law, Holly Milner, Frederick Asamoah, Daniel Galeana, Marcia Gibson, Caressa Goss, Pamela Jones, Joshua Lee, Cheryl Hendrickson, Rachel Cook, Erin Daniel, Centhla Washington, Carolina Pastrana-Medina, Dayo Nylander-Thompson, William Johnson, Eliza Debose, Chloe Twomey, Rachel White, Grace Bailey, Hayley Meier, Jennifer Te Vazquez, Ascary Arias, Allison Castillo, Dynesha Perry, Gwen McKnight, Lucie Mangala, Jessica Gingles, Maggie Harman, Marie Oriol, Sean McMurray, Christy Litel, Noshima Darden-Tabb, Yerson Padilla, Danna Frederick | University of North Carolina | Chapel Hill, NC |
| Bruce C. Gebhardt, MD | Padma N. Mangu, Debra Beck Schroeck, Rajesh Kumar Davit, Gayle D. Hennekes, Donna Percy | Sterling Research Group | Cincinnati, OH |
| Carl P. Griffin, MD | Raymond Cornelison, Shanda Gower,William Schnitz, Angela Genovese, Ryan Morgan, Destiny S. Heinzig-Cartwright, April Green, Kim Hamilton, Chalimar Rojo, Lacey Dietz, Sharee Wright, Aja George, Karen Hames, Sharla Lister, Brandy Ball, Andrea Romero, Krystal Hightower, Dalia Tovar, Kim Calloway, Samelia Farni, Chris Hyatt, Linda Lopez, Kathi Shaw, Natacha Tull, Katelyn Hughes, Selwyn Oruh, Lauren Schwab, Samantha Ting | Lynn Health Science Institute | Oklahoma City, OK |
| Joseph Grillo, MD | Amy Potts, Julie White, Carla Bender, Debra Daugomah, Caitlin Harris, Brian White, Alannah Hill, Chelsea Lairson, Karen Blevins | Medical Research International | Oklahoma City, OK |
| Bernard Grunstra, MD | Donald Quinn, Shelby Olds, Phillip Claybrook, Amy Dye, Shai Perry, Joshua Bullen, Jennie Eller, Sandy Daggs, Nicole Everhart, Dennis Lee, Farrah Fuston | PMG Research of Bristol | Bristol, TN |
| Greg Hachigian, MD | Deborah Murray, Michael Cancilla, Logan Ledbetter, Masaru Oshita | Benchmark Research | Sacramento, CA |
| Charles Harper MD | Keith Vrbicky, Chelsie Nutsch, Sally Eppenbach, Wendell Lewis, Alisha Kiepke, Misty Appeldorn, Cyla Rohde, Catherine King, Kayla Andal, Ashley Frisch, Courtney Green, Kelsey Kelley, Katlyn Mace, Jordan Suckstorf, Torie Johnson, Linden DeBoer, Christy Lee, Eric Graber, Jeni Hoppe, Jill Smith, Heather Ebel, Taysha Hingst, Samantha Wieseler, Diahn Pekny, Elijah Schantz | Meridian Clinical Research | Norfolk, NE |
| C. Mary Healy, MD | Chianti Wade Bowers, Chanei Henry, Sheri Ordonez, Janet Brown, Cathy Faw, Shetel Anassi, Trent Davis, Kim Taylor | Baylor College of Medicine | Houston, TX |
| Jeffrey A. Henderson, MD, MPH | Jeffrey A. Henderson, MD, MPH, Marcia O'Leary, RN, Kendra Enright, RN, Jill Kessler, MS, Pete Ducheneaux, LPN, Asha Inniss, MS, APRN | Black Hills Center for American Indian Health / Missouri Breaks Industries Research Inc | Rapid City, SD |
| Ripley Hollister, MD | Jeremy Brown, Melody Ronk, Jill York, Shelby Pickle, Jami Wagner, Lisa Jackson, Felipa Ramdeholl, Angelica Romero | Lynn Institute of the Rockies | Colorado Springs, CO |
| Matthew Hong, MD | Wayne Harper, Lisa Cohen, Priti Patel, Kendra Lisec, Makayla Dutton, Lynn Eckert, Aubrey Farray, Jenee Jiggetts, Emily Reilly, Jill Holmes, Aaron Deaver, Christine Grissom, Judith Shand, Brianca Farmer, Eric Henderson, Kristen Shireman, Brad Muskelley, Franziska Gassaway, Darian Lawrance, Sabine Ucik, Toni Bland, Katedra Dixon, Reginald Santiago, Caroline Zhu, Kathleen Sander, Brian Joseph, Marsha Peery, Lori Bridges, Sadia Khan, Adnan Nasir, Sofia Sequiera, Raquell Messick, Kyra Brown, James Hull | M3-Wake Research | Raleigh, NC |
| Gregory Holt, MD | Jennifer Denizard, Juanita Johnson, Sehrish Sikandar, Gisel Urdaneta, Silvana Cobain, Melyssa Sueiro, Precious Leaks, Evelyn Guadalupe, Rochelle Thompson, Dexter Peart, Leidi Paez, Krystal Hosang, Runxia Tian, Ali Vaeli Zadeh | Miami Veterans Affairs Medical Center | Miami, FL |
| Lilly Immergluck, MD | LaKesha Tables, Harold Gene Stringer, Jacquelyn Ali, Cristina Wilson, Noor Mohamed, Kay Woodson, Tiffany White | Morehouse School of Medicine | Atlanta, GA |
| Michael Jacobs, MD | Kathleen Menasche, Vincent Mirkil,Yazil Ramirez, Michael Yee, Laura Elio, Candice Garcia, Azucena Valdovino, Cristina Garcia, Sharla Peahi-Ching, Kristina Arcos | Alliance for Multispecialty Research | Las Vegas, NV |
| Jeffry Jacqmein, MD | Maggie Bowers, Dawn Robison,Victoria Mosteller, Janet Garvey, Alpa Patel, Darlene Bartilucci, Kenneth Aung-Din, Margaret Gannaway, Carolyn Tran, Michael Koren, Mitchell Rothstein, Sonia Gerardo, Cassie Lawler, Yvonne Douglas, Chris Ganzhorn, Emery Noles, | Jacksonville Center for Clinical Research | Jacksonville, FL |

| | | | |
|---|---|---|---|
| | Angela Morris, Lisa Carl, Andrea West, Laura Little, Ramil Castillo, Abbey Ras, Nalini Jones, Annan Nurrenbern, Deirdre Arrington, Jacob Wolfer, Brenda Anderson, Amanda Elwood, Amber DeVries, Cara Seifart, Jimmy Knowles, Vy Dang, Mary Strickland, Pam Garmon, Caron Whitelaw, Sharon Smith, Ivy Guillermo, Nate Grant, Khatija Hussein, Caron Whitelaw RN, Bernadette Moineau, Robert Nix | | |
| Robert Jeanfreau, MD | Susan Jeanfreau, Katelyn Jackson, Kynisha "Nicki" Johnson, RaeShanta McKendall, Shonna James, Calisha Sadiq, Susan Tortorich, Lori Goins, Steven Darden, Melissa Spedale, Kristen Robinson, Joseph Favret, Yordanka Koleva, April Spears, David Conroy | MedPharmics | Metairie, LA |
| William Jennings, MD | Hilario Alvarado, Michele Baka, Malina Regalado | Synexus Clinical Research | San Antonio, TX |
| Carol Kauffman, MD | Andrea Starnes, Andrea Woods, Karen Brudzinski | VA Ann Arbor Healthcare System | Ann Arbor, MI |
| Colleen F. Kelley, MD, MPH | Carlos del Rio, Sheetal Kandiah, Catherine Abrams, Erin Andrew, Felicia Atkinson, Erica Baker, Juliet Brown, Tucker Colvin, Natasha Renee Cook, Meena Dhir, Christopher Foster, Ronald Gaston, Gabriela Gerogial, John Gharbin, Betsy Hall, Valarie Hunter, Aastha KC, Kelly Likos, Bezuayehu Mandefro, Myles Mason, Humberto Orozco, Isaac Perez, Philip Powers, Christin Root, Brittany Spiegel, Pamela Weizel, Sarah Wiatrek, Felicia Wright | Ponce de Leon Center | Atlanta, GA |
| Jeff Kingsley, DO | April Pixler, LaKondria Curry, Sarah Afework, Austin Swanson, Alyssa Middlebrook, Christine Senn, Keyrhea Ritter, Katlin Salewski, Sierra Holmes, Jean Niles, Taylor Hernandez, Lacey Shaw, Kaila Maddox, Klarissa Bohnstedt, Emily Gilder, Cassandra Motley, Alyssa Middlebrook, Hephzibah Udo, Mattison Sherer, Wayman Petty, Joseph Surber | IACT Health | Columbus, GA |
| Judith Kirstein, MD | Marcia Bernard, Erica Sanchez, Nolan Mackey, Clarisse Baudelaire, Hanna He, Brenda Delgado, Brandon Steppe, Bonnie Goodale, Nicole Abels, Carol Remigio, Dipal Patel, Emily Zacarias, Nuvia Espinoza, Esmeralda Machado, Katia Talamante, Lizeth Romero | Velocity Clinical Research | Banning, CA |
| Susan Kline, MD, MPH | Sara Eischen, Rebecca Cote, Diondra Howard, Editha Jordan, Joyce Bolea, Annie McFarland, Asfaw Mesfin, Andrew Snyder, Darlette Luke, Derek LaBar, Theresa Christiansen, Beth Jorgenson, Christina Glasgow, Melissa Schedler | University of Minnesota | Minneapolis, MN |
| Mark E. Kutner, MD | Mark E. Kutner, Jorge Caso, Janet Mendez, Maria Hernandez, Carmen Amador, Amanda G. Colina, Alain Chang, Alondra Diaz, Arael Ayala, Carmen Ballester, Claudia Rodriguez, Dalila Del Valle, Eduardo Rodriguez, Gloria Moreno, Jennifer Ortega, Jhobana Vargas, Jonathan Fernandez, Juan Carlos Delgado, Laura Gonzalez, Leidy Montoya, Marianela Carvajal, Mariete Rendon, Maury Santos, Michelle Browne Mirnaya, Mujica, Neiner Enriquez, Noelio Hernandez, Paola Garcia<br>Raydel Valdes, Saray Carvajal, Susel M. Figueredo, Vanessa Hechevarria, Yanelis Dominguez, Yusleidy Diaz | Suncoast Research Group | Miami, FL |
| Mark Leibowitz, MD | Fernanda Morales, Rosario Sanchez, Mike Delgado, Norma Vega, Nelly Ayala, Iliana Gallaga, Cassandra Celis, Jennifer Muniz, Mariela Quiroz, Juan Frias, Rea Abaniel, John Nelson, Maricor Grio, Alejandro Moreno, Coralia Soto, Jose Espino, Daniel Vargas, Stephanie Lopez | National Research Institute | Los Angeles, CA |
| Derek Lewis, MD | Fred Newton, Aeiress Duhart, Breana Watkins, April Green, Chala Simpson, Briana Dean, Brandy Ball, Shakita Stevenson, Lashonda Stephenson | Lynn Institute of the Ozarks | Little Rock, AR |
| Gregg Lucksinger, MD | Jaleh Ostovar, Audrey Kuehl, Viviana Juncal, Avery Kerwin | Velocity Clinical Research | Medford, OR |
| Benjamin J. Luft, MD | Jorge Alves, Melissa Carr, Ryan Chacon, Barsha Chakraborhy, Aymon Faizi, Laurel Gumpert, Andrew Handel, Kayla Henkel, Erin Infanzon, Andrew Kanner, Lily Limsuvanrot, Michelle Miroddi, Jeanine Morelli, Sharon Nachman, Rena Nanan, Alexander Newman, Alison Pellecchia, Trisha Rush, Jennifer Russell, Stephanie Santiago-Michels, Jonathan Sicoli, Candace Smith, Michael Truhlar, Bruno Valenti, Jennifer Valentine, Kathy Vivas, Yasmine Brown-Williams | Stony Brook University - Stony Brook Medicine | Commack, NY |
| Siham Mahgoub, MD | Alice Ukaegbu, Immaculate Okonkwo, Shannon Gopaul, Tara Gibbons, Yuanxiu Chen, Debra Ordor, Linda Fletcher, Megan Ware, Florencia Gonzalez, Michael Perini, Carla Williams, Mulu Mengistab, Robert Postell, Yejide Obisesan, Adetokunbo Adedokun, Reyneir Magee, Jeremy Smith, Edward Bauer, Lora Collins, Urelda Allman, Deborah Clements, Sarah Shami, Nathaniel Blaboe, Pedro Lima, Michael Crawford | Howard University Hospital / Howard University College of Medicine | Washington, DC |
| Mary Beth Manning, MD | Toby Briskin, Denise Roadman, Sarah Dzigiel, Jennifer Gaston, Brooke Glivar, Brianna Arman, Briana Jackson, Brian Sharpe, Naqib Ahmad, Nicole Baitt | Velocity Clinical Cleveland | Cleveland, OH |
| Rickey D. Manning, MD | James Wilson Hurst, Rodney E. Sturgeon, Paul H. Wakefield, John A. Kirby | Accellacare | Knoxville, TN |
| Kristen Marks, MS, MD | Marshall Glesby, Roy Gulick, Timothy Wilkin, Ole Vielemeyer, Mary Vogler, Carrie Johnston, Rebecca Fry, Daniel Finn, Caitlin Rhoades, Noah Goss, Shaun Barcavage, Valery Hughes, Jonathan Berardi, Ashley Machado, Caique Mello, Mia Crowley, Monique Williams, Minkyung Lee, Mary Ann Zwiebel, Patrice Weller, Antonio Rivera-Lopez, Harrison Chan, Ruby Lee, Victoria Lesina, Vasilika Koci, Paul Kim, Steven Wang, Malissa Robinson, Edward Kenny, Danny Garcia, Venus Fernandez, Parul Shah, Celine Arar, Byron Bullough, Jonattan Rodriguez, Jessenia Fuentes, Jiamin Li, Arthur Goldbach, Genessi Rodriguez, Catherine Jerry, Nadi Islam, Madeline Gomez, Rajshri Hirpara, Ioanna Pahountis, Wayne Burns, Tahera Begum, Gianna Resso, Sophia Alvarez, Elizabeth Connolly, Roxanne Rosario, Sierra Derti, Britta Witting, Anna Gwak | Weill Cornell Chelsea CRS | New York, NY |
| Paul G. Matherne, MD | Cassie Beeks, Sarah Bowen, Deven Fejka, Nicole Guttierrez, Lakeyla Bates, Pam Taylor, Gigi Benoit, Micki Le | Medpharmics | Gulfport, MS |
| Monica McArthur, MD, PhD | Cheryl Young, Helen Powell, Levis Contreras, Panagiota Komninou, Christine Wade, Jumoke Oladapo, Kaitlin Mason, Robin Barnes, Leslie Howe, Cheilon Bolanos, Shannon Bittner, Elva Valle-Maldonado, Wanda Somrajit, Biraj Shrestha, Justin Ortiz, Nancy Greenberg, Kathleen Strauss, Lisa Chrisley, Melissa Billington, Sudhaunshu Joshi, Lavida Porter, Megan McGilvray, Daryl Grays, Shirley George, Jennifer Marron, Kelly Brooks, Natelaine Fripp, Mardi Reymann, Brenda Dorsey, Patricia Farley, Melissa Myers, Natasha | University of Maryland School of Medicine | Baltimore, MD |

| | | | |
|---|---|---|---|
| | Harris, Alyson Kwon, Marcela Pasetti, Daniel Cohen, Myounghee Lee, Laura Liberman, Sherry McCammon, James Campbell, Ana Herbert, Julia Silva, DeAnna Friedman-Klabanoff, Alythia Vo, Jennifer Winkler, Lisa Turek, Colleen Boyce, Anne Thurston, Daniele Nitkowski, Ginny Cummings, Sandra Molina, Susan Holian, Matthew Laurens, Rekha Rapaka, Megan Deming, Mark Travassos, Kirsten Lyke, Henry Seifert, George Escobar, Norma Martinez, Abigail Arias, Ana Maria Davila, Dolores Fontalvo, Elsa Aracely Vargas, Elva Jaldin, Gladis Lopez, Irma Justiniano, Judith Tenezaca, Julio Fernandez, Luz de Maria Osorio, Maria de los Angeles Pichardo, Morena Lemus, Maria Elena Rocha, Rosa Angelica Vigil, Sandra Herrera | | |
| R. Scott McClelland, MD, MPH | Devinder Garcha, Christopher P. Hawk, Bonnie Duran, Donna Lodge-Moore, Leigh Tao, Cristina J. Toledo-Cornell, Mona Jalili, Susan Lottimer, Sheila Samra, Seslee Alsup, Karlee Cooper, Theresa George-Greene, Spencer Hanson, Joni Hensley, Emily Barnett Highleyman, Jewell Jefferson, Jessica Lane, Jessica Long, Alex Martinez, Kerri Sloan, Kelly Smith, Kristee Lewis, Tara Babu, Dwyn Dithmer, Matthew Dustrude, McKenna Eastment, Emily Ford, Abir Hussein, Christine Johnston, Pamela Kohler, Debra Metter, Thepthara N. Pholsena, Meredith Potochnic, Tara Reid, Miko Robertson, Michelle Sabo, Helen Stankiewicz Karita, Jina Taub, Dana Varon, , Brian Wood, Alyssa Braun, David Crawford, Mark Drummond, Jess Heimonen, Lawrence Hemingway, Madelaine Humphreys, Bianca Kalia, Mary Kirk, Taylor Krause, Ray Larsen, Gisella Logioia, Cristina Luevano Santos, Anya Mathur, Lindsey McClellan, Jessica Moreno, Nicole Roed, Matthew Seymour, Katie Wicklander | University of Washington & Lummi Tribal Health Center | Seattle, WA |
| Bruce McClenathan, MD | Mary Hussain, Aaron Poch, Amy Santangelo, Anne Poch, David De Blasio, Evelyn Lomasney, Jacob Turnquist, Kathryn Lago, Laurie Housel, Sherry Lamberth, Sheryl Bedno, Lauren Blevins, Laura Brown, Alice Clay, Gervon Collins, Kaitlyn Covington, Amy Davis, Patricia Davis, Nicole Friedberg, Lacey Gazlay, Helen Gooden, Evelyn Hall, Kim Locklear, Kayla Majors, Shamona McRae, Bryce Meerhaeghe, Kendalyn Stephens, Jade Tran, Lisette Watkins, Katie Williams, Kema Matthews, Karrie Greive, Brandi Carroll, Amanda Williams, Brittany Garner, DeLisa Crosby, Jennifer Ritschl, Jamie Frahm, Karen Stewart, Priti Patel, Dilay Uras, Allison Northrop, Anika Uson, Olyvia Ray, CynDavia McKoy, April Beals, Deidre Turner, Christina Spooner | Womack Army Medical Center | Fort Bragg, NC |
| Mark McKenzie, MD | Teresa Deese, Christy Schmeck, Vickie Leathers, Christy Sweet, Misti Earwood, Erica Osmundse, Gisela Heintz, Lilian Nunkuna, Michelle Forgey, Shelly Brooks, Justian Jarrett, Elizabeth Michael, Lisa Guider, Zack Harmon, Diane Sproles, Randy Cooper, Jessica Benvenuto, Stefanie Mullins, Quinetrice Bennett, Corey Flack | WR Clinsearch | Chattanooga, TN |
| Jorge F. Méndez Galván, MD | Adriana Sordo Durán, Martha Yarelli Valencia Mejía, Froylan David Martínez Sánchez, Ana María Piña Rodríguez, Diana Alim Mena Martínez, Melany Susel Fernández Valdez, Laura Ruy Sánchez Guerrero, Ana Fabiola Ruiz Villagrana, Mónica B. Carrascal, Martha Cecilia Gomora Madrid, Anahí García Álvarez, Ismael Delgado Ginebra, Omar Alfonso Heredia Nieto, Yanni Maldonado Ventura, Jonathan E. Ramírez Salazar, Mariela Salgado Zagal, María Fernanda Espinosa García, Yolanda Albor Hernández, Ricardo Antonio González, Germán Alonso Lara, Marbella Rojas Ortega, Bernardo Kleinfinger Chayet, Victor Emmanuel Alva López, Diego Carlos Angel Perez, Alejandro Cortes Meda,Ivonne Hernandez Giron | Centro de Atención e Investigación Médica (CAIMED) | Mexico City, Mexico |
| Rajan Merchant, MD | Anelgine Crans Yoon, Janet Hill, Lucy Ng-Price, Teri Thompson-Seim, Alejandra Cazares Hernandez, Danielle Hornbuckle, Adriane Rubit, Ann Campbell, Dawn Diorio, Adeline Stabler, Jasdeep Shergill, Claudia Gross, Anne Nguyen | CommonSpirit Health Research Institute | Woodland, CA |
| Vicki E. Miller, MD, MPH | Amy Starr, Shiela Varghese, Sonia Guerrero, Monica Murray, Vanessa Gonzales, Blanca Gomez, Zainab Rizvi, Victoria Aguilar, Anna Pena, Madiha Baig, Dustin Watson, Pauline Ngban, Afifah Ayub, Laura Drampou, Shelby Danforth, Diana Avalos, Jacquelyn Gonzales, Ragen Powell, Sajjad Naqyi, Ambily Dileep, Alefiyah Motiwala, Heather Leary, Humera Siddiqui, Miatta George, Kastyn Kelly, Nicole Segura, Maryam Jamil, Husain Motiwala, Sandra Smith, Sally Hussein, Yousra Yousif, Carlyn Robinson, Cannon Lenfield, Luis Leal, Muhammad Irfan, Nayab Croher, Pattie Tate, Sandra Natalia Perez, Fredric Santiago, Syeda Riaz, Arsani Iskandar, Alefya Hussain | DM Clinical Research | Tomball, TX |
| Linda Murray, DO | Christy Delcamp, Monica Hoewt, Kristin Shade, Tara McTigue | Synexus Clinical Research | Pinellas Park, FL |
| David Musante, MD | William P. Silver, Linda R. Belhorn, Nicholas A. Viens, David Dellaero, Shandelle Parker, Andrew Zimmerman, Roger Ordronneau, Bryan Stanislaus, Kevrin Johnson, Megan Dice, Megan Heron, Sarah Wilkerson | M3-Emerging Medical Research | Durham, NC |
| Sherif Naguib, MD | Justin Singletary, Sha-Wanda Richmond, Sarah Omodele, Emily Oppenheim, Jalisha Hemphill, Marqueta Jones, Millat Gedefa, Janean Smith, Bonnie Raufman, Lesley Whitehead, Elia O'Dell, Sarah Omodele, David Taylor, ShaWanda Richmond, Alexis Melson, Justin Singletary | Synexus Clinical Research | Atlanta, GA |
| Joseph Newberg, MD | Laura Pearlman, Reuben Martinez, Victoria Andriulis, Jacquilyn McCormick, Anna Maddox, Rosalinda Vazquez, Nicole Leahy, Marian Padilla, Mary Reyes | Synexus Clinical Research | Chicago, IL |
| Paul J. Nugent, DO | Leonard Singer, Jeanne Blevins, Meagan Thomas, Christine Hull | Synexus Clinical Research | Cincinnati, OH |
| Yaa D. Oppong, MD | Ryan P. Starr, Scott N. Syndergaard, Nafisa Saleem, Cheryl Norris, Nicole Austin, Rozeli Shelly, Md Mashrur Islam Majumder, Annette Bunnells, Michelle Wallace, Avia McClain-Stocker, Rachel Ryan, Katie Wood, Arien Stebbins, Crystal Schmitt, Jeffrey Pemberton, Mitchel Arlidsen, Daniel Tomita, Geraldine McRae, Amy Sheets, Jeanette Mangual-Coughlin,<br>Margo Miller-Smith, Melinda Thomas | Carolina Institute for Clinical Research | Fayetteville, NC |
| J. Scott Overcash, MD | Adrienna Marquez, Hanh Chu, Kia Lee, Kim Quillin, Jordan Coslet, Yashveer Dubbula, Adam Prince, John Rodriguez, Lee Tomatsu, Erin Vawter, Michael Voskanian, Michael Waters, Gina Weaver, Karla Zepeda, Angela Anorve, Gordon Bovee, Jennifer Baker, Laura Castillo, Allie Davis, Jacob Esparza, Andrea Garcia, Jessica Gonzales, Lizette Gonzalez, | Velocity Clinical Research | La Mesa, CA |

| | | | |
|---|---|---|---|
| | Ashleigh Lindsay, Erica Marinelli, Cathy Meza, Shandel Odom, Makenna Orel, Grecia Perez, Helen Pu, Cesar Ramirez, Melania Riordan, Deidre Romines, Raquel Taitingfong, Katrina Tyler, Bernadette Wilson | | |
| Yogesh K. Paliwal, MD | Amit Paliwal, Renu Bhupathy, Krystle Edwards, Sarah Gordon, Cynthia Montano-Pereira, Blanca Gomez, Yazmin Nunez, Cassandra Martinez, Connie Navarrete, Mayra Casas, Ysabel Lopez, Anthony Macias, Alexandria Vasquez, Maria Gomez | Empire Clinical Research | Pomona, CA |
| Isabel Pereira, MD | Gina Rivero, Tracy Okonya, Frances Downing, Paulina Miller, Yasmin Camberos | Synexus Clinical Research | Vista, CA |
| Bruce Rankin, DO | John M. Hill, Steven Shinn, Vivek Rajasekhar, Marshall Nash, Michelle Tutt, Kimberlee Del Campo, Douglas F. Winter, Leandro Fernandez, Melissa Hodges, Michelle Jones, Sean Lemoine, Veronica Walker, Roy D. Richardson, Angeline Petracca. Katina Marchione, Michelle Morgan, Ashley McCaffrey, Amber Vasquez, Amy Houck-Dominy, Angela Hammerle, Antonio Rivera, Claxton Copeland, Crystal Paccione, Diana Toney, Fadhel Alyunis, Jennifer Dittman, Kriston Applewhite, Lora Parahovnik, Over Seijas, Ryan Hobbick, Samantha Watts, Shatonia Fields, Stacie Evans, Teresa Logsdon, Thais Truffa, Tifany Huertas, Vienna Bauer,William Serrano, Daisy Sawyer, Giovanni Urquilla, Tonya Toby, Albert Garcia, Alicia J. Cevera,Jeffery Hood, Hannah Hodges, Melissa Willard | Accel Research Sites | DeLand, FL |
| María José Reyes Fentanes, MD | Pablo Fermín González Limón, Luis Ricardo Acosta Beuló, Paulina Cleer García Valdovinos, Olivia de la Puente Flores, Eduardo Rugama Martel, Ana Gabriela Mier Flores, Ulises Abel Rodríguez Vargas, Diego Guillermo Muñoz Bolaños, Martha Alejandra Alonso Trejo, Elvia Ramírez Gutiérrez, Alberto Aaron del Rosal Medina, Jaime Chavez Baron, Ana Gabriela Guizar Zamora, Felipe Arredondo Saldaña, Juan De Dios Martín Luján Palacios, Juan José Pardo Moreno, Jorge Torres Ferrera, Itzetl Guzman Mendieta | PanAmerican Clinical Research México | Querétaro, Mexico |
| Margaret Rhee, MD | Jeffrey Klein, Katherine Stapleton, Stacy Collins, Dawn Greer, Kelli Meissner, Brenda Moore, Tylene Falkner, Celeste Blazy, Nicole Johnson, Christina Carter, Annette Pangle, Rosamond Hong | Synexus Clinical Research | Akron, OH |
| Robert Riesenberg, MD | Robert Riesenberg, Stanford Plavin, Mark Lerman, Leana Woodside, Maria Johnson | Atlanta Center for Medical Research | Atlanta, GA |
| Barbara Rizzardi, MD | Michelle King, Vanessa Abad, Jennifer Knowles, Benjamin Richeson, Denise Pessetto, Heather Holtman, Lori Luth, Wyatt Walsh, Andrea Johnson, Dreama Fackrell, Patrick O’Keefe, Sara Isolampi, Michelle Walkingshaw, Josh Carrillo, Renu Landage, Stephanie Wallace | Velocity Clinical Research | West Jordan, UT |
| Carina A. Rodriguez, MD | Patricia Emmanuel, Lucy Guerra, Asa Oxner, Alicia Marion, Reed Ryan, Tiffany Vasey, Susannah Hall, Amanda Morton, Emma Gonzalez, Elisabeth Ballans, Rachel Karlnoski, Luz Santamaria, Rosalinda Cruz, Joshua Finley, Michael Hayes, Oliver Emberger, Mark Pennington, Meghana Vankatesh, Kimberly Johnson, Marina Wassif, Janelle Perkins, Veroniya Winkfield, Amavyvis Garcia, John Jones, Lori Brock, Kyle Cesareo, Dilcina Dragon, Dominic Moore, Catherine Marten, Thi Nguyen, April Roberts, Kristi Bojaxhi, Chrestenie Mouse | University of South Florida, Morsani College of Medicine | Tampa, FL |
| David Rosenberg, MD | Lee Tomatsu, Viviana Gonzalez, Millie Manalo, Nicole Rudin | Pharmacology Research Institute | Los Alamitos, CA |
| Vida Veronica Ruiz Herrera, MD | Vida Veronica Ruiz Herrera, Eduardo Gabriel Vazquez Saldaña, Laura Julia Camacho Choza, Karen Sofia Vega Orozco, Sandra Janeth Ortega DominguezMaria, Carolina Molina Roman, Julian Camacho Choza, Rodolfo Fabian Lomeli Guerrero, Giuliana Magaña Garcia, Carlos Andres Perez Navarro, Cesar Alberto Lopez Martin, Luis Arturo Rico Godinez, Felipe de Jesus Lopez Cordova, Daniel Arroniz Bernal, Luisana Aldaco Cota, Edgar Cordova Pulido, David Aguila Rivera | PanAmerican Clinical Research México | Guadalajara, Mexico |
| Beth Safirstein, MD | Luz Zapata, Lazaro Gonzalez, Evelyn Quevedo, Farah Irani, Julio Vigil, Steven Rapp, Mark Firestone, Humberto Mucientes, Ali Yasells Garcia, Florence Baum, Robert Hacman, Martha Ravelo, Carlos Alzate, Keyanna Francois, Alberto Napoles, Jamie Lorenzo, Deandra Clarke, Disneydi Gutierrez, Yean Alfonso, Nestor Lopez, Ana Bustos, Ilya Faybishenko, Lynnette Perez, Evelyn Qulies, Maria del Valle, Natalie Joseph, Judith Powell, Jessica Hernandez, Rafael Sanchez, William Torres, Damaris Alonso, Dragos Juravle, Roberto Valledor, Veronica Valledor, Maria Pazos, Teresa Rios, Maria Lascano | MD Clinical | Hallandale Beach, FL |
| Howard Schwartz, MD | Nelia Sanchez-Crespo, Terry Piedra, Barbara Corral, Jennifer Schwartz | Cenexel RCA | Hollywood, FL |
| Elizabeth Secord, MD | Roy Collins, Marita Poff, Jamal Chehab, Sajith Matthews, Thomas Mazzocco, Chantel Karmo, Sarah Meram, Janie Faris, Valerie Mika, Shobi Mathew, Brian O’Neil, James Paxton, Amy Stolinski, Stacie Smith, Benjamin Wasinski, Lisa Palmer, Katherine Cross, Samuel Ceckowski, Theodore Falcon, Jeffrey Harrison, Abe Lovelace, Selmir Mahmutovic | Wayne State University | Detroit, MI |
| Marian E. Shaw, MD | Mark A. Turner, Cory J. Huffine, Esther S. Huffine, Jacqueline Hanson, Nicholas Tuttle, Shannon Veach, Antonio Navarrete, Jammie Smith | Velocity Clinical Research | Meridian, ID |
| Teresa S. Sligh, MD | Scott Sligh, Parul Desai, Vincent Huynh, Carlos Lopez, Erika Mendoza, Dennis Perez, Samuel Ceballos, Jennifer Gomez, Janneth Becerra, Tiffany Martinez, Erika Navarro Fausto | Providence Clinical Research | North Hollywood, CA |
| Joel Solis, MD | Carmen Medina, Westley Keating | Centex Studies | McAllen, TX |
| Jonathan Staben, MD | Jessica Horton, Hannah Neill-Gubitz, Hilary Koenigs, Autumn Dlugas, Stacie Rebar, Anne Reedy, Roslyn Pierce, Kali Karst, Jaimee Gribben, Sarah Troutt, Mimi Meipel, Ann Carson, Paige Ramos, Natosha Hardy, Zack Brownell, Dot Heid, Annie Estes, Andrea Fry, Veronica Navarro | MultiCare Institute for Research and Innovation | Cheney, WA |
| Kathryn E. Stephenson, MD, MPH | Karen A. Lorenc, Audrey B. Nathanson, Michelle Beck, Shaelah M. Huntington, Wendy Hori, Uyen Rasphoumy, Ashley Beckles, Jody Dushay, Vijai Bhola, Wilanda Gabriel, Annika Gompers, Halle Hall, Nicholas Manickas-Hill, Toluwanimi Ajayi, Nicole Magner, Conor Cronin, James Arrico, Heena Patel, Janet Mullington, Michael Seaman, Katherine Yanosick, Ariana Leonelli, Eric Dai | Beth Israel Deaconess Medical Center | Boston, MA |
| Danny Sugimoto, MD | Jeffrey Dugas Sr., Dolores Rijos, Sandra Shelton, Stephan Hong | Cedar Crosse Research Center | Chicago, IL |
| Suzanne Swan, MD | Sharine Phan, Tami Wahlin, Elizabeth Bennett, Amy Salzl, Jeannette Blaisdell, Stacie Mahowald, Dominick Thibodeau, Sophia Houser, Tammy Hanson | Synexus Clinical Research | Richfield, MN |

| | | | |
|---|---|---|---|
| Karen Tashima, MD | Helen Patterson, Stacey Chapman, Giselle Pinto, Jennifer Brashears, Evelyn Hipolito, Laura Elmasian, Timothy Flanigan, Joseph Garland, Britt Harrington, Anthony Harrison, Jenny Thai, Mazen Taman, Krista Kiser, Kay Rutherford, Shivani Patel, Jimin Shin, Kim Rapoza, Sujata Sahu, Kristine Hauser, Kendra Vieira, Elliott Bosco, Christopher Federico, Kanika Malani, Christian Schroeder, Janet O'Connell, Meghan McCarthy, Anna Hippchen | The Miriam Hospital | Providence, RI |
| Barbara S. Taylor, MD, MS | Bhoja Katipally, Jessica Blower, Kimberly Kone Ellis, Heta Javeri, Danielle Dixon, Anna Taranova, Diana Cavazos, Robin Tragus, Irma Scholler, Lisa Longoria, Laura Najvar, Meredith Hosek, Bridgette Soileau, Morgan Brown | University of Texas Health Science Center San Antonio | San Antonio, TX |
| Christine B. Turley, MD | Lewis McCurdy, Tonisha Brown, Martha Pawlicki, Jennifer Reeves, Jona Bauer, Cedrick Griner, Cameron Russell, Veena Sampathkumar, Zeynep Alimchandani, Robin Muller, Tracey Coakley, Mary Sours, Saifelnasr Mohamed, Sone Alanoh, Amy Yeh, Sahra Khan, Eleojo Abutu, Genena Buck, Sarah Hicks, Andreana Alexander, Tammy Patterson, Maria Martilnsalaco, Amy Clontz, Marina Leonidas, Zainab Shahid, Jay I. Patel, Ryan Bender | The Charlotte-Mecklenburg Hospital Authority d/b/a Atrium Health | Charlotte, NC |
| Lisa S. Usdan, MD | Lora J. McGill, Valerie K. Arnold, Carolyn Scatamacchia, Codi M. Anthony, Carol R. Marsh, Cathy T. Houpt, Charles L. Grandberry, Debra A. O'Brien, Kelsey N. Evans, Leslie M. Lazar, Mary J. Williams, Megann F. Fickle, Robyn M. Presley, Shelby R. McWhorter, Julia Sinatra, Irene W. Powell, Tavia S. Flagg, Melissa N. Flowers, Penny J. McCracken, Reagan A. Boone, Dominique L. Ross, Amber J. Jones, LaKeshia N. Pipkin, Victoria J. Neal, Monica Toor, Brandi Gruber, Erin L. Wells, Kelly Iskiwitz, Carolyn J. Scatamacchia, Codi M. Anthony, Lisa S. Usdan, Lora J. McGill, Valerie K. Arnold | Clinical Neuroscience Solutions | Memphis, TN |
| Larkin Tyler Wadsworth III, MD | Horacio Marafioti, Lyly Dang, Lauren Clement, Kristen Johnson, Anya Penly, Elizabeth Garner, Angie Kean, Sophia Bolakas, Andrea Deffenbaugh, Cerece Miles, Lindsay Nooter, Christy Shultz, George Cherniawski, Stephanie Tesson, Ash Dale, Laura Hartupee, Breanna Galibert, Karen Knapp | Sundance Clinical Research | St. Louis, MO |
| Michael Waters, MD | Dalia Tover, Scott Overcash, Jordan Coslet, Michael Voskanian, Giuliano Zolin, Matthew Petro, Gina Weaver, Kia Lee, Hanh Chu, Karla Zepeda, Crystle Rajania, John Rodriguez, Tracey Fabrega, Kaitlyn Sandler, Alex Tapia, Cecilia Barbabosa, Renee Pasion, Jacob Pineda, Rosalynn Landazuri, Angelica Franco, Estee Garcia, Marilynn Rodriguez, Joanna Ocampo | Velocity Clinical Research | Chula Vista, CA |
| Jordan Whatley, MD | Jordan Whatley, Christopher Dedon, Emily Best, Amie Breaux Shannon, Mary Margaret Dobson, Nicole Harrell, Lindsey Kobetz Hall, Kristen LeBleu Losavio, Patricia Whatley, Tana Bourgeois, Alexandra Caillouet, Samantha Brooke McMillon, Amy Thomassie, Donna Michelle Hurst, Michelle Symms, Lyndsea Folsom, Crystal Rowell, Loney Girod, Lauren Sternfels, Makaylea Truitt, Lori Martin, April Mims | Meridian Clinical Research | Baton Rouge, LA |
| Jewel Johnny White, MD | Amanda Occhino, Ruth Paiano, Morgan McLaughlin, Elisa Swieboda | Synexus Clinical Research | The Villages, FL |
| Hayes Williams, MD, PhD | LaShondra Cade, Mitzi Roberts, Aileen Cunningham, Rhodna Fouts, Connie Moya, Gary Boyd, Justina Owens, Abby Wellinghurst | Achieve Clinical Research | Birmingham, AL |
| Clint Wilson, MD | Jason Milligan, Danielle Raley, Joseph Bocchini, Carrie Kay, Shannon Saksa, Courtney Harmon, Ashley Primos, CJ McKenna, Star Roberts | Willis-Knighton Health System / WKB Family Medicine Associates | Bossier City, LA |
| Peter J. Winkle, MD | Amina Z. Haggag, Elizabeth Lee, Michelle Haynes, Marysol Villegas, Sabina Raja, Mary Grace Lejarde, Caroline Villanueva, Natalie Ureno, Jessica Cramer, Steven Garcia, Yesenia Barraza, Ashley Barajas, Lauren Ferreira, Lucy Rems, Zaki Abawi, Damon Pineda, Patricio Ordonez, Gaby Huizar, Lesbia Alarcon, Anna Luz Belarmino, Nenita Llarena, Alberto Heshike, Isabel Rangel, Karen Cruz, Rynel Villanueva, Matthew Rohrig, Han Tran, Axl Dyer, Maria Webb, Akihisa Kodama, Cynthia Juarez, Sandra Gaona, Moriah Wilson, Mark Gonzalez | Anaheim Clinical Trials | Anaheim, CA |
| Patricia L. Winokur, MD | N/A | University of Iowa Medical Center | Iowa City, IA |
| Paul E. Wylie, MD | Renea Henderson, Natasa Jenson, Fan Yang, Amy Kelley, Kelly Knight, Jessica Watson, Stacy Tierney, Emily Knight, Jessica Woosley, Faith Fields, Glen Scott Thrower | Preferred Research Partners | Little Rock, AR |
| Carmen D. Zorrilla, MD | Carmen Irizarry, Gloria Martino, Natalia Muler, Lázaro Valdés | Universidad de Puerto Rico - Recinto de Ciencias Médicas - Maternal Infant Studies Center (CEMI) | San Juan, Puerto Rico |

**United States Government (USG)/Coronavirus Prevention Network (CoVPN) Biostatistics Team**

| Affiliation | Team Members |
|---|---|
| Biomedical Advanced Research and Development Authority (BARDA), Washington, DC | Di Lu, James Zhou |
| Department of Biostatistics and Bioinformatics, Rollins School of Public Health, Emory University | David Benkeser, Sohail Nizam |
| Vaccine and Infectious Disease Division, Fred Hutchinson Cancer Center, Seattle, WA | Jessica Andriesen, Bhavesh Borate, Lindsay N. Carpp, Andrew Fiore-Gartland, Youyi Fong*, Peter B. Gilbert*, Ying Huang*, Yunda Huang*, Ellis Hughes, Ollivier Hyrien, Holly E. Janes*, Michal Juraska, Yiwen Lu, April K. Randhawa, Brian Simpkins, Brian D. Williamson*, Lars W.P. van der Laan, Chenchen Yu |
| Biostatistics Research Branch, NIAID, NIH, Bethesda, MD | Michael P. Fay, Dean Follmann, Martha Nason |
| Department of Biostatistics, University of Washington, Seattle, WA | Marco Carone, Avi Kenny, Kendrick Li, Wenbo Zhang |
| Department of Statistics, University of Washington, Seattle, WA | Alex Luedtke |
| Division of Biostatistics, School of Public Health, Department of Population Health Sciences, Weill Cornell Medicine, New York, New York | Nima S. Hejazi |
| Department of Population Health Sciences, Weill Cornell Medical College, New York, New York | Iván Díaz |

*YF, PBG, YiH, and HEJ are also affiliated with the Department of Biostatistics, University of Washington, Seattle, WA. PBG and YuH are also affiliated with the Public Health Sciences Division, Fred Hutchinson Cancer Research Center, Seattle, WA. YuH is also affiliated with the Department of Global Health, University of Washington, Seattle, WA. Brian D. Williamson is also affiliated with Kaiser Permanente Washington Health Research Institute, Seattle, Washington, USA.

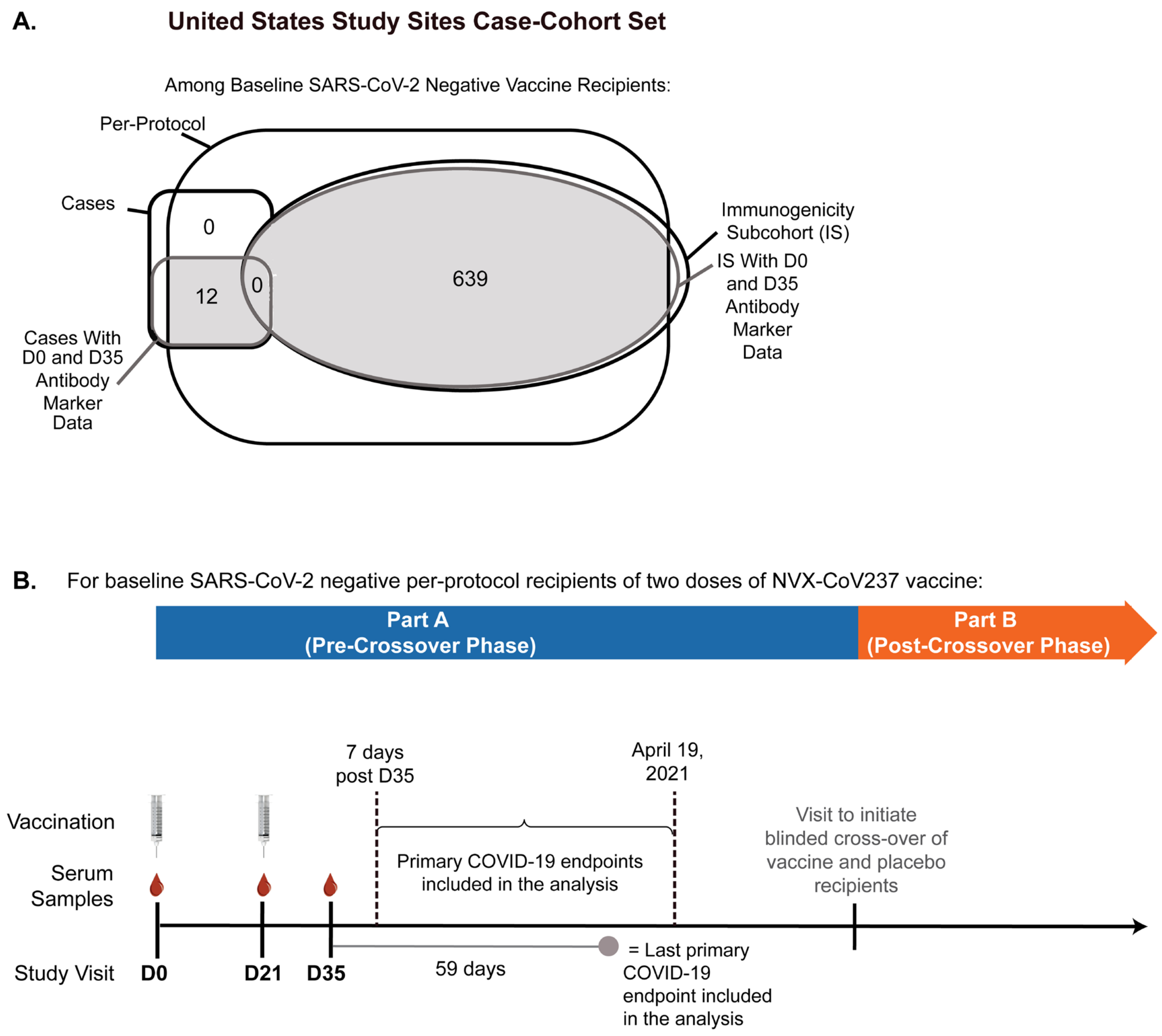


**Supplementary Figure 1. (A) Case-cohort set (U.S. study sites). (B) Phases of the PREVENT-19 trial, timing of NVX-CoV2373 doses and serum sampling, and the time period for COVID-19 primary endpoints included in the Day 35 marker correlates analysis (correlates analyses restrict to Part A Pre-Crossover Phase and U.S. study sites).** In (A), cases are baseline SARS-CoV-2 negative per-protocol vaccine recipients with the primary COVID-19 endpoint starting 7 days post D35 visit through to the efficacy data cut (April 19, 2021). “Baseline SARS-CoV-2 negative” is defined as in ref.[1], i.e. seronegative for anti-SARS-CoV-2 nucleoprotein and SARS-CoV-2 RNA RT-PCR-negative nasal swab at baseline. “Per-protocol” is also defined as in ref.[1], i.e. received both planned vaccinations, had no specified protocol deviations, and were SARS-CoV-2 negative on the D21 visit. Primary COVID-19 endpoints were as in ref.[1]: RT-PCR–confirmed symptomatic COVID-19 occurring at least 7 days after dose two. As all breakthrough cases occurred in the United States in Part A as of the efficacy data cut date, the case-cohort set (Panel A) was restricted to the U.S. cohort and therefore all primary COVID-19 endpoints included in the analysis (Panel B) were in the U.S.

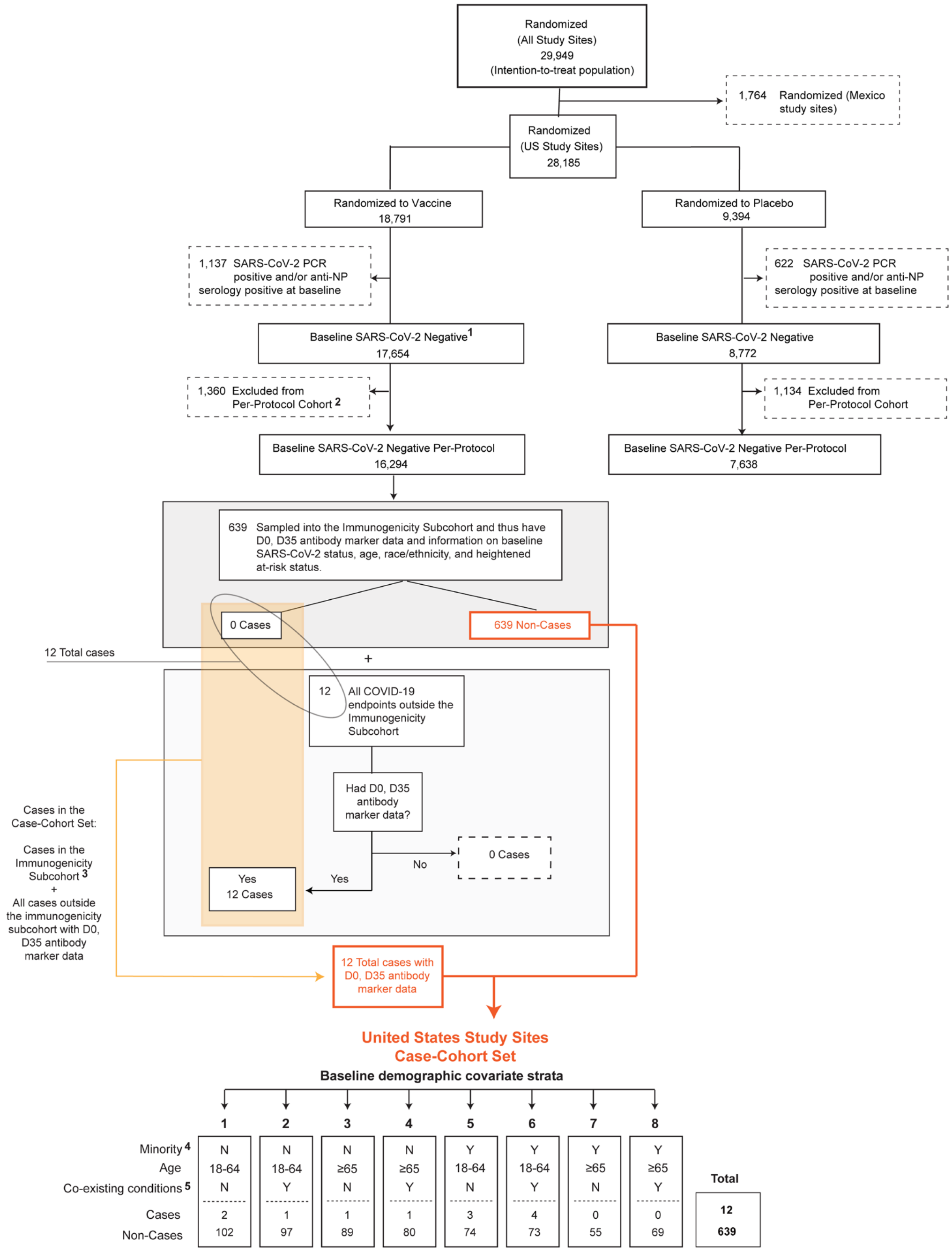


1 Participants with missing baseline SARS-CoV-2 RT-PCR or anti-NP (nucleocapsid protein) serology data were considered baseline negative for the corresponding assay. After this, "baseline SARS-CoV-2 negative" was defined as baseline SARS-CoV-2 RT-PCR negative **and** baseline anti-NP serology negative.
2 Reasons for exclusion from per-protocol included: Did not receive two doses, doses out of allowed window, major protocol deviation.
3 There were no cases in the immunogenicity subcohort.
4 Minority includes Blacks or African Americans, Hispanics or Latinos, American Indians or Alaska Natives, Native Hawaiians, and other Pacific Islanders. Non-Minority includes all other races with observed race (Asian, Multiracial, White, Other) and observed ethnicity Not Hispanic or Latino. Therefore Unknown and Not reported have missing values for this.
5 Co-existing conditions are the same as those listed in Table 1 of Dunkle et al. NEJM 2022: obesity (defined as a body-mass index [the weight in kilograms divided by the square of the height in meters] of ≥30.0), chronic lung disease, diabetes mellitus type 2, cardiovascular disease, or chronic kidney disease.

**Supplementary Figure 2. Flowchart of study participants from randomization through membership in the baseline SARS-CoV-2 negative per-protocol case-cohort set (U.S. study sites).** Membership in the case-cohort set required availability of D0 and D35 antibody data and no evidence of SARS-CoV-2 infection through 6 days post D35. Antibody data from the placebo arm are not used in correlates analyses, given no variability in values; they were only used to verify low false positive rates of the immunoassays.

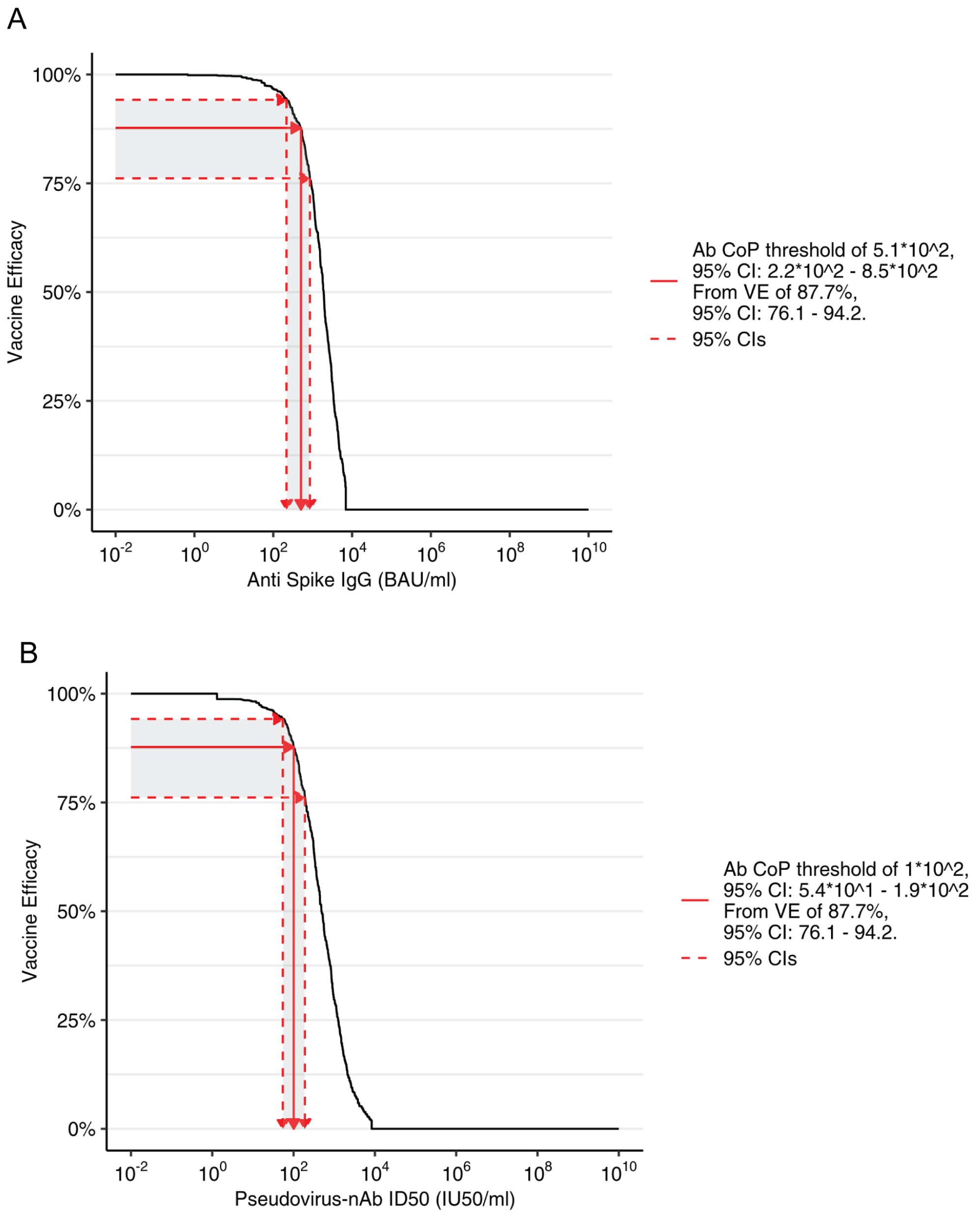


**Supplementary Figure 3. Inverse probability sampling (IPS)-weighted empirical reverse cumulative distribution function curves for each Day 35 marker (spike IgG, nAb ID50) and application of the Siber (2007) method[2] for estimating a threshold of perfect vs. no protection.**

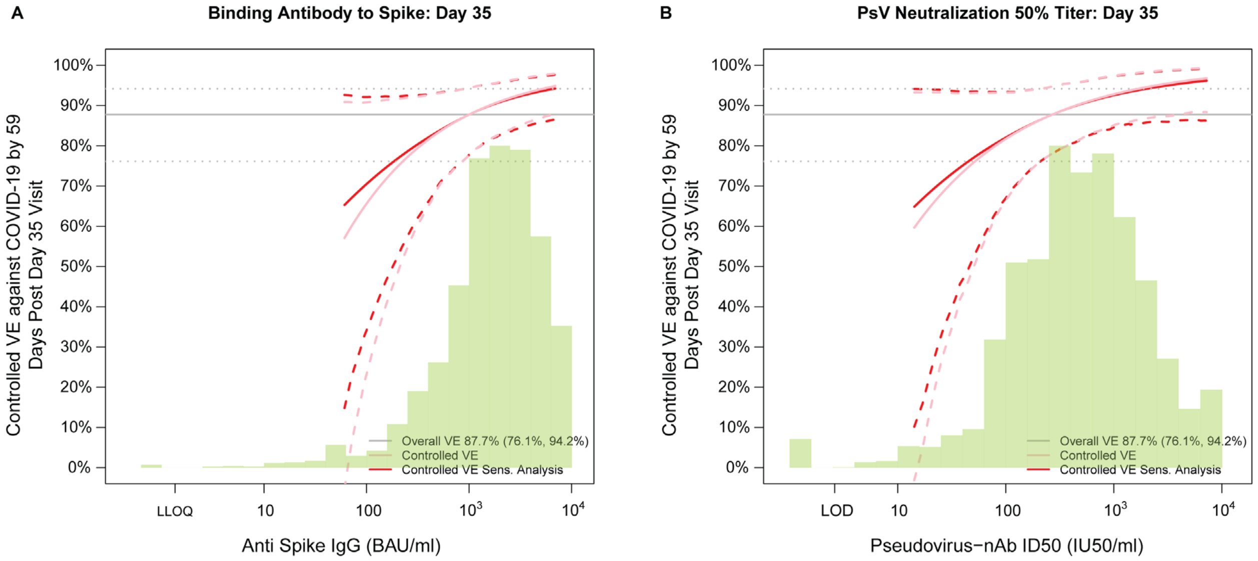


**Supplementary Figure 4. Vaccine efficacy in the US study population with causal sensitivity analysis by Day 35 (A) anti-spike IgG concentration or (B) pseudovirus (PsV)-nAb ID50 titer.** Vaccine efficacy estimates were obtained using the method of Gilbert et al.[3] The green histogram is an estimate of the density of D35 antibody marker level and the horizontal gray line is the overall vaccine efficacy from 7 to 59 days post D35, with the dotted gray lines indicating the 95% confidence intervals (this number 87.7% differs from the 90.4% reported in ref.[1], which was based on counting COVID-19 endpoints starting 7 days post D35 in both the US and Mexico study sites). The pink solid line is point estimates assuming no unmeasured confounding; the dashed lines are bootstrap point-wise 95% CIs. The red solid line is point estimates assuming unmeasured confounding in a sensitivity analysis (dashed lines are bootstrap point-wise 95% CIs); see the Statistical Analysis Plan for details. LLOQ, lower limit of quantitation; LOD, (lower) limit of detection. Analyses adjusted for baseline risk score.

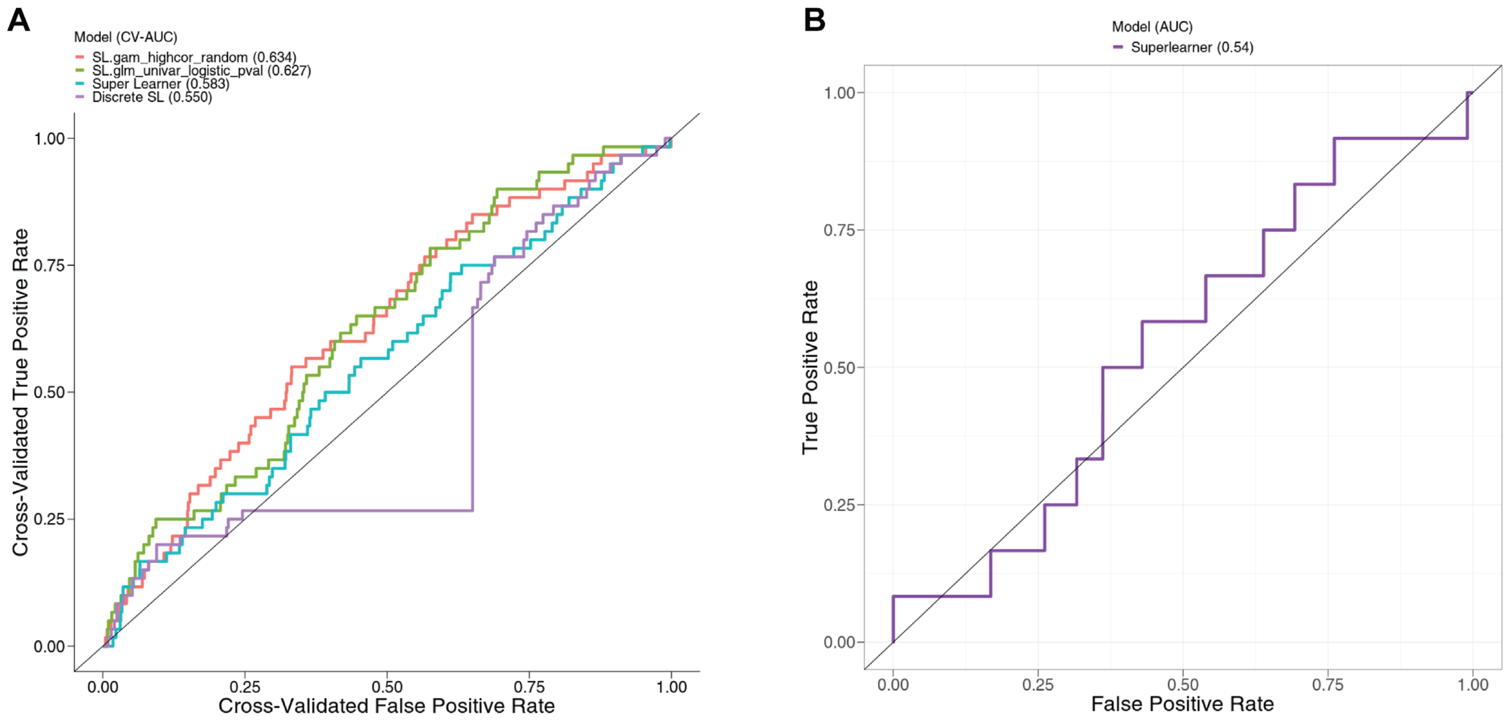


**Supplementary Figure 5. Performance of the baseline risk score built from ensemble machine learning**. (A) Receiver operating characteristic (ROC) curves based on cross-validated (CV)-estimated predicted probabilities for the top two learners, Superlearner and Discrete Superlearner. CV-estimated predicted probabilities were computed using only data from the placebo arm in the US. (B) ROC curve based on Superlearner predicted probabilities in vaccine recipients.

**Supplementary Table 1. Sample sizes of baseline SARS-CoV-2 negative per-protocol vaccine recipients included in the case-cohort set included in immune correlates analyses (U.S. study sites), by baseline sampling strata and case/non-case strata.**

Case-cohort set = Baseline SARS-CoV-2 negative per-protocol vaccine recipients included in D35 marker correlates analysis [in the immunogenicity subcohort (IS) and/or a breakthrough COVID-19 case)]*

| | Baseline Sampling Strata of Baseline SARS-CoV-2 Negative Per-Protocol Vaccine Participants Included in Correlates Analyses | | | | | | | | |
|---|---|---|---|---|---|---|---|---|---|
| | 1 | 2 | 3 | 4 | 5 | 6 | 7 | 8 | Total |
| Breakthrough COVID-19 cases (both within and outside the IS*) with D0, D35 Ab marker data | 2 | 1 | 1 | 1 | 3 | 4 | 0 | 0 | 12 |
| Non-cases in the IS with D0, D35 Ab marker data | 102 | 97 | 89 | 80 | 74 | 73 | 55 | 69 | 639 |

Ab, antibody; IS, immunogenicity subcohort

Demographic covariate strata:
1. U.S. White Non-Hispanic**, age 18-64, No coexisting conditions***
2. U.S. White Non-Hispanic, age 18-64, Coexisting conditions
3. U.S. White Non-Hispanic, age ≥ 65, No coexisting conditions
4. U.S. White Non-Hispanic, age ≥ 65, Coexisting conditions
5. U.S. Minority, age 18-64, No coexisting conditions
6. U.S. Minority, age 18-64, Coexisting conditions
7. U.S. Minority, age ≥ 65, No coexisting conditions
8. U.S. Minority, age ≥ 65, Coexisting conditions

Cases are baseline SARS-CoV-2 negative per-protocol vaccine recipients with the primary COVID-19 endpoint starting 7 days post D35 visit through to the data cut (April 19, 2021).
Non-cases/Controls are baseline negative per-protocol vaccine recipients sampled into the immunogenicity subcohort with no evidence of SARS-CoV-2 infection up to the end of the correlates study period (the data cut-off date April 19, 2021). IS membership required availability of D0 and D35 antibody data and no evidence of SARS-CoV-2 infection through 6 days post D35.

*All breakthrough COVID-19 cases with D0, D35 Ab marker data were outside the IS

** White Non-Hispanic is defined as Race=White and Ethnicity=Not Hispanic or Latino. All other Race subgroups are defined as Black, Asian, American Indian or Alaska Native, Native Hawaiian or Other Pacific Islander, Multiracial, Other, Not reported, or Unknown.
Minority is defined as the complement of being known to be White Non-Hispanic.

***Coexisting conditions are the same as those listed in Table 1 of Dunkle et al.[1]: obesity (defined as a body-mass index [the weight in kilograms divided by the square of the height in meters] of ≥30.0), chronic lung disease, diabetes mellitus type 2, cardiovascular disease, or chronic kidney disease.

**Supplementary Table 2. PREVENT-19 U.S. cohort demographic and clinical characteristics at enrollment in the baseline SARS-CoV-2 negative per-protocol immunogenicity subcohort***

| Characteristics | Vaccine (N = 669*) | Placebo (N = 76) | Total (N = 745) |
|---|---|---|---|
| **Age** | | | |
| Age 18-64 | 355 (53.1%) | 42 (55.3%) | 397 (53.3%) |
| Age ≥ 65 | 314 (46.9%) | 34 (44.7%) | 348 (46.7%) |
| Mean (Range) | 55.0 (18.0, 86.0) | 54.6 (22.0, 80.0) | 55.0 (18.0, 86.0) |
| **Coexisting Conditions**** | | | |
| Yes | 329 (49.2%) | 41 (53.9%) | 370 (49.7%) |
| No | 340 (50.8%) | 35 (46.1%) | 375 (50.3%) |
| **Age, Coexisting Conditions** | | | |
| Age 18-64 Coexisting conditions | 174 (26.0%) | 22 (28.9%) | 196 (26.3%) |
| Age 18-64 No coexisting conditions | 181 (27.1%) | 20 (26.3%) | 201 (27.0%) |
| Age ≥ 65 | 314 (46.9%) | 34 (44.7%) | 348 (46.7%) |
| **Sex** | | | |
| Female | 309 (46.2%) | 39 (51.3%) | 348 (46.7%) |
| Male | 360 (53.8%) | 37 (48.7%) | 397 (53.3%) |
| **Hispanic or Latino Ethnicity** | | | |
| Hispanic or Latino | 143 (21.4%) | 12 (15.8%) | 155 (20.8%) |
| Not Hispanic or Latino | 521 (77.9%) | 64 (84.2%) | 585 (78.5%) |
| Not reported and unknown | 5 (0.7%) | 0 (0.0%) | 5 (0.7%) |
| **Race** | | | |
| White | 453 (67.7%) | 47 (61.8%) | 500 (67.1%) |
| Black or African American | 127 (19.0%) | 19 (25.0%) | 146 (19.6%) |
| Asian | 46 (6.9%) | 6 (7.9%) | 52 (7.0%) |
| American Indian or Alaska Native | 18 (2.7%) | 1 (1.3%) | 19 (2.6%) |
| Native Hawaiian or Other Pacific Islander | 1 (0.1%) | 0 (0.0%) | 1 (0.1%) |
| Multiracial | 12 (1.8%) | 1 (1.3%) | 13 (1.7%) |
| Not reported and unknown | 12 (1.8%) | 2 (2.6%) | 14 (1.9%) |

*Of the 669 sampled vaccine recipients, 639 had antibody marker data measured at D0 and D35 and had no evidence of SARS-CoV-2 infection through 6 days post D35 and hence are in the immunogenicity subcohort.

** Coexisting conditions are the same as those listed in Table 1 of Dunkle et al.[1]: obesity (defined as a body-mass index [the weight in kilograms divided by the square of the height in meters] of ≥30.0), chronic lung disease, diabetes mellitus type 2, cardiovascular disease, or chronic kidney disease.

**Supplementary Table 3. Distribution of variants among primary COVID-19 endpoints in PREVENT-19 starting 7 days post D35 through to the data cut-off (April 19, 2021) (no primary endpoints were with the ancestral/Wuhan-Hu-1 strain) with available sequence data.**

| Variant (PANGO lineage)* | **CDC and Prevention Classification (June 2021)** | **Placebo (n=37)** | Vaccine (n=7) |
|---|---|---|---|
| B.1 | Wuhan Ancestral lineage | 2 | 1 |
| B.1.1 | Wuhan Ancestral lineage | 1 | 0 |
| B.1.1.519** | Wuhan Ancestral lineage | 1 | 0 |
| B.1.1.7 (Alpha) | VoC | 21 | 3 |
| B.1.2 | Wuhan Ancestral lineage | 2 | 0 |
| B.1.311 | Wuhan Ancestral lineage | 1 | 0 |
| B.1.351 (Beta) | VoC | 0 | 1 |
| B.1.526 (Iota) | VoI | 2 | 2 |
| B.1.596 | Wuhan Ancestral lineage | 1 | 0 |
| **B.1.617.1 (Kappa)** | VoI | 1 | 0 |
| **B.1.623** | Wuhan Ancestral lineage | 1 | 0 |
| **B.1.637** | Wuhan Ancestral lineage | 1 | 0 |
| **P.1 (Gamma)** | VoC | 2 | 0 |
| **P.2 (Zeta)** | VoI | 1 | 0 |

*Variants without a Greek letter are of the Wuhan Ancestral lineage and have never been classified by the Center for Disease Control and Prevention as a variant of interest (VoI) or as a variant of concern (VoC).
**Formerly monitored variant never classified as a VoI or VoC.

**Supplementary Table 4. D35 antibody marker response rates and geometric means in the U.S. cohort by COVID-19 outcome status.** Analysis based on baseline SARS-CoV-2 negative per-protocol placebo recipients in the case-cohort set. Median (interquartile range) days from vaccination to D35 was 38 (5).

| | Placebo COVID-19 Cases[1] | | | Placebo Non-Cases in Immunogenicity Subcohort[2] | | | Comparison | |
|---|---|---|---|---|---|---|---|---|
| D35 Marker | N | Proportion with Antibody Response[3] (95% CI) | Geometric Mean (GM) (95% CI) | N | Proportion with Antibody Response[3] (95% CI) | Geometric Mean (GM) (95 % CI) | Response Rate Difference (Non-Cases – Cases) | Ratio of GM (Non-Cases/ Cases) |
| Anti Spike IgG (BAU/ml) | 41 | 7.3% (2.3%, 21.0%) | 1.19 (0.89, 1.59) | 72 | 0.9% (0.2%, 3.5%) | 0.82 (0.70, 0.95) | 0.2% (0%, 1.7%) | 0.68 (0.50, 0.95) |
| Pseudovirus-nAb ID50 (IU50/ml) | 41 | 0.0% (0.0%, 0.0%) | 1.31 (1.31, 1.31) | 72 | 0.2% (0.0%, 1.7%) | 1.32 (1.29, 1.35) | -6.4% (-20.1%, -0.8%) | 1.0 (0.99, 1.03) |

[1]Cases are baseline SARS-CoV-2 negative per-protocol placebo recipients with the primary COVID-19 endpoint (symptomatic RT-PCR-confirmed COVID-19) starting 7 days post D35 visit through to the efficacy data cut-off date (April 19, 2021).

[2]Non-cases are are baseline negative per-protocol placebo recipients sampled into the immunogenicity subcohort with no evidence of SARS-CoV-2 infection up to the end of the correlates study period (the data cut-off date April 19, 2021).

[3]Antibody response defined by Spike IgG concentration above the antigen-specific positivity cut-off (10.8424 BAU/ml) or by detectable ID50 > limit of detection (LOD) = 2.612 IU50/ml.

**Supplementary Table 5. Assay limits of the two antibody markers evaluated as immune correlates.** BAU = binding antibody units; IU = International Units; LLOQ, lower limit of quantitation; ULOQ, upper limit of quantitation.

| MSD Binding Assay (Nexelis) (Spike IgG marker) | |
|---|---|
| Reported units | BAU/ml |
| | Spike |
| Positivity Cutoff | 10.8424 |
| LOD | 0.3076 |
| ULOD | 172,226.2 |
| LLOQ | 1.35 |
| ULOQ | 6934 |
| All values < LLOQ were set to LLOQ/2 | |
| All values > ULOQ were set to ULOQ (for immune correlates analyses) | |
| **Pseudovirus neutralization titer (Monogram) (nAb ID50 marker)** | |
| Reported units | IU50/ml |
| LOD* | 2.612 |
| LLOQ | 3.3303 |
| ULOQ | 8319.938 |
| Values < LOD are denoted as undetectable responses and were set to LOD/2 | |
| All values > ULOQ were set to ULOQ (for immune correlates analyses) | |

*The limit of detection (LOD) was not formally defined; we denote the value corresponding to the starting dilution level of the assay as the LOD.
