## Supplementary material for "Immune Correlates Analysis of the PREVENT-19 COVID-19 Vaccine Efficacy Clinical Trial": Statistical Analysis Plan

### Statistical Analysis Plan for Assessing Immune Correlates in the Coronavirus Efficacy (PREVENT-19) Phase 3 Trial of the NVX-CoV2373 COVID-19 Vaccine

USG COVID-19 Response Team / Coronavirus Prevention Network (CoVPN) Biostatistics Team

Peter B. Gilbert[1,2*], Youyi Fong[1,2], David Benkeser[3], Jessica Andriesen[1], Bhavesh Borate[1], Marco Carone[2], Lindsay N. Carpp[1], Iván Díaz[4], Michael P. Fay[5], Andrew Fiore-Gartland[1], Nima S. Hejazi[6], Ying Huang[1,2], Yunda Huang[1], Ollivier Hyrien[1], Holly E. Janes[1,2], Michal Juraska[1], Alex Luedtke[7], Martha Nason[5], April K. Randhawa[1], Lars van der Laan[6], Brian D. Williamson[1], Dean Follmann[5]

[1]Vaccine and Infectious Disease and Public Health Sciences Divisions, Fred Hutchinson Cancer Research Center, Seattle, Washington

[2]Department of Biostatistics, University of Washington, Seattle, Washington

[3]Department of Biostatistics and Bioinformatics, Rollins School of Public Health, Emory University, Atlanta, Georgia

[4]Department of Population Health Sciences, Weill Cornell Medical College, New York, New York

[5]National Institute of Allergy and Infectious Diseases, Bethesda, Maryland

[6]Division of Biostatistics, School of Public Health, University of California, Berkeley, California

[7]Department of Statistics, University of Washington, Seattle, Washington

June 22, 2022

Contents

List of Tables 4

List of Figures 5

1 Introduction 6

2 Antibody Assays and and Day 35 Markers 7

3 Study Cohorts and Endpoints 10
- 6.2 Methods for Positive Response Calls for bAb and nAb Assays 22
- 6.3 SARS-CoV-2 Antigen Targets Used for bAb and nAb Markers 22

7 Baseline Risk Score (Proxy for SARS-CoV-2 Exposure) 22

8 Correlates Analysis Descriptive Tables by Case/Non-Case Status 24

9 Correlates of Risk Analysis Plan 25

**12 Estimating a Threshold of Protection Based on an Established or Putative CoP (Population-Based CoP) 45**

**13 Considerations for Baseline SARS-CoV-2 Positive Study Participants 46**

**14 Avoiding Bias with Pseudovirus Neutralization Analysis due to Use of Anti-HIV Antiretroviral Drugs 47**

**15 Novavax Binary Principal Stratification Results 55**

5 Two-stage correlates analysis. Stage 1 consists of analyses of Day 35 markers as correlates of risk and of protection of the primary endpoint and potentially also of some secondary endpoints, and includes antibody marker data from all COVID and SARS-CoV-2 infection cases (COV-INF) through to the time of the data lock for the first correlates analyses. Stage 2 consists of analyses of Day 35 markers as correlates of risk and of protection of longer term endpoints and analyses of longitudinal markers as outcome-proximal correlates of risk and of protection, and includes antibody marker data from all subsequent COVID and COV-INF cases. Stage 1 measures Day 1, Day 35 antibody markers and COV-INF and COVID diagnosis time point markers; Stage 2 measures antibody markers from all sampling time points and COV-INF plus COVID diagnosis sampling time points not yet assayed. The same immunogenicity subcohort is used for both stages. . . . . . . . . . . . . . . . 51

#### 1 Introduction

This SAP describes the statistical analysis of antibody markers measured at Day 35 as immune correlates of risk and as immune correlates of protection against the COVID primary endpoint in the Coronavirus Efficacy (PREVENT-19) phase 3 trial of the NVX-CoV2373 COVID-19 vaccine. In this trial, estimated efficacy of the NVX-CoV2373 vaccine against symptomatic COVID illness was 90.4% (95% confidence interval, 82.9 to 94.6%) [?]. Some key facts about this trial:

- There are sister trials in UK and South Africa; this SAP restricts to data from the US and Mexico phase 3 trial
- Two doses regimen at Day 0 and 21. Primary efficacy analysis starts at 7 days post dose 2 (second dose at Day 21). Markers measured at Day 35.
- Enrollment occurred between December 27, 2020, and February 18, 2021. Final analysis cutoff date is April 19, 2021.
- Per protocol means receiving two doses, baseline negative ( 6.5% baseline positive, where positive means either serology or nucleic acid positive), and being at risk at Day 28. Full analysis set means receiving one dose. 2:1 randomization ratio
- In PP population, a total of 77 cases (mild, moderate or severe) are observed, 14 in the vaccine arm and 63 in the placebo arm (Dunkle et al.).
- 31 of 77 cases are Alpha.
- COVID is classified into mild, moderate and severe. Mild requires at least two from a list of symptoms. Mild and moderate in PREVENT-19 roughly correspond to non-severe in COVE, which requires two symptoms, and moderate but not mild in ENSEMBLE, which require two and one symptom, respectively.
- Study include 6 sites in Mexico and 113 sites in US. In the primary analysis, all vaccine cases are in US and one placebo case is in Mexico.

For correlates all cases are in the US such that only US sites are included.

#### 2 Antibody Assays and and Day 35 Markers

The antibody markers of interest are measured using two different humoral immunogenicity assays [more detail on assay type (2) can be found in Sholukh et al. (2020)]:

(1) **bAbs**: **Binding antibodies** to the vaccine insert SARS-CoV-2 proteins;

and (2) **Pseudovirus-nAbs**: **Neutralizing antibodies** against viruses **pseudotyped** with the vaccine insert SARS-CoV-2 proteins.

The Supplementary text in the article provides details of the assays. We include the necessary statistical details below.

(1) **bAb assay**: The MSD-ECL Multiplex Assay (MSD-ECL = meso scale discovery-electrochemiluminescence assay).

The MSD assay measures binding antibody to antigens corresponding to: Spike (an engineered version of the Spike protein harboring a double proline substitution (S-2P) that stabilizes it in the closed, prefusion conformation [McCallum et al. (2020)]); the Receptor Binding Domain (RBD) of the Spike protein; and Nucleocapsid protein (N), which is not contained in any of the COVID-19 vaccines.

The bAb assay readouts are in units AU/ml, where AU stands for arbitrary units from a standard curve. The process of validating the assay defined a lower limit of detection (LOD), an upper limit of detection (ULOD), a lower limit of quantitation (LLOQ), an upper limit of quantitation (ULOQ), and a positivity cut-off for each antigen that defines positive vs. negative response. These values are as follows:

- bAb Spike:
  - Pos. Cutoff = 1204.711 AU/ml
  - LLOQ = 150.4 AU/ml
  - ULOQ = 770,464.6 AU/ml

– LOD = N/A

– ULOD = N/A

The Vaccine Research Center established factors for bridging the MSD assay readouts from AU/ml to Binding Antibody Units/ml (BAU/ml), based on bridging to the WHO International Standard for anti-SARS-CoV-2 immunoglobulin. For the three binding antibody variables CoV-2 Spike IgG, CoV-2 RBD IgG, and CoV-2 N IgG, these conversion factors are 0.0090, 0.0272, and 0.0024, respectively. These conversion factors are applied, such that all binding Ab readouts are reported in BAU/ml, for all analyses. These conversion factors are also applied to yield the LOD, ULOD, LLOQ, and ULOQ on the WHO IU/ml scale. The following shows the assay limits on the BAU/ml scale:

- bAb Spike:
  - Pos. Cutoff = 10.8424 BAU/ml
  - LOD = 0.3076 BAU/ml
  - ULOD = 172,226.2 BAU/ml
  - LLOQ = 1.35 BAU/ml
  - ULOQ = 6934 BAU/ml

All values below the LLOQ are assigned the value LLOQ/2. A positive response is defined by value above the Pos. Cutoff. For immunogenicity reporting, values greater than the ULOQ are not given a ceiling value of the ULOQ, the actual readouts are used. For the immune correlates analyses, values greater than the ULOQ are assigned the value of the ULOQ.

(2) **Pseudovirus-nAb assay**: A firefly luciferase (ffLuc) reporter neutralization assay for measuring neutralizing antibodies against SARS-CoV-2 Spike-pseudotyped viruses.

Based on the assay in the Monogram lab, serum inhibitory dilution 50% titer (ID50) values are estimated based on a starting serum dilution of 1:40, with a total of ten 3-fold dilutions. (Each sample is diluted initially at 1:20, then diluted serially 3-fold for a total of 10 concentrations. The starting dilution of 1:20 is reported as 1:40 after addition of the virus.) So, the dilution series is 1:40 to 1:787,320 (= 40 * 39). Thus 1:40 is the LOD on the scale of the assay. The process of validating the assay defined the LOD, LLOQ, and ULOQ for ID50 as follows:

- ID50:
  - LOD = 40
  - LLOQ = 51
  - ULOQ = 127411

ID50 values below the LOD are assigned the value LOD/2 = 40/2 = 20. For immunogenicity reporting, values greater than the ULOQ are not given a ceiling value of the ULOQ, the actual readouts are used. For the immune correlates analyses, values greater than the ULOQ are assigned the value of the ULOQ.

ID50 values are reported in international units with the following calibration factor, defined using the D614G strain in the assay:

- Calibration factor ID50: 0.0653

The original readouts are calibrated to the IU scale by multiplying each original ID50 value by 0.0653 (See Feng et al. Table 2 and Gilbert et al. Supplementary Material), and units are reported in international units as IU50/ml for ID50. Consequently, the LOD, LLOQ and ULOQ for IU50/ml are as follows in International Units:

- IU50/ml:
  - LOD = 2.612
  - LLOQ = 3.3303
  - ULOQ = 8319.938

Therefore the lowest possible value of ID50 readouts on the log10 scale is log10(2.612/2) = 0.116. Positivity cutoff is assigned LOD.

Based on each immunoassay applied to serum samples collected from participants on Day 1 (baseline, first dose of vaccination visit)and Day 35 (post-vaccination visit), the following set of antibody markers was defined for immunogenicity and immune correlates analyses.

- For bAb: $\log_{10}$ IgG concentration (BAU/ml) at each time point, and the difference in $\log_{10}$ concentration (Day 35 minus Day 1) representing $\log_{10}$ fold-rise in IgG concentration from baseline to 14 days post dose two. These markers are defined for Spike.
- For PsV nAb: $\log_{10}$ serum inhibitory dilution 50% titer (ID50 in IU50/ml) at each time point, as well as the $\log_{10}$ fold-rise of these markers over Day 1 to Day 35.

### 3 Study Cohorts and Endpoints

#### 3.1 Study Cohort for Correlates Analyses

The analysis cohort for the correlates analysis is baseline SARS-CoV-2 negative participants in the per-protocol cohort, with the per-protocol cohort defined as those who received both planned vaccinations without any specified protocol deviations, and who were SARS-CoV-2 negative at the terminal vaccination visit. We refer to this cohort representing the primary population for correlates analysis as the Per-Protocol Baseline Negative Cohort.

As the primary analysis of vaccine efficacy is conducted in baseline negative individuals, correlates of risk (CoR) and correlates of protection (CoP) analyses are only done in baseline negative individuals, and the analysis of data from baseline positive individuals is for purposes of immunogenicity characterization, given too-few anticipated vaccine breakthrough study endpoints for CoR/CoP assessment (although if there are many baseline positive vaccine breakthrough endpoint cases that baseline positive subgroup analyses may be considered). In baseline negative individuals, antibody marker data in placebo recipients is relevant for verifying the expectation that almost all Day 35 marker responses will be negative, given the lack of SARS-CoV-2 antigen exposure.

##### 3.2 Study Endpoints

Endpoints for correlates analyses of Day 35 markers are included if they occur at least 7 days after the Day 35 visit, to help ensure that the endpoint did not occur prior to Day 35 antibody measurement.

Figure 2 defines five study endpoints assessed in COVID-19 vaccine efficacy trials, where COVID (symptomatic infection) is used as the primary endpoint in the PREVENT-19 trial. Only the COVID endpoint is assessed in the current manuscript. For the correlates analysis, all available follow-up for participants is included through to the time of the data base lock for the correlates analysis, for every CoR and CoP analysis that is conducted. This means that the time of right censoring for a given failure time endpoint is the first event of loss to follow-up or the date of administrative censoring defined as the last date of available follow-up. For CoP analyses, which use both vaccine and placebo recipient data and leverage the randomization, follow-up is censored at the time of unblinding. In general for the current manuscript all blinded follow-up is included and no post-unblinding follow-up is included.

#### 4 Objectives of Immune Correlates Analyses of a Phase 3 Trial Data Set

##### 4.1 Correlates of Risk and Correlates of Protection

We broadly classify the proposed analyses into two related categories: correlates of risk (CoR) and correlates of protection (CoP) analyses. CoR analyses seek to characterize correlations/associations of markers with future risk of the outcome amongst vaccinated individuals in the study cohort. CoP analyses seek to formally characterize causal relationships among vaccination, antibody markers and the study endpoint, and use data from both vaccine and placebo recipients. Table 1 summarizes these objectives and statistical frameworks that are commonly used to these ends.

The advantage of CoR analyses it that it is possible to obtain definitive answers from the phase 3 data sets, that is one can credibly characterize associations between markers and outcome. The advantage of CoP analyses is that the effects being estimated have interpretation directly in terms of how an antibody marker can be used to reliably predict vaccine efficacy (the criterion for use of a non-validated surrogate endpoint for accelerated approval, Fleming and Powers, 2012). The disadvantage of CoR analyses are that a CoR may fail to be a CoP, for example due to unmeasured confounding, lack of transitivity where a vaccine effect on an antibody marker occurs in different individuals than clinical vaccine efficacy, or off-target effects (VanderWeele, 2013). The disadvantage of CoP analyses is that statistical inferences rely on causal assumptions that cannot be completely verified from the phase 3 data, such that compelling evidence may require multiple phase 3 trials and external evidence on mechanism of protection (e.g., from adoptive transfer or vaccine challenge trials). Our approach presents results for both CoR and CoP analyses, seeking clear exposition of how to interpret results, the assumptions undergirding the validity of the results, and diagnostics of these assumptions and assessment of robustness of findings to violation of assumptions.

Table 1: Correlates of Risk (CoRs) and Correlates of Protection (CoPs) Objectives for Day 35 Markers

| Objective Type | Objective |
|---|---|
| **CoRs (Risk Prediction Modeling)** | **To assess Day 35 markers as CoRs in vaccine recipients**<br>a. Relative risks of outcome across marker levels<br>b. Absolute risk of outcome across marker levels<br>c. Machine learning risk prediction for multivariable markers |
| **CoP: Correlates of VE** | **To assess Day 35 markers as correlates of VE in vaccine recipients**<br>a. Principal stratification effect modification analysis<br>b. Assesses VE across subgroups of vaccine recipients defined by Day 35 marker level in vaccine recipients |
| **CoP: Controlled Effects on Risk and VE** | **To assess Day 35 markers for how assignment to vaccine and a fixed marker value would alter risk compared to assignment to placebo** |
| **CoP: Stochastic Interventional Effects on Risk and VE** | **To assess Day 35 markers for how stochastic shifts in their distribution would alter mean risk and VE** (Hejazi et al., 2020) |
| **CoP: Mediators of VE** | **To assess Day 35 markers as mediators of VE**<br>a. Mechanisms of protection via natural direct and indirect effects<br>a. Estimate the proportion of VE mediated by a marker or markers |

Because there are only 12 vaccine breakthrough COVID-19 endpoints with D35 antibody data available for correlates analyses, only a subset of the statistical methods were applied. In particular only the CoR analyses a. and b., the CoR nonparametric threshold analyses, and the CoP Controlled Effects on Risk and VE through marginalized Cox modeling are applied.

#### 5 Case-cohort Sampling Design for Measuring Antibody Markers

Figure 4 illustrates the case-cohort (Prentice, 1986) sampling design that is used for measuring Day 1, 35 antibody markers in a random sample of trial participants. The random sample is stratified by the key baseline covariates: assigned randomization arm, baseline SARS-CoV-2 status (negative means negative in both the anti-NP binding antibody assay and the nasal swab RT-

PCR assay; positive means positive in either assay, Dunkle et al., 2021), and demographics strata (8 US strata formed by Age 18-64 or $\geq$ 65, Underrepresented Minority: Yes vs. No /Unknown, and Coexisting conditions: Yes vs. No, and 2 Mexico strata, Age 18-64 or $\geq$ 65). Because the design uses a stratified random sample instead of the simple random sample proposed by Prentice (1986), the design may also be referred to as a "two-phase sampling design" (Breslow et al., 2009b,a), where "phase one" refers to variables measured in all participants and "phase two" refers to variables only measured in a subset (thus the "case-cohort sample" constitutes the phase-two data).

The case-cohort design enables obtaining marker data (Day 1, 35) for the immunogenicity subcohort during early trial follow-up in real-time batches, thereby accelerating the time until final data set creation and hence data analysis and results on Day 35 marker correlates. The design allows using the same immunogenicity subcohort to assess correlates for multiple endpoints, relevant for the COVID-19 VE trials with multiple endpoints (Figure 2). This makes the design operationally simpler than a case-control sampling design.

##### 5.1 Immunogenicity subcohort

The immunogenicity subcohort was sampled from the subset of participants in the Full Analysis Set (FAS) cohort used in the primary analysis of vaccine efficacy against the primary endpoint (with the FAS defined as all randomized participants who received at least one dose of investigational product) for whom <u>all</u> of the following information was available: baseline SARS-CoV-2 status; age, race/ethnicity (needed to define Minority status as described below), and presence of coexisting conditions associated with high risk of severe COVID-19; <u>and</u> Day 1 and Day 35 samples collected.

Figure 1 summarizes the planned size of the immunogenicity subcohort, by the baseline factors used to stratify the random sampling. The subcohort sampling is implemented to create representative sampling across the entire period of enrollment.

For the sampling, Minority includes Blacks or African Americans, Hispanics or Latinos, American Indians or Alaska Natives, Native Hawaiians, and other

| | Numbers of Participants Sampled Into **40** Strata (Total N=**1620**) | | | |
|---|---|---|---|---|
| Baseline SARS-CoV-2 Status[1] | Negative | | Positive | |
| Covariate Strata[2] | 1-8 (US) | 9-10 (Mex) | 11-18 (US) | 19-20 (Mex) |
| Vaccine | 100 each | 50 each | 35 each | 13 each |
| Placebo | 12 each | 6 each | 35 each | 13 each |

[1] Baseline SARS-CoV-2 status as defined in the SAP, i.e., negative means negative in both the anti-NP binding antibody assay and the nasal swab RT-PCR assay; positive means positive in either assay. Missing values from either assay are assigned negative, to be consistent with the primary efficacy analysis.

[2] Strata 1-20 subdivide the vaccine arm and strata 21-40 subdivide the placebo arm. Take the vaccine arm as example. Strata 1-10 subdivide the baseline negative population. They include 8 US strata formed by dividing the baseline negative according to (Age 18-64 or >= 65), (Underrepresented Minority: Yes vs. No /Unknown), and (Comorbidity: Yes vs. No), and 2 Mexico strata, (Age 18-64 or >= 65). Strata 11-20 correspondingly subdivide the baseline positive population.

Figure 1: Planned Immunogenicity Subcohort Sample Sizes by Baseline Strata for Antibody Marker Measurement

Pacific Islanders. Non-Minority includes all other races with observed race (Asian, Multiracial, White, Other) and observed ethnicity Not Hispanic or Latino. Therefore Unknown and Not reported have missing values for this sampling stratum variable.

Coexisting conditions refers to participants having coexisting conditions associated with high risk of severe COVID-19 illness, with co-existing conditions defined in Dunkle et al. (2021). (obesity, chronic lung disease, diabetes mellitus type 2, cardiovascular disease, and/or chronic kidney disease)

### 6 Unsupervised Feature Engineering of Antibody Markers (Stage 1: Day 1, 35)

#### 6.1 Descriptive Tables and Graphics

##### 6.1.1 Antibody marker data

Binding antibody titers to full length SARS-CoV-2 Spike protein is measured in all participants in the immunogenicity subcohort (augmented with COVID-19 endpoint cases). Binding antibody IgG Spike, as well as fold-rise in this marker from baseline, are measured at each pre-defined time point. Indicators of 2-fold rise and 4-fold rise in IgG concentration (fold rise [post/pre] $\geq 2$ and $\geq 4$, 2FR and 4FR) are measured at each pre-defined post-vaccination timepoint. Binding antibody responders to a given antigen at each pre-defined timepoint are defined as participants with value above the antigen-specific positivity cut-off. Binding antibody IgG 2FR (4FR) at each pre-defined timepoint to a given antigen are defined as participants who had baseline values below the LLOQ with IgG concentration at least 2 times (4 times) above the assay LLOQ, or as participants with baseline values above the LLOQ with at least a 2-fold (4-fold) increase in IgG concentration.

Pseudovirus neutralizing antibody ID50 titers, as well as fold-rise in ID50 titers from baseline, are measured at each pre-defined time point. Indicators of 2-fold rise and 4-fold rise in ID50 titer (fold rise [post/pre] $\geq 2$ and $\geq 4$, 2FR and 4FR) are measured at each pre-defined post-vaccination timepoint. Neutralization responders at each pre-defined timepoint are defined as participants who had baseline values below the LOD with detectable ID50 neutralization titer above the assay LOD, or as participants with baseline values above the LOD with a 4-fold increase in neutralizing antibody titer. Neutralization 2FR (4FR) at each pre-defined timepoint are defined as participants who had baseline values below the LLOQ with ID50 at least 2 times (4 times) above the assay LLOQ, or as participants with baseline values above the LLOQ with at least a 2-fold (4-fold) increase in neutralizing antibody titer.

Note that for defining positive response, 2FR, and 4FR, a reason why values below the LOD are set to half the LOD before calculating the indicator of response, is to ensure that a vaccine recipient that has an unusually low antibody readout at baseline and a post-vaccination value below or near the LOD is not erroneously counted as a responder.

The following list describes the antibody variables that are measured from immunogenicity subcohort and infection case participants. (The pre-defined time points are Day 1, 35.)

1. Individual anti-Spike antibody concentration at each pre-defined time

point

2. Individual anti-Spike antibody fold-rise concentration post-vaccination relative to baseline at each pre-defined post-vaccination time point
3. 2-fold-rise and 4-fold rise (fold rise in anti-Spike antibody concentration [post/pre] $\geq 2$ and $\geq 4$, 2FR and 4FR) at each pre-defined post-vaccination time point
4. Pseudovirus-nAb responders, at each pre-defined timepoint defined as participants who had baseline values below the LLOQ with detectable pseudovirus-nAb ID50 titers above the assay LLOQ or as participants with baseline values above the LLOQ with a 4-fold increase in pseudovirus-nAb ID50 titers

Summaries of the immunogenicity data will be reported in tables. In particular, the tables will include, for each pre-defined post-baseline time point:

1. For each binding antibody marker, the estimated percentage of participants defined as responders, and with concentrations $\geq$ 2x LLOQ or $\geq$ 4 x LLOQ, will be provided with the corresponding 95% CIs using the Clopper-Pearson method.

   In addition, the estimated percentage of participants defined as responders, participants with 2-fold rise (2FR), and participants with 4-fold rise (4FR) will be provided with the corresponding 95% CIs using the Clopper-Pearson method.
2. For the ID50 pseudo-virus neutralization antibody marker, the estimated percentage of participants defined as responders, participants with 2-fold rise (2FR), and participants with 4-fold rise (4FR) will be provided with the corresponding 95% CIs using the Clopper-Pearson method
3. Geometric mean titers (GMTs) and geometric mean concentrations (GMCs) will be summarized along with their 95% CIs using the t-distribution approximation of log-transformed concentrations/titers (for each of the four Spike-targeted marker types including pseudovirus-nAb ID50 and ID80, as well as for binding Ab to N).

4. Geometric mean titer ratios (GMTRs) or geometric mean concentration ratios (GMCRs) are defined as geometric mean of individual titers/concentration ratios (post-vaccination/pre-vaccination for each injection)

5. GMTRs/GMCRs will be summarized with 95% CI (t-distribution approximation) for any post-baseline values compared to baseline, and post-Day 35 values compared to Day 35

6. The ratios of GMTs/GMCs will be estimated between groups with the two-sided 95% CIs calculated using t-distribution approximation of log-transformed titers/concentrations [the groups compared are vaccine recipient non-cases vs. vaccine recipient breakthrough cases used for Day 35 marker correlates analyses (Post Day 35 cases)].

7. The differences in the responder rates, 2FRs, 4FRs between groups will be computed along with the two-sided 95% CIs by the Wilson-Score method without continuity correction (Newcombe, 1998) (the groups for comparison are as described in the previous bullet).

All of the above point and confidence interval estimates will use inverse probability of antibody marker sampling weighting in order that estimates and inferences are for the population from which the whole study cohort was drawn. In two-phase sampling data analysis nomenclature, the "phase 1 ptids" are the per-protocol individuals excluding individuals with a COVID failure event or any other evidence of SARS-CoV-2 infection $<$ 7 days post Day 35 visit (the RT-PCR assay is used to define any evidence of SARS-CoV-2 infection). The "phase 2 ptids" are then the subset of these phase 1 ptids in the immunogenicity subcohort with Day 1 and Day 35 Ab marker data available. Thus, marker data for the COVID endpoint cases outside the subcohort will not be used in immunogenicity analyses; these cases are excluded from immunogenicity analyses.

The estimated weight $\hat{w}_{subcohort.35x}$ is the inverse sampling probability weight, calculated as the empirical fraction (No. Day 35 phase 1 ptids / No. Day 35 phase 2 ptids) within each of the baseline strata [(vaccine, placebo) $\times$ (baseline negative, baseline positive) $\times$ (demographic strata)]. For individuals outside the phase 1 ptids, $\hat{w}_{subcohort.35x}$ is assigned the missing value code NA.

All other individuals have a positive value for $\hat{w}_{subcohort.35x}$, including cases not in the subcohort. This weight is only used for case outcome-status blinded immunogenicity inferential analyses. Note that $\hat{w}_{subcohort.35x}$ is used for all immunogenicity analyses, which are based solely on the immunogenicity subcohort, for Day 1 and Day 35 markers. (Not used for correlates analyses.)

Tables will be provided separately for (1) baseline negative individuals, (2) baseline positive individuals, (3) baseline negative individuals by subgroup defined as in Table 2, and (4) baseline positive individuals by the same subgroups as in (3). Each table will show data for all available time points and for each of the vaccine and placebo arms.

Table 2: Baseline Subgroups that are Analyzed[1].

| |
|---|
| **Age:** 18-64, $\geq$ 65 |
| **Coexisting conditions:** Yes, No |
| 18-64 Coexisting conditions, 18-64 No coexisting conditions, $\geq$ 65 Coexisting conditions, $\geq$ 65 No coexisting co[illegible] |
| **Sex:** Male, Female |
| **Age x Sex:** |
| 18-64 Male, 18-64 Female, $\geq$ 65 Female, $\geq$ 65 Male |
| **Hispanic or Latino Ethnicity:** Hispanic or Latino, Not Hispanic or Latino |
| **Race or Ethnic Group:** |
| White Non-Hispanic[2], Black, Asian, American Indian or Alaska Native (NatAmer) |
| Native Hawaiian or Other Pacific Islander (PacIsl), Multiracial, |
| Other, Not reported, Unknown |
| **Underrepresented Minority Status in the U.S.:** |
| Communities of color (Comm. of color), White[2] |
| **Age x Underrepresented Minority Status in the U.S.:** |
| Age $\geq$ 65 Comm. of color, Age $<$ 65 Comm. of color, Age $\geq$ 65 White, Age $\geq$ 65 White |

[1]All analyses are done within strata defined by randomization arm and baseline positive/negative status, such that these variables are not listed here as subgroups for analysis.

[2]White Non-Hispanic is defined as Race=White and Ethnicity=Not Hispanic or Latino. All of the other Race subgroups are defined solely by the Race variable, with levels Black, Asian, American Indian or Alaska Native, Native Hawaiian or Other Pacific Islander, Multiracial, Other, Not reported, Unknown. Communities of color is defined by the complement of being known White Non-Hispanic.

For comparing antibody levels between groups, the following groups are compared:

- Baseline negative vaccine vs. baseline negative placebo
- Baseline positive vaccine vs. baseline positive placebo
- Baseline negative vaccine vs. baseline positive vaccine
- Within baseline negative vaccine recipients, compare each of the following pairs of subgroups listed in Table 2: Age $\geq 65$ vs. age $< 65$; risk for severe COVID: at risk vs. not at risk; age $\geq 65$ at risk vs. age $\geq 65$ not at risk; age $< 65$ at risk vs. age $< 65$ not at risk; male vs. female; Hispanic or Latino ethnicity: Hispanic or Latino vs. Not Hispanic or Latino; Underrepresented minority status: Communities of color vs. White Non-Hispanic (within the U.S.).

The entire immunogenicity analysis is done in the per-protocol cohort with Day 1 and Day 35 marker data available (the two-phase sample).

###### 6.1.2 Graphical description of antibody marker data

The Day 1, 35 antibody marker data collected from the immunogenicity subcohort participants will be described graphically. These data are representative of the entire study cohort. Importantly, only antibody data from the immunogenicity subcohort are included (i.e., no data from cases outside the subcohort are included). This makes the analyses unsupervised (independent of case-control status), enabling interrogation and optimization of the antibody biomarkers prior to the inferential correlates analyses.

Plots are developed for the following purposes. All of the analyses are done separately within each of the four subgroups defined by randomization arm cross-classified with baseline negative/positive status. In addition, many of the descriptive analyses will also be done separately for each demographic subgroup of interest listed above. For descriptive plots of individual marker data points that pool over one or more of the baseline strata subgroups, plots show all observed data points.

For each antibody marker readout, both Day 35 and baseline-subtracted Day 35 readouts are of interest. We will refer to the latter as 'delta.' All readouts, including delta, will be plotted on the $\log_{10}$ scale, with plotting labels on the natural scale. As such, delta is $\log_{10}$ fold-rise in the marker readout from baseline.

The following descriptive graphical analyses are done.

1. The distribution of each antibody marker readout at Day 1 and Day 35 will be described with plots of empirical reverse cumulative distribution functions (rcdfs) and boxplots (including individual data points) within each of the four groups defined by randomization arm (vaccine, placebo) and baseline positivity stratum (seronegative, seropositive). Inverse probability of sampling into the subcohort weights ($\hat{w}_{subcohort.35x}$) are used in the estimation of the rcdf curves; henceforth we refer to these weights as "inverse probability of sampling" (IPS) weights. Analyses of Day 1 markers always pool across vaccine and placebo recipients given that the two subgroups are the same at baseline.

2. Plots are arranged to compare each Day 35 marker readout between randomization arms within each of the baseline seropositive and baseline seronegative subgroups.

3. Plots are also arranged to compare each Day 35 marker readout between baseline serostatus groups within each randomization arm.

4. The correlation of each antibody marker readout among Day 1 and Day 35, and between Day 1 and fold-rise to Day 35 (delta), is examined within each randomization arm and baseline positivity stratum. Pairs plots/scatterplots will be used, annotated with baseline strata-adjusted Spearman rank correlations, implemented in the PResiduals R package available on CRAN. For calculating the correlation within each randomization arm and baseline positivity stratum, because PResiduals does not currently handle sampling weights, the correlation estimates are computed as follows: For each re-sampled data set in the second approach to graphical plotting, the covariate-adjusted Spearman correlation is calculated. The average of the estimated correlations across re-sampled data sets is reported.

5. The correlation of each pair of Day 1 antibody marker readouts are com-

pared within each baseline positivity stratum, pooling over the two randomization arms. Pairs plots/scatterplots and baseline-strata adjusted Spearman rank correlations are used, with covariate-adjusted Spearman rank correlations computed as described above. The same analyses are done for each pair of Day 35 antibody marker readouts.

6. Point estimates of Day 35 marker positive response rates for each randomization arm within each baseline positivity stratum are provided. The point and 95% CI estimates include all of the data and use IPS weights.

##### 6.2 Methods for Positive Response Calls for bAb and nAb Assays

As noted above, binding antibody responders at each pre-defined timepoint are defined as participants with concentration above the specified positivity cut-off, with a separate cut-off for each antigen Spike, RBD, N (10.8424, 14.0858, and 23.4711, respectively, in BAU/ml). This approach is used for each of the Spike and RBD and N protein antigen targets.

Pseudovirus neutralization responders at each pre-defined timepoint are defined as participants who had baseline ID50 values below the LOD with detectable ID50 neutralization titer above the assay LOD, or as participants with baseline values above the LOD with a 4-fold increase in neutralizing antibody titer. Otherwise a value is negative for pseudovirus neutralization.

##### 6.3 SARS-CoV-2 Antigen Targets Used for bAb and nAb Markers

The homologous vaccine strain antigens are used for the immune correlates analyses for the bAb markers, whereas the homologous vaccine strain with D614G mutation is used for the pseudovirus nAb markers.

#### 7 Baseline Risk Score (Proxy for SARS-CoV-2 Exposure)

We will develop baseline risk score for US only (COUNTRY==“USA”) because there are no cases in the vaccine arm and there is a single case in the placebo arm from outside of the US in the primary analysis cohort (PP-EFFFL=="Y") for the primary endpoint (PARAMCD =="PCRMMS").

Imputation of marker variables, if needed, will be done for US and Mexico together. Since we may need separate immunogenicity reports for US and Mexico, we will create two analysis-ready datasets, one for US and one for Mexico. This requires some tailoring of the correlates_processing code.

Restrictions on Treatment assignment (TRT01P=="Placebo" or "SARS-CoV-2rS"), PP status (PARAMCD =="PCRMMS1"), country and case accrual period were applied as described. The list of baseline covariates potentially relevant for SARS-CoV-2 exposure and risk of COVID was specified below (xxTable S4 in the Supplementary Material) (See the .tex file for corresponding variable names in the dataset): Age, Sex at birth (Male/Female), Race (7 categories), Ethnicity (3 categories), Height, Weight, BMI, and Protocol-defined high-risk (yes/no).

Based on these covariates, a baseline risk score is developed and controlled for in correlates analyses to adjust for potential confounding. The risk score is defined as the logit of the predicted outcome probability from a regression model estimated using the ensemble algorithm superlearner (i.e. stacking), where this logit predicted outcome is scaled to have empirical mean zero and empirical standard deviation one. The settings of superlearner (i.e., loss function, cross-validation technique, library of learners) that are used for implementation of superlearner for building a baseline risk score are described in Section 9.4.

The development of risk score will involve training the superlearner using placebo arm data and predictions made on vaccine arm data (CV-predictions will be made on placebo arm data). In both arms, risk score development will be restricted to baseline negative per-protocol subjects with cases as COVID endpoints starting post-enrollment. The CV-prediction performance of superlearner (CV-AUC calculation and CV-ROC curves) will be derived with cases as COVID endpoints starting post-enrollment as well. The prediction performance of superlearner (AUC calculation and ROC curve) in the vaccine arm, however, will be restricted to the same set of vaccine recipients as used in the correlates analyses with cases considered as COVID endpoints starting 7 days post second vaccination visit and non-cases as participants with follow-up beyond 7 days post second vaccination visit and never registered a COVID endpoint.

Independent of the superlearner risk score, important individual risk factors are also specified for inclusion as adjustment factors in correlates analyses. In particular, in addition to the risk score the at-risk indicator and the communities of color indicator are adjusted for in all correlates analyses. This choice is justified by the epidemiological data showing that these two indicators are strong infection and COVID-19 risk factors, and making use of the flexibility of super learner to develop a model for how age relates to risk.

Henceforth we refer to the baseline variables that are adjusted for in correlates analyses as "baseline factors" which, depending on the risk score results and performance, will consist of only the individual key risk factors, or key individual risk factors plus the baseline risk score.

### 8 Correlates Analysis Descriptive Tables by Case/Non-Case Status

The key table summarizing the distribution of each of the two antibody markers at the Day 1 and 35 times points is listed below. For each time point Day 1 and Day 35 separately, the positive response rate with 95% CI, and the GMT or GMC with 95% CI, is reported for each of of the case and non-case groups. In addition, the point and 95% CI estimate of the difference in positive response rate (non-cases vs. cases) and the GMT or GMC ratio (non-cases/cases), is reported.

- Immunogenicity table: Antibody levels in the baseline SARS-CoV-2 negative per-protocol cohort (vaccine recipients). Post Day 35 cases are baseline negative per-protocol vaccine recipients with the symptomatic infection COVID-19 primary endpoint diagnosed starting 7 days after the Day 35 study visit. Non-cases/Controls are baseline negative per-protocol vaccine recipients sampled into the immunogenicity subcohort with no COVID primary endpoint up to the time of data cut and no evidence of SARS-CoV-2 infection up to six days post Day 35 visit.

The point and confidence interval estimates are computed using inverse probability sampling weights $\hat{w}_{subcohort.35x}$ for Post Day 35 cases and for Non-cases, as defined in Section 9.3.1.

### 9 Correlates of Risk Analysis Plan

This analysis plan for CoRs and CoPs focuses on the COVID primary endpoint, with its continuous failure times (failure time defined by the day of the event) and no competing risks.

#### 9.1 CoR Objectives

The following CoR objectives are assessed in baseline seronegative per-protocol vaccine recipients:

1. **Univariable CoR** To assess each individual Day 35 antibody marker as a CoR of outcome in vaccine recipients, adjusting for baseline factors (See Section 7)

#### 9.2 Outline of the Set of CoR Analyses

The univariable CoR objective is addressed by Cox proportional hazards regression and nonparametric threshold regression. All of these analyses are implemented in automated and reproducible press-button fashion.

#### 9.3 Day 35 Markers Assessed as CoRs and CoPs

The following two markers at Day 35 are assessed as CoRs and CoPs, usually as quantitative variables and in some analyses as ordered trinary variables or binary variables, all of which do not subtract Day 1 (baseline) values:

1. binding Ab to Spike (IgG BAU/ml)
2. pseudovirus neutralization ID50 (IU50/ml)

For all univariable CoR analyses (first objective), the non-baseline subtracted versions of the Day 35 antibody markers are studied; the baseline-subtracted versions are not studied given that the analyses are done in the baseline negative cohort for which Day 1 readouts will generally be negative.

###### 9.3.1 Inverse probability sampling weights used in CoR analyses

In section 6.1, estimated inverse probability sampling (IPS) weights $\hat{w}_{subcohort.35x}$ were defined for per-protocol immunogenicity subcohort members, for the purpose of immunogenicity analyses. This section describes the IPS weight used for Day 35 marker correlates analyses ($\hat{w}_{35.x}$).

For baseline sampling stratum $x$ [(vaccine, placebo) $\times$ (demographic strata)], the IPS weight $w_{35.x}$ assigned to a non-case participant in stratum $x$ is defined by $\hat{w}_{35.x} = 1/\hat{\pi}_{35}(x) = N_x/n_x$, where $N_x$ is the number of stratum $x$ vaccine recipient non-cases in the Per-Protocol Baseline Negative (PPBN) cohort and $n_x$ is the number of these participants that also have Day 1, 21, and 35 marker data available, where participants with any evidence of SARS-CoV-2 infection before 7 days post Day 35 visit are excluded from the counts $N_x$ and $n_x$. For non-case participant $i$ in the immunogenicity subcohort, $\hat{w}_{35.i} = 1/\hat{\pi}_{35}(X_i)$ denotes the weight $\hat{w}_{35.x}$ for this individual's sampling stratum. All Post Day 35 cases are assigned sampling weight $N_1/n_1$ where $N_1$ is the total number of vaccine recipient cases in the PPBN cohort restricting to cases with event time starting 7 days post Day 35, and $n_1$ is the number of these participants that also had the Day 1 and 35 markers measured, and again participants with any evidence of SARS-CoV-2 infection $<$ 7 days post Day 35 visit are excluded from the counts $N_x$ and $n_x$.

In terms of two-phase sampling data analysis nomenclature, for the Day 35 marker analyses "phase 1 ptids" are defined as the entire PPBN cohort except excluding participants with any evidence of SARS-CoV-2 infection $<$ 7 days post Day 35 visit. The "phase 2 ptids" are then the subset of these phase 1 ptids with Day 1, 21, and 35 Ab marker data available. Thus the weight $\hat{w}_{35.x}$ is the inverse sampling probability weight, calculated as the empirical fraction (No. phase 1 ptids / No. phase 2 ptids) within each of the baseline negative strata (defined by PPBN vaccine group cases, PPBN placebo group cases, PPBN vaccine group non-cases divided into the demographic strata, and PPBN placebo group non-cases divided into the demographic strata).

For baseline negative individuals outside the phase 1 ptids, $\hat{w}_{35.x}$ is assigned the missing value code NA. All other individuals have a positive value for $\hat{w}_{35.x}$.

###### 9.3.2 Choice of regression methods

Time-to-event methods of Day 35 marker correlates analyses use the Day 35 visit date as the time origin.

The IPWCC Cox regression model designed for case-cohort sampling designs will be used for estimation and inference on hazard ratios of outcomes by Day 35 marker levels, and for estimation and inference on marginalized marker-conditional cumulative incidence over time. The models will be fit using the *survey* R package available on CRAN, and will adjust for the baseline factors. We use a method from the survey package that assumes without replacement two-phase sampling and not Bernoulli sampling, which matches the sampling design and approach to weight estimation (**?**).

The final time point $t_F$ of follow-up for correlates analyses is taken to be the latest COVID outcome event time. Let $T$ be the failure time, $S$ a Day 35 marker of interest, and $X$ the vector of baseline factors that are adjusted for. With $S_1(t|s,x) = P(T > t|S = s, X = x, A = 1)$, the Cox model fit yields an estimate of $S_1(t|s, X_i)$ for each individual $i$ in the phase-two sample. The marginalized conditional risk $risk_1(t|s) = E_X[P(T \leq t|s, X, A = 1)]$ through time $t$ (for all times $t$ through $t_F$ simultaneously) is estimated based on the equation

$$risk_1(t|s) \;=\; \int (1 - S_1(t|s,x))dH(x) \qquad (1)$$

where $H(\cdot)$ is the distribution of $X$ in $A = 1$ individuals.

The function $risk_1(t|s)$ can be estimated by

$$\widehat{risk}_1(t|s) = \frac{\sum_{i=1}^{n} \frac{1}{\hat{\pi}(X_i)}(1 - \hat{S}_1(t|s, X_i))}{\sum_{i=1}^{n} \frac{1}{\hat{\pi}(X_i)}}, \qquad (2)$$

where $n$ is the number of participants with phase-two data.

The bootstrap is used to obtain 95% pointwise confidence intervals for $risk_1(t_F|s)$.

The bootstrap process will be performed by resampling with replacement the subjects within the subcohort and the subjects outside the subcohort separately within each stratum and by resampling with replacement subjects with undetermined stratification variables. Across all bootstrap samples, the number of participants in each stratum in the immunogenicity subcohort remains fixed, but the number of cases does not stay the same.

The results of the above Cox modeling will be output in a variety of ways:

1. Plot $\widehat{risk}_1(t_F|s)$ vs. $s$ with 95% CIs for continuous $S = s$ varying over its whole range. Include on the plot the estimate of $\widehat{risk}_0(t_F)$ with a 95% CI for the placebo arm (horizontal bands), computed by a Cox model marginalizing over the same baseline factors as for the analysis of the vaccine arm.

2. Based on a fit of the Cox model to a nominal categorical antibody marker defined as the tertiles of $S$, plot $\widehat{risk}_1(t|s)$ for each category of $S$ values with 95% CIs, for all time points $t$ from Day 35 through $t_F$. If more than 20% of vaccine recipients have $S$ below the LOD of the assay, then the categories instead will be (1) values $\leq$ LOD; (2) values below the median of values $>$ LOD; (3) values above the median of values $>$ LOD. Include on the plot the estimated curve $\widehat{risk}_0(t)$ with 95% CIs for the placebo arm, computed by a Cox model marginalizing over the same baseline factors as for the analysis of the vaccine arm.

3. Tabular reporting of the hazard ratio per 10-fold change in the quantitative Day 35 antibody marker with 95% confidence interval and 2-sided p-value.

4. Tabular reporting of the hazard ratio for the Middle and Upper categories of the categorical Day 35 antibody marker vs. the Lower category, with 95% confidence interval and 2-sided p-value, as well as a global generalized Wald two-sided p-value for whether the hazard rate of the endpoint varies across the three categories. The table includes the attack rate (with no. of cases / no. at risk) through $t_F$ for each of the three vaccine

marker subgroups and for the placebo arm.

5. Report point and 95% CI estimates for the hazard ratio per 10-fold change in the Day 35 antibody marker, for the entire per-protocol baseline negative vaccine cohort and for each of the baseline demographic strata subgroups defined in Table 2 (reported via forest plotting).

6. Westfall-Young (1997) q-values and FWER-adjusted p-values for the generalized Wald tests are included in the table.

The bootstrap is used to calculate 95% pointwise CIs for $risk_1(t_F|s)$ in $s$. The 2-sided Wald p-value for testing the regression coefficient of the marker in the Cox model provides a valid test of the null hypothesis $H_0 : risk_1(t_F|s) = risk_1(t_F)$ for all $s$, and is reported.

In addition, the same Cox model analysis will be used to estimate the alternative marginalized conditional risk parameter defined by $risk_1(t|S \geq s)$ where $risk_1(t|S \geq s) = E_X[P(T \leq t|S \geq s, X, A = 1)]$, which can be estimated by

$$\widehat{risk}_1(t|S \geq s) = \frac{\sum_{i=1}^{n} \frac{1}{\hat{\pi}(X_i)}(1 - \hat{S}_1(t|S \geq s, X_i))}{\sum_{i=1}^{n} \frac{1}{\hat{\pi}(X_i)}}.$$

This parameter is useful because typically subgroups of interest are defined by having marker response above a threshold. We will plot $\widehat{risk}_1(t_F|S \geq s)$ vs. $s$ with 95% CIs for continuous $S$ with $s$ varying over the range of $S$ in which the number of cases to estimate $\hat{S}_1(t|S \geq s, X_i)$ is 5 or more. This type of analysis is also included because it analyzes the same parameter as the nonparametric threshold estimation method described below, providing a way to address the threshold question both by Cox modeling and by nonparametric analysis.

###### 9.3.3 Univariate CoR: Nonparametric threshold regression modeling

The targeted minimum loss-based estimation (TMLE) method of van der Laan et al. (2022) extension of the nonparametric CoR threshold estimation method of Donovan et al. (2019) is applied to each of the non-baseline subtracted antibody markers at Day 35, using the version that defines the binary outcome $Y = I(T \leq t)$ of interest as $Y = 1$ if a COVID endpoint occurred during the blinded period of follow-up and $Y = 0$ otherwise, and accounts for right-censoring times of participants. The analyses adjust for the same baseline factors $X$ as used in the Cox model CoR analyses.

The extension adjusts for baseline covariates by estimating the conditional mean function $E[Y|S \geq s, X, A = 1]$ using discrete-SuperLearner and then empirically averaging over the baseline covariates $X$ to estimate the marginal risk $risk_1^Y(S \geq s) = E_X[P(Y = 1|S \geq s, X, A = 1)]$ for each threshold $s$ of the the antibody marker in a specified discrete set. We do not perform pooled regression across the thresholds $s$, which ensures we are totally nonparametric in estimating the threshold dependence of $risk_1^Y(S \geq s)$ on $s$. The SuperLearner library includes only L1-penalized logistic regression (glmnet), because of the small number of vaccine breakthrough cases with Day 35 antibody data. An advantage of the nonparametric CoR threshold method compared to Cox modeling that specifies a log linear hazard ratio with the marker is that it can potentially detect a threshold of very low risk. The method is implemented with and without the monotonicity constraint that $risk_1^Y(S \geq s)$ is monotone non-increasing in $s$, where the results assuming monotonicity are reported unless there is evidence for violation of this assumption.

The results are reported in the same way that Donovan et al. (2019) reports results in its Figure 2, where point estimates, pointwise 95% confidence bands, and simultaneous 95% confidence bands for $risk_1^Y(S \geq s)$ are plotted for a range of threshold values. The simultaneous confidence bands cover the entire curve in $s$ with at least 95% probability and are useful for judging whether risk varies over threshold subgroups, whereas the pointwise 95% confidence bands are useful for quantifying precision at particular threshold values. The method uses the same empirical two-phase sampling estimated weights (IPS weights) as used for the other univariable IPWCC CoR analyses. In addition, for each pre-specified risk threshold $c$ set to take values over a grid with lowest value 0, the method is applied to estimate the inverse function $s_c = inf\{s : E_X[P(Y = 1|S \geq s, A = 1, X] \leq c\}$, where $s_c$ is estimated by substitution of the marginal risk function estimate. Note that the substitution estimator of $s_c$ requires that the marginal risk function is estimated for all thresholds, which is computationally infeasible. Instead, we estimate the marginal risk function on a sufficiently large discrete set and linearly interpolate to obtain marginal risk estimates for all thresholds outside the discrete set. In order for this estimand to be well defined, we operate (for this estimand only) under the assumption that $s \mapsto risk_1^Y(S \geq s)$ is monotone. For the substitution-based estimator of the inverse function $s_c$ to be well-defined, we require the estimate of $s \mapsto risk_1^Y(S \geq s)$ to be monotone as well. If there is evidence that the function estimate is not monotone then we replace the estimate with its monotone projection, which preserves its theoretical properties (Westling, van der Laan, Carone, 2020).

###### 9.3.4 P-values and Multiple hypothesis testing adjustment for CoR analysis

In general, p-values are only reported from pre-specified and automated (press-button) analyses. For the CoR analyses, p-values are reported for the univariable Cox regression analyses of the four specified Day 35 antibody marker variables. Two-sided p-values for hypothesis testing of a Day 35 marker CoR are calculated both for the Cox regression of quantitative markers (two-sided Wald tests), and for the Cox regression of markers binned into tertiles (two-sided Generalized Wald tests). Therefore a total of eight 2-sided p-values for Day 35 CoRs are calculated. However, if there are fewer than 20 vaccine breakthrough cases with Day 35 antibody data, then no p-values will be reported for the tertilized marker correlates analyses.

It is not completely clear whether to perform multiple hypothesis testing adjustment, given the expectation that the correlations among the markers are high, and possibly very high, meaning that multiplicity correction could incur a relatively high cost on the false negative error rate. However, given that robust evidence supporting an antibody marker as a CoR will be required for qualifying a marker, we will conduct multiplicity adjustment for CoR analysis, as the ability to make an inference that a marker passed pre-specified multiplicity adjusted criteria should aid an overall evidence package for establishing a validated or non-validated surrogate endpoint. Therefore, multiplicity adjustment is performed across the set of 2-sided p-values.

A permutation-based method (Westfall et al., 1993) will be used for both family-wise error rate (Holm-Bonferroni) and false-discovery rate (q-values;

Benjamini-Hochberg) correction. $10^4$ replicates of the data under the null hypotheses will be created by randomly resampling the immunologic biomarkers with replacement. For each Cox regression CoR analysis the unadjusted p-value, the FWER-adjusted p-value, and the q-value is reported for whether there is a covariate-adjusted association, where all p-values and q-values are 2-sided. The FWER-adjusted p-values and q-values are computed pooling over both the quantitative marker and tertilized marker CoR analyses. However, if there are fewer than 20 vaccine breakthrough cases with Day 35 antibody data, then no p-values are reported for the tertilized marker CoR analyses, in which case the FWER-adjusted p-values and q-values are computed pooling for the quantitative marker CoR analyses only. As a guideline for interpreting CoR findings, markers with FWER-adjusted p-value $\leq 0.05$ are flagged as having statistical evidence for being a CoR. Additionally, markers with unadjusted p-value $\leq 0.05$ and q-value $\leq 0.10$ are flagged as having a hypothesis generated for being a CoR.

##### 9.4 Implementation of superlearner for baseline risk score development

For baseline risk score development, Superlearner is applied to the placebo arm only, as mentioned in Section 7. The following details are used in the implementation of superlearner:

- Pre-scale each quantitative and ordinal variable to have empirical mean 0 and standard deviation 1.
- For the Novavax PREVENT-19 trial, there are 60 endpoint cases starting Day 1 post-enrollment. So, superlearner modeling was conducted using maximum of 20 risk score variables and 5-fold cross-validation with negative log-likelihood loss function.
- The library of adaptive and non-adaptive learners and the screens selected for superlearning are shown in Table 3. Most of the learners are non-data-adaptive type learning algorithms, such as parametric regression models (e.g., generalized linear models [glms]), which are simple, stable, and advantageous for an application with a limited number of endpoint events. Data-adaptive type algorithms are also included if the

number of endpoint events is high enough, for increasing flexibility of modeling and reducing the risk of model misspecification: SL.ranger, SL.gam, and SL.xgboost. All of the selected learners are coded into the SuperLearner R package.

- Screens used will be: 1) glmnet (lasso) pre-screening (with default tuning parameter selection), 2) logistic regression univariate 2-sided p-value screening (at level $p < 0.10$), and 3) high-correlation variable screening (described below).
- Include high-correlation variable screening, not allowing any pair of input variables to have Spearman rank correlation $r > 0.9$.
- The superlearner is conducted averaging over 10 random seeds, to make results less dependent on random number generator seed.
- No IPS weighting is needed.
- Two levels of cross-validation are used:
  - Outer level: CV-AUC computed over 5-fold cross-validation repeated 10 times to improve stability
  - Inner level: 5-fold inner CV used to estimate ensemble weights with no more than max(20, floor($n_p$/20)) input variables included in each model, where $n_p$ is the number of evaluable placebo arm cases.
- Results for comparing classification accuracy of different models are based on point and 95% confidence interval estimates of cross-validated area under the ROC curve (CV-AUC) and difference in CV-AUC as a predictiveness metric (Hubbard et al., 2016; **?**). Results are presented as forest plots of point and 95% confidence interval estimates similar to those used in Figure 3 of Neidich et al. (2019) and Magaret et al. (2019). CV-AUC is estimated using the R package *vimp* available on CRAN.

Table 3: Learning Algorithms in the Superlearner Library of Estimators of the Conditional Probability of Outcome, for Building the Baseline Risk Score Based on the Placebo Arm[1].

| Algorithms | Screens/ Tuning Parameters |
|---|---|
| SL.mean | None |
| SL.glm | Low-collinearity and (All, Lasso, LR)[2] |
| SL.glm.interaction | Low-collinearity and (Lasso, LR) |
| SL.glmnet | (alpha=1; All) |
| SL.gam | Low-collinearity and (Lasso, LR) |
| SL.xgboost[3] | All and (maxdepth,shrinkage,balance)= (4, 0.1, no) |
| SL.ranger[3] | All and balance = no |

[1]All continuous and ordinal covariates are pre-standardized to have empirical mean 0 and standard deviation 1.

[2]**All** = include all variables; **Lasso** = include variables with non-zero coefficients in the standard implementation of SL.glmnet that optimizes the lasso tuning parameter via cross-validation; **Low-collinearity** = do not allow any pairs of quantitative variables with Spearman rank correlation > 0.90; **LR** = Univariate logistic regression Wald test 2-sided p-value < 0.10.

[3]Covariate balancing (if requested) is done using option `scale_pos_weight` in SL.xgboost and option `case.weights` in SL.ranger.

Table 4: Learning Algorithms in the Superlearner Library of Estimators of the Conditional Probability of Outcome: Simplified Library in the Event of Fewer than 50 Placebo Arm Cases for an Analysis, for Building a Baseline Behavioral Risk Score in Novavax PREVENT-19[1].

| Algorithms | Screens/ Tuning Parameters |
|---|---|
| SL.mean | None |
| SL.glm | Low-collinearity and (All, Lasso, LR)[2] |
| SL.glmnet | alpha=0, 1 |
| SL.xgboost | (maxdepth,shrinkage,balance[3])= (2, 0.1, yes) (2, 0.1, no) (4, 0.1, yes) (4, 0.1, no) |
| SL.ranger | balance = (yes, no) |

[1]All continuous and ordinal covariates are pre-standardized to have empirical mean 0 and standard deviation 1.

[2]**All** = include all variables; **Lasso** = include variables with non-zero coefficients in the standard implementation of SL.glmnet that optimizes the lasso tuning parameter via cross-validation; **Low-collinearity** = do not allow any pairs of quantitative variables with Spearman rank correlation > 0.90; **LR** = Univariate logistic regression Wald test 2-sided p-value < 0.10.

[3]Covariate balancing (if requested) is done using option `scale_pos_weight` in SL.xgboost and option `case.weights` in SL.ranger.

In order to evaluate the performance of the superlearner estimated model, derived using the learning algorithms specified in Table 3, the CV-AUC is estimated with a 95% confidence interval (Hubbard et al., 2016; Williamson et al., 2022). The point and 95% confidence interval estimates of CV-AUC are reported in a forest plot, which provide a way to discern which antibody assays and readouts/markers provide the most information in predicting COVID or other outcomes. As noted above CV-AUC is estimated using the R package *vimp* available on CRAN.

If there are fewer than 50 placebo arm COVID-19 cases included in a correlates analysis, then the library of learners will be simplified to that specified in Table 4.

In addition, for selected variable sets, similar forest plots will be made comparing performance of the various estimated models (e.g., by individual learning algorithm types such as lasso), including discrete superlearner and superlearner models. The plot will be examined to determine which individual learning algorithm types are performing the best.

Cross-validated ROC curves are plotted for the superlearner estimated models for each of the input variable sets. In addition, boxplots of cross-validated estimated probabilities of outcome by case-control status (as estimated from the superlearner models) are plotted.

#### 10 Correlates of Protection: Generalities

In general, for all of the correlate of protection analyses, the same antibody markers are assessed that were analysed as correlates of risk: the Day 35 antibody markers not subtracting for the Day 1 baseline readout are used. Each of the Day 35 antibody biomarkers are separately studied as CoPs by one analysis approach summarized below.

#### 11 Correlates of Protection: Interventional Effects

In these analyses, we seek to understand whether, how, and to what extent Day 35 antibody markers impact vaccine efficacy in causal ways. We describe three approaches to this problem. Each involves consideration of a binary counterfactual outcome $Y(a, s)$ (e.g., indicator of the COVID disease endpoint by a pre-specified time) under a hypothetical intervention that both sets randomization assignment $A = a$ and sets the Day 35 immunologic marker $S$ to a fixed value or based upon a random draw from a analyst-specified distribution. Below, we assume that $S$ is scalar-valued, but some of the approaches below naturally extend to the case where a vector of immunologic markers are considered (currently such analyses are not planned). Given the central goal to develop a parsimonious surrogate endpoint based on a single immunoassay, the main analysis will use each of the methods to assess each of the four quantitative readouts (not baseline-subtracted) separately as CoPs, adjusting for the same set of baseline covariates as used in the CoR analyses previously described in Section 9.

##### 11.1 CoP: Controlled Vaccine Efficacy

We first describe the controlled vaccine efficacy curve defined as

$$\mathrm{CVE}(s) = 1 - \frac{P(Y(1, s) = 1)}{P(Y(0) = 1)} \ .$$

The value of $\mathrm{CVE}(s)$ represents the relative decrease in the probability of endpoint occurrence achieved by administering vaccine and setting Day 35 immunologic marker level to $s$ compared to the placebo control intervention, for which the Day 35 immunologic marker level is structurally set equal to the lowest possible value. Under our approach, the value of $\mathrm{CVE}(s)$ is assumed to be monotone non-decreasing in $s$; in other words, vaccine efficacy can only potentially be improved by setting greater marker levels. The extent to which the marker plays a role in determining vaccine efficacy can be determined by the degree of flatness of the graph of $\mathrm{CVE}(s)$ versus $s$.

In addition, because the primary study cohort for correlates analysis is naive to SARS-CoV-2, each of the Day 35 markers $S$ has no variability in the placebo arm [all values are 'negative,' below the positivity cut-off of the binding antibody variables and below the lower limit of detection (LOD) of the neutralizing antibody variables]. Therefore, advantageously in this setting $CVE(s)$ has a special connection to the mediation literature, where $CVE(s = LOD)$ is the natural direct effect, and vaccine efficacy is 100% mediated through $S$ if and only if $CVE(s = LOD) = 0$. Therefore inference on $CVE(s = LOD)$ evaluates full mediation.

Since $P(Y(0) = 1) = P(Y = 1 \mid A = 0)$ in view of vaccine versus placebo randomization, the controlled vaccine efficacy CVE($s$) at level $s$ can be identified using the fact that

$$P(Y(1, s) = 1) = E\left[P(Y = 1 \mid S = s, A = 1, X)\right]$$

whenever $Y(1, s)$ and $S$ are independent given $A = 1$ and a vector $X$ of covariates, and $P(S = s \mid A = 1, X) > 0$ almost surely. In other words, identification of the controlled vaccine efficacy CVE($s$) requires that a rich enough set of covariates be available so that deconfounding of the relationship between endpoint $Y$ and marker $S$ is possible in the subpopulation of vaccine recipients (no-unmeasured confounding assumption), and that marker level $S = s$ may occur within each subpopulation defined by values of the covariates $X$ (positivity assumption).

###### 11.1.1 Point and 95% confidence interval estimation of CVE($s$) and of $RR_C(s_1, s_2) = (1 - \mathbf{CVE}(s_2))/(1 - \mathbf{CVE}(s_1))$ assuming the causal assumptions hold

In this subsection, we describe how the point and 95% confidence interval estimates for CVE($s$) that are reported in the main article (**Fig. 4C**) and the Supplement are calculated, which assume that both causal assumptions mentioned above hold (no unmeasured confounders and positivity). In this subsection we also describe how the point and 95% confidence interval estimates for $RR_C(0, 1) = (1 - \text{CVE}(1))/(1 - \text{CVE}(0))$ for a binary marker $S$ are calculated, with results for $S = 1$ representing the upper tertile and $S = 0$ representing the lower tertile reported in Supplementary Text S2. In the next subsection, we describe how the sensitivity analysis is conducted, which quantifies the sensitivity of the results to potential unmeasured confounding.

Gilbert, Fong, Kenny, and Carone (2022) details the inferential and sensitivity analysis approach, which was applied to the CYD14 and CYD15 dengue phase 3 data sets (Moodie et al., 2018); the same approach was applied to the current Novavax PREVENT-19 trial data set, given that the structure of the problem is the same. We summarize here the key details needed for understanding the analysis of the PREVENT-19 trial. Under the two causal assumptions, the numerator term $P(Y(1, s) = 1)$ of $\text{CVE(s)} = 1 - P(Y(1, s) = 1)/P(Y(0) = 1)$ is

$$P(Y(1, s) = 1) = E\left[P(Y = 1 \mid S = s, A = 1, X)\right] = risk_1(t_F|s),$$

as defined in Section 9.3.2, using the notation of Section 9.3.2. That section described the Cox modeling approach that was used to compute an estimate $\widehat{risk}_1(t_F|x)$ of $risk_1(t_F|s)$, where $Y = I(T \leq t_F)$, $T$ is the time from the Day 35 marker measurement date until the COVID outcome starting 7 days post measurement date, and $t_F$ is taken to be the latest COVID outcome event time, as noted earlier.

The same estimate $\widehat{risk}_1(t_F|s)$ is used to estimate the numerator term $P(Y(1, s) = 1)$ of CVE(s). That is, there is a harmonization of the correlate of risk and controlled VE analyses, where the estimate $\widehat{risk}_1(t_F|x)$ used for the former is also used for the numerator term $P(Y(1, s) = 1)$ of CVE(s) for the latter:

$$\widehat{\text{CVE(s)}} = 1 - \frac{\widehat{risk}_1(t_F|x)}{\widehat{P}(Y(0) = 1)}$$

(where we detail the estimator $\widehat{P}(Y(0) = 1)$ next).

To estimate the denominator of CVE(s), $P(Y(0) = 1) = P(Y = 1 \mid A = 0) = P(T \leq t_F|A = 0)$, note that there is no concern about unmeasured confounding given the study is randomized. While this denominator may be estimated validly ignoring the potential baseline confounders $X$, it was estimated with adjustment for the same covariates $X$ adjusted for in the estimation of the numerator $\widehat{risk}_1(t_F|s)$. In particular, $E[P(Y = 1 \mid A = 0, X)]$ was estimated with a standard Cox model (without two-phase sampling, i.e., including all baseline negative per-protocol placebo recipients without evidence of infection by 6 days post Day 35 visit, respectively), with point estimate the average of the fitted values $\widehat{E}[P(Y_i = 1|A_i = 0, X_i)]$ across the included placebo recipients. Then, the point estimate of CVE(s) is computed as one minus the ratio of the numerator point estimate divided by the denominator point estimate. Pointwise 95% confidence intervals for $CVE(s)$ were computed using the same set of bootstrap estimates of the numerator $\widehat{risk}_1(t_F|s)$ as used for the correlates of risk analysis, and also including bootstrap estimates of the denominator $\widehat{E}[P(Y = 1 \mid A = 0, X)]$. The nonparametric percentile bootstrap method was used for the confidence intervals.

###### 11.1.2 Sensitivity analysis (to unmeasured confounding) for the Cox model controlled vaccine efficacy analysis

Sensitivity analysis is generally warranted when a no-unmeasured confounders assumption is made. The sensitivity analysis quantifies the rigor of evidence for a controlled VE CoP after accounting for potential bias from unmeasured confounding. We define $S$ to be a controlled $VE$ CoP if CVE($s$) is monotone non-decreasing in $s$ with CVE(s) < CVE(s') for at least some $s < s'$, where point and 95% confidence interval estimates of CVE($s$) versus $s$, with built in robustness to unmeasured confounding, describe the strength of the CoP in terms of the amount and nature of increase. Because the denominator $P(Y(0) = 1)$ of CVE($s$) does not depend on $s$, a controlled $VE$ CoP can equivalently be defined as the numerator $P(Y(1, s) = 1)$ being monotone non-increasing in $s$ with $P(Y(1, s) = 1) > P(Y(1, s') = 1)$ for at least some $s < s'$, where point and 95% confidence interval estimates of $P(Y(1, s) = 1)$ versus $s$ indicate some robustness to unmeasured confounding.

Two sensitivity analyses are conducted, the first of which considers the binary immunologic marker $S$ with 0 indicating the first tertile and 1 indicating the third tertile. The second sensitivity analysis considers the quantitative marker $S$ varying over its full range.

As set-up for both sensitivity analyses, for any two marker values $s_1$ and $s_2$, define the controlled risk ratio

$$RR_C(s_1, s_2) = \frac{r_C(s_2)}{r_C(s_1)} = \frac{(1 - \text{CVE}(s_2))}{(1 - \text{CVE}(s_1))},$$

where $r_C(s) = P(Y(1,s) = 1)$ is the controlled risk at $S = s$. From the observed data without the causal assumptions, the statistical parameters $r_M(s) = risk_1(t_F|s)$ (the marginalized conditional risk) and

$$RR_M(s_1, s_2) = \frac{r_M(s_2)}{r_M(s_1)}$$

(the marginalized conditional risk ratio) can be estimated. Moreover, under the causal assumptions (no-unmeasured confounding and positivity), $r_M(s) = r_C(s)$ and $RR_M(s_1, s_2) = RR_C(s_1, s_2)$. Given that CoR analysis is based on observational data — the biomarker value is not randomly assigned — a central concern is that unmeasured or uncontrolled confounding of the association between $S$ and $Y$ could render $r_M(s) \neq r_C(s)$, biasing estimates of the causal parameters of interest $r_C(s)$ and $RR_C(s_1, s_2)$. Because we can never be certain that confounding is adequately adjusted for, sensitivity analysis is warranted, as considered in extensive literature — see, e.g., VanderWeele and Ding (2017) and references therein.

Sensitivity analysis is useful to evaluate how strong unmeasured confounding would have to be to explain away an observed causal association, that is, to determine the strength of association of an unmeasured confounder between $S$ and $Y$ needed for the observed exposure-outcome association to not be causal, $r_M(s) \neq r_C(s)$ and $RR_M(s_1, s_2) \neq RR_C(s_1, s_2)$. We follow the recommendation of VanderWeele and Ding (2017) to report the E-value as a summary measure of the evidence of causality, or, in our application, evidence of whether $S$ is a controlled risk CoP based on variation in the controlled risk curve. We also include other closely related measures of sensitivity.

The E-value is the minimum strength of association, on the risk ratio scale, that an unmeasured confounder would need to have with both the exposure variable ($S$) and the outcome ($Y$) in order to fully explain away a specific observed exposure–outcome association, conditional on the measured covariates [VanderWeele and Ding (2017); VanderWeele and Mathur (2020)]. Here, in this section alone, we refer to the antibody marker $S$ as an "exposure" variable following the typical set-up in the causal inference statistical methods literature. If, as in CoP analyses, the estimated marginalized risk ratio

$\widehat{RR}_M(s_1, s_2) = \widehat{r}_M(s_2)/\widehat{r}_M(s_1)$ for $s_1 < s_2$ is less than one, then the E-value for $\widehat{RR}_M(s_1, s_2)$ is calculated as

$$e_{RR}(s_1, s_2) = \frac{1 + \sqrt{1 - \widehat{RR}_M(s_1, s_2)}}{\widehat{RR}_M(s_1, s_2)} \ . \tag{3}$$

We include the argument $(s_1, s_2)$ in the notation, with $s_1 < s_2$ by convention, to be clear that the E-value depends on specification of two specific marker-level subgroups.

To illustrate the interpretation of an E-value, suppose $S$ is binary with levels 0 and 1 and regression analysis yields an estimate $\widehat{RR}_M(0, 1) = \widehat{r}_M(1)/\widehat{r}_M(0) = 0.40$ with 95% confidence interval (CI) $(0.14, 0.78)$. An E-value $e(0, 1)$ of 4.4 means that a marginalized risk ratio $RR_M(0, 1)$ at the observed value 0.40 could be explained away (i.e., $RR_C(0, 1) = 1.0$) by an unmeasured confounder associated with both the exposure and the outcome by a marginalized risk ratio of 4.4-fold each, after accounting for the vector $X$ of measured confounders, but that weaker confounding could not do so.

In addition, we follow the recommendation of VanderWeele and Ding (2017) to also report the E-value $e_{UL}(s_1, s_2)$ for the upper limit $\widehat{UL}(s_1, s_2)$ of the 95% CI for the observed marginalized risk ratio $\widehat{RR}_M(s_1, s_2)$, computed as 1 if $\widehat{UL}(s_1, s_2) \geq 1$ and, otherwise, as

$$\frac{1 + \sqrt{1 - \widehat{UL}(s_1, s_2)}}{\widehat{UL}(s_1, s_2)} \ ,$$

which in the example equals $e_{UL}(0, 1) = 1.88$. This E-value for the upper limit indicates, for given $s_1 < s_2$, the strength of unmeasured confounding at which statistical significance of the inference that $RR_C(s_1, s_2) < 1$ would be lost. The two E-values above are useful for judging how confident we can be that an immunologic biomarker is a controlled risk CoP, with E-values near one suggesting weak support and evidence increasing with greater E-values.

Because $RR_C(s_1, s_2) = (1 - CVE(s_2))/(1 - CVE(s_1))$, evidence for $RR_C(s_1, s_2) < 1$ is equivalently evidence for $CVE(s_1) < CVE(s_2)$. Thus in a placebo-controlled trial $RR_C(s_1, s_2)$ can be interpreted as the multiplicative degree of superior vaccine efficacy caused by marker level $s_2$ vs. marker level $s_1$, and E-values quantify evidence for whether $CVE(s_1)$ is less than $CVE(s_2)$. It is also useful to provide conservative estimates of controlled risk ratios and of the controlled risk curve, accounting for unmeasured confounding. We approach these tasks based on the sensitivity analysis, or bias analysis, approach of Ding and VanderWeele (2016). We give their main result and refer readers to the paper for details.

We begin by defining two (possibly context-specific) fixed sensitivity parameters. First, we set $RR_{UD}(s_1, s_2)$ to be the maximum risk ratio for the outcome $Y$ comparing any two categories of the unmeasured confounders $U$, within either exposure group $S = s_1$ or $S = s_2$, conditional on the vector $X$ of observed covariates. Second, we set $RR_{EU}(s_1, s_2)$ to be the maximum risk ratio for any specific level of the unmeasured confounder $U$ comparing individuals with $S = s_1$ to those with $S = s_2$, with adjustment already made for the measured covariate vector $X$. Thus, $RR_{UD}(s_1, s_2)$ quantifies the importance of the unmeasured confounder $U$ for the outcome, and $RR_{EU}(s_1, s_2)$ quantifies how imbalanced the exposure/marker subgroups $S = s_1$ and $S = s_2$ are in the unmeasured confounder $U$. The values $RR_{UD}(s_1, s_2)$ and $RR_{EU}(s_1, s_2)$ are always specified as greater than or equal to one. We suppose that $RR_M(s_1, s_2) < 1$ for the fixed values $s_1 < s_2$ — this is the case of interest for immune correlates.

Define the bias factor

$$B(s_1, s_2) = \frac{RR_{UD}(s_1, s_2)RR_{EU}(s_1, s_2)}{RR_{UD}(s_1, s_2) + RR_{EU}(s_1, s_2) - 1}$$

for $s_1 \leq s_2$, and define $RR_M^U(s_1, s_2)$ the same way as $RR_M(s_1, s_2)$, except marginalizing over the joint distribution of $X$ and $U$. Then, $RR_M^U(s_1, s_2) \leq RR_M(s_1, s_2) \times B(s_1, s_2)$, where $RR_M^U(s_1, s_2) = E\{r(s_2, X^*)\}/E\{r(s_1, X^*)\}$ with $X^* = (X, U)$ and $r(s, x, u) = P(Y = 1 \mid S = s, A = 1, X = x, U = u)$ conditional risk. Translating this result to our problem context, under the positivity assumption, we have that $RR_M^U(s_1, s_2) = RR_C(s_1, s_2)$ and so, it follows that

$$RR_C(s_1, s_2) \leq RR_M(s_1, s_2) \times B(s_1, s_2)\ . \tag{4}$$

This inequality states that the controlled risk ratio is bounded above by the marginalized risk ratio multiplied by the bias factor. It follows that a conservative (upper bound) estimate of $RR_C(s_1, s_2)$ is obtained as $\widehat{RR}_M(s_1, s_2) \times B(s_1, s_2)$, and a conservative 95% CI is obtained by multiplying each confidence limit for $RR_M(s_1, s_2)$ by $B(s_1, s_2)$. These estimates for $RR_C(s_1, s_2)$ account for the presumed-maximum plausible amount of deviation from the no unmeasured confounders assumption specified by $RR_{UD}(s_1, s_2)$ and $RR_{EU}(s_1, s_2)$. An appealing feature of this approach is that the bound (4) holds without making any assumption about the confounder vector $X$ or the unmeasured confounder $U$.

*Conservative (bounded) estimation of $r_C(s)$ and $RR_C(s_1, s_2)$ for a quantitative marker $S$* The above approach does not directly provide a conservative estimate of the controlled risk curve $r_C(s)$, because additional information is needed for absolute versus relative risk estimation. To provide conservative inference for $r_C(s)$, we next select a central value $s^{cent}$ of $S$ such that $\widehat{r}_M(s^{cent})$ matches the observed overall risk, $\widehat{P}(Y = 1|A = 1)$. This value is a 'central' marker value at which the observed marginalized risk equals the observed overall risk. Next, we 'anchor' the analysis by assuming $r_C(s^{cent}) = r_M(s^{cent})$, where picking the central value $s^{cent}$ makes this plausible to be at least approximately true. Under this assumption, the bound (4) implies the bounds

$$r_C(s) \ \leq \ r_M(s)B(s^{cent}, s) \quad \text{if } s \geq s^{cent} \tag{5}$$

$$r_C(s) \ \geq \ r_M(s)\frac{1}{B(s, s^{cent})} \quad \text{if } s < s^{cent}. \tag{6}$$

Therefore, after specifying $B(s^{cent}, s)$ and $B(s^{cent}, s)$ for all $s$, we conservatively estimate $r_c(s)$ by plugging $\widehat{r}_M(s)$ into the formulas (5) and (6).

Because $B(s_1, s_2)$ is always greater than one for $s_1 < s_2$, formula (5) pulls the observed risk $\widehat{r}_M(s)$ upwards for subgroups with high biomarker values, and formula (6) pulls the observed risk $\widehat{r}_M(s)$ downwards for subgroups with low biomarker values. This makes the estimate of the controlled risk curve flatter, closer to the null curve, as desired for a sensitivity/robustness analysis.

To specify $B(s_1, s_2)$, we note that it should have greater magnitude for a greater distance of $s_1$ from $s_2$, as determined by specifying $RR_{UD}(s_1, s_2)$ and $RR_{EU}(s_1, s_2)$ increasing with $s_2 - s_1$ (for $s_1 \leq s_2$). We consider one specific approach, which sets $RR_{UD}(s_1, s_2) = RR_{EU}(s_1, s_2)$ to the common value $RR_U(s_1, s_2)$ that is specified log-linearly: $\log RR_U(s_1, s_2) = \gamma(s_2 - s_1)$ for $s_1 \leq s_2$. Then, for a user-selected pair of values $s_1 = s_1^{fix}$ and $s_2 = s_2^{fix}$ with $s_1^{fix} < s_2^{fix}$, we set a sensitivity parameter $RR_U(s_1^{fix}, s_2^{fix})$ to some value above one. It follows that

$$\log RR_U(s_1, s_2) = \left(\frac{s_2 - s_1}{s_2^{fix} - s_1^{fix}}\right) \log RR_U(s_1^{fix}, s_2^{fix}), \quad s_1 \leq s_2.$$

We anchor the analysis by setting $s_1 = s_1^{fix}$ at the $15^{th}$ percentile of the Day 35 antibody marker and $s_2 = s_2^{fix}$ at the $85^{th}$ percentile of the Day 35 antibody marker.

Once $r_C(s)$ is conservatively estimated via the formulas (5) and (6), it is immediate how to obtain a conservative estimate of CVE$(s)$:

$$\widehat{\text{CVE}}(s) = 1 - \frac{\widehat{r}_M(s)B(s^{cent}, s)}{\widehat{P}(Y(0) = 1)},$$

where the estimate of the placebo arm risk, $\widehat{P}(Y(0) = 1)$, is the same as for the controlled VE analysis assuming no-unmeasured confounders.

*Sensitivity analyses for controlled vaccine efficacy reported in the article*

The sensitivity analysis is done for each of the two Cox model CoR analyses described in Section 9.3.2, first for the binary Day 35 marker and second for the quantitative Day 35 marker. For the former analysis, E-values are reported for both the point estimate and the upper 95% confidence limit for $RR_C(0, 1)$, where category 1 is the upper tertile (vaccine recipients with antibodies $S$ in the top third), category 0 is the lower tertile (vaccine recipients with antibodies in the bottom third), and the intermediate middle tertile subgroup of vaccine recipients is excluded from the analysis. In addition, we set $RR_{UD}(0,1) = RR_{EU}(0,1) = 2$, such that $B(0,1) = 4/3$, and report conservative estimation and inference on the controlled risk ratio $RR_C(0,1)$ and equivalently on the ratio of controlled vaccine efficacy curves $RR_C(0,1) = (1 - CVE(1))/(1 - CVE(0))$.

Next, we conduct the sensitivity analysis treating $S$ as a quantitative variable, as detailed in the subsection above "*Conservative (bounded) estimation of $r_C(s)$ and $RR_C(s_1, s_2)$ for a quantitative marker $S$*." This analysis reports results in terms of point and 95% point-wise confidence interval estimates of CVE($s$) vs. $s$ assuming the specified amount of unmeasured confounding that makes the estimates of CVE($s$) flatter than under the assumption of no unmeasured confounding.

For validity the controlled risk/vaccine efficacy analyses require the positivity assumption, and thus the methods will only be applied if the data are reasonably supportive of the positivity assumption. To check positivity, we study the antibody marker distribution in vaccine recipients within each subgroup of the covariates $X$ that are adjusted for. For the tertiles analysis we require evidence that within each subgroup some vaccine recipients have lower tertile responses and some vaccine recipients have upper tertile responses. For the quantitative $S$ analysis, we look for evidence that $S$ varies over its full range within each level of the potential confounders that are adjusted for.

The details of the above causal sensitivity analysis are described in the in-press article (Gilbert et al., 2022).

#### 12 Estimating a Threshold of Protection Based on an Established or Putative CoP (Population-Based CoP)

For each antibody marker studied as a CoP, we will apply the Chang-Kohberger (2003) / Siber (2007) method to estimate a threshold of the antibody marker associated with the estimate of overall vaccine efficacy observed in the trial.

This method makes two simplifying assumptions: (1) that a high enough antibody marker value $s^*$ implies that individuals with $S > s^*$ have essentially zero disease risk (perfect protection) regardless of whether they were vaccinated; and (2) $P(Y = 1|S \leq s^*, A = 1)/P(Y = 1|S \leq s^*, A = 0) = 1$ (zero vaccine efficacy if $S \leq s^*$). Based on these assumptions, $s^*$ is calculated as the value equating $1 - \widehat{P}(S \leq s^*|A = 1)/\widehat{P}(S \leq s^*|A = 0)$ to the estimate of overall vaccine efficacy. This estimate is supplemented by estimating the reverse cumulative distribution function (RCDF) of $S$ in baseline negative vaccine recipients and calculating a 95% confidence interval for the threshold value $s^*$ as the points of intersection of the estimated RCDF curve with the 95% confidence interval for overall vaccine efficacy (as in the figure in Andrews and Goldblatt, 2014).

This method essentially assumes that $S$ has already been established as a CoP, and under that assumption estimates a threshold that may be considered as a benchmark / study endpoint for future immunogenicity vaccine trial applications.

It is acknowledged that this approach makes simplifying assumptions that are diagnosed to be violated in the PREVENT-19 trial; nonetheless it may yield a useful benchmark and complementary information on a threshold correlate of protection.

#### 13 Considerations for Baseline SARS-CoV-2 Positive Study Participants

As stated above, if enough COVID cases in baseline positive vaccine and/or placebo recipients occur, then additional correlates analyses may be planned in baseline positive individuals. For example, the same or similar correlates of risk analysis plan that is used to analyze Day 35 marker correlates of risk in baseline negative vaccine recipients could be applied to assess Day 1 marker correlates of risk in baseline positive placebo recipients. In addition, analyses could be done to assess how vaccine efficacy in baseline positive participants varies with Day 1 markers. It is straightforward to make this analysis rigorous because Day 1 markers are a baseline covariate, such that regression analyses are valid based on the randomization.

#### 14 Avoiding Bias with Pseudovirus Neutralization Analysis due to Use of Anti-HIV Antiretroviral Drugs

Because the lentivirus-based pseudovirus neutralization assay uses an HIV backbone, the presence of anti-retroviral drugs in serum will give a false positive neutralization signal. This can be easily screened for using an MuLV pseudotype control. Therefore, Day 1 and Day 35 samples of all study participants with data included in correlates analyses will be tested for presence of anti-retroviral drugs. Samples with the control indicating likely anti-retroviral are excluded from analyses, for all analyses that include pseudovirus neutralization. Analyses that do not consider pseudovirus neutralization are unaffected by this issue.

A

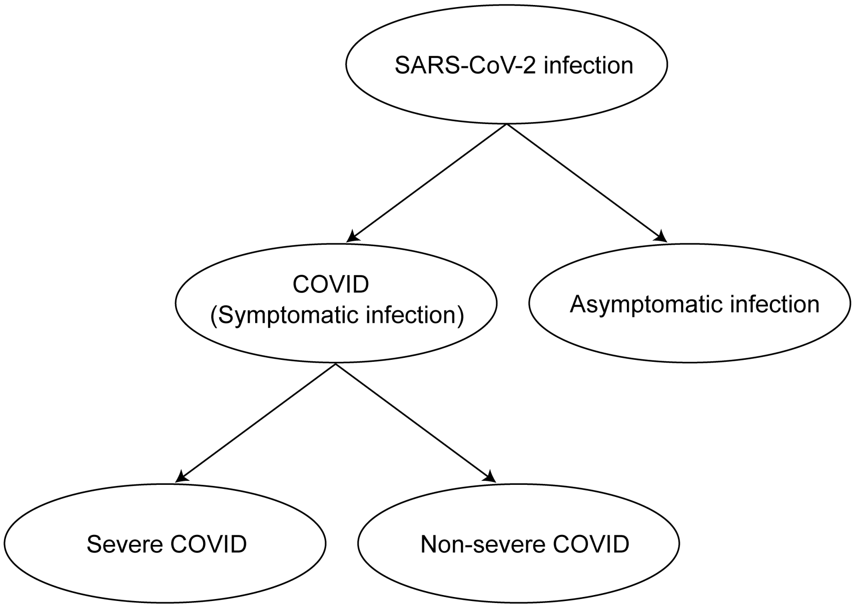

B

| **Clinical Endpoint** | **Definition** |
|---|---|
| SARS-CoV-2 infection | Positive RNA PCR test or SARS-CoV-2 seroconversion*, whichever occurs first |
| COVID (Symptomatic infection) | Meeting a protocol-specified list of COVID-19 symptoms with virological confirmation of SARS-CoV-2 infection (symptom triggered) |
| Asymptomatic infection | SARS-CoV-2 seroconversion* without prior diagnosis of the COVID endpoint¶ |
| Severe COVID | COVID endpoint with at least one protocol-specified severe disease event |
| Non-severe COVID | COVID endpoint with zero protocol-specified severe disease events |

*Seroconversion is assessed via a validated assay that distinguishes natural vs vaccine-induced SARS-CoV-2 antibodies
¶ Alternatively, the asymptomatic infection endpoint can also include an RNA PCR+ test result obtained through testing regardless of symptoms (e.g., as a requirement for travel, return to school or work, or elective medical procedures) and follow-up to confirm the participant remains asymptomatic

Figure 2: A) Structural relationships among study endpoints in a COVID-19 vaccine efficacy trial (Mehrotra et al., 2020). B) Study endpoint definitions.

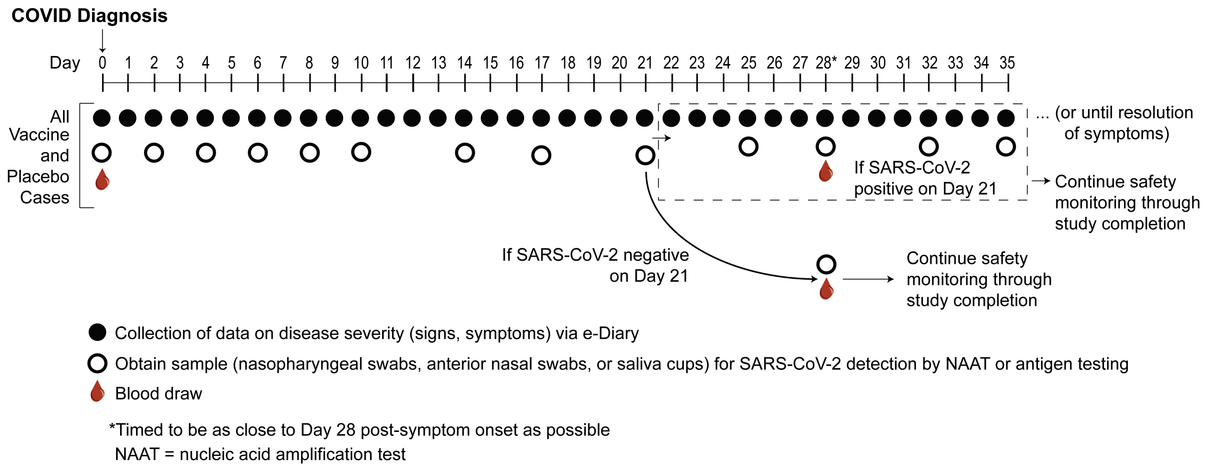

Figure 3: Example at-COVID diagnosis and post-COVID diagnosis disease severity and virologic sampling schedule, in a setting where frequent follow-up of confirmed cases can be assured. Participants diagnosed with virologically-confirmed symptomatic SARS-CoV-2 infection (COVID) enter a post-diagnosis sampling schedule to monitor viral load and COVID-related symptoms (types, severity levels, and durations).

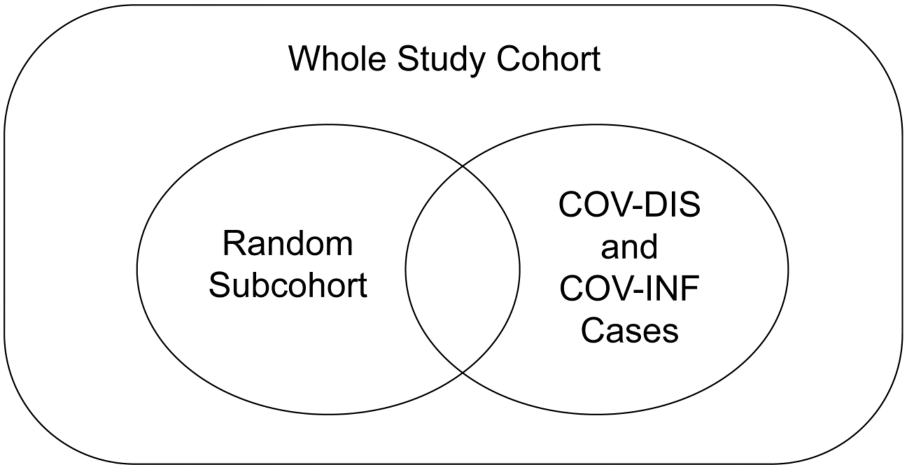

Figure 4: Case-cohort sampling design (Prentice, 1986) that measures Day 1, Day 35 antibody markers in all participants selected into the subcohort and in all COVID and COV-INF cases occurring outside of the subcohort.

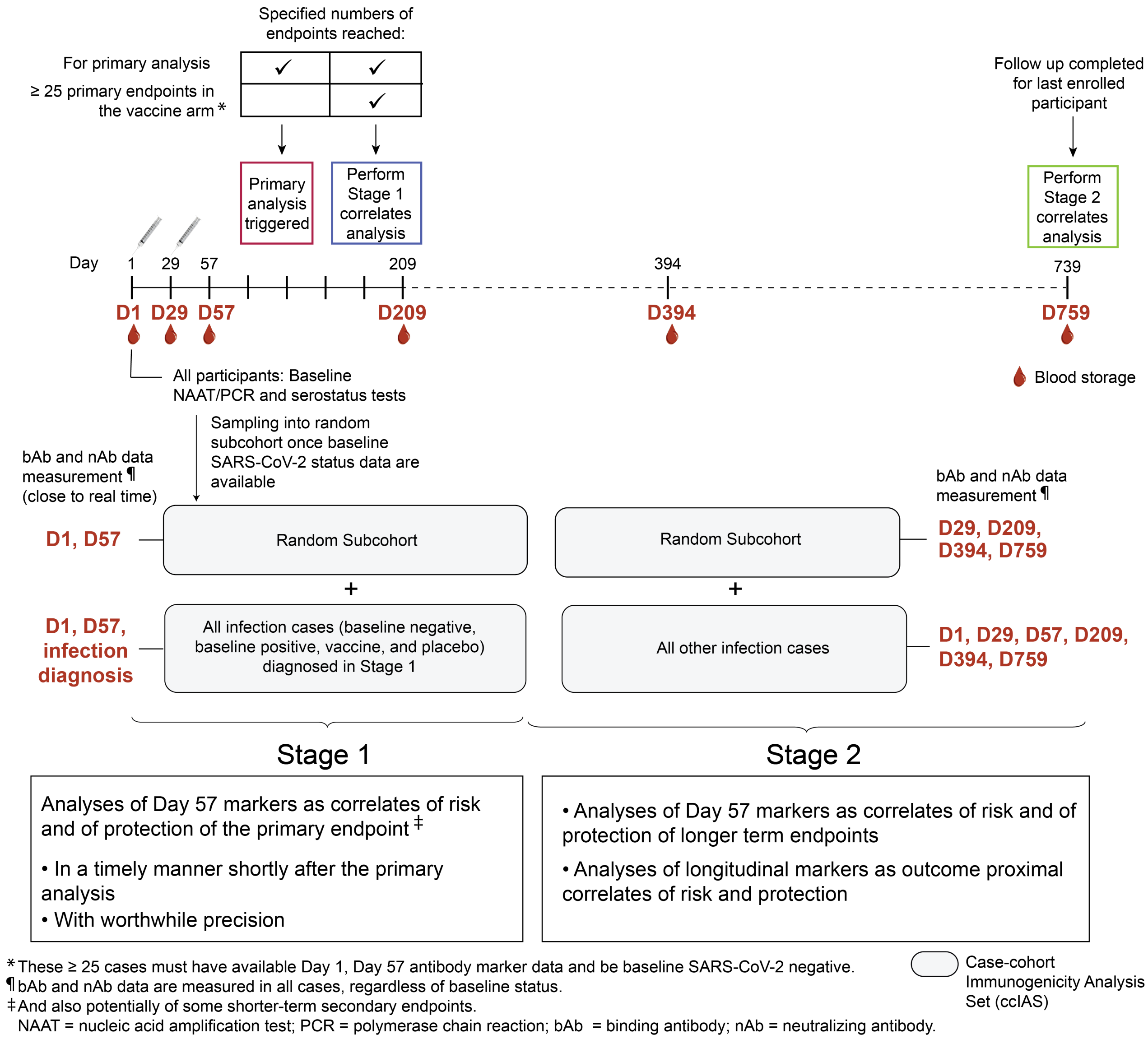

Figure 5: Two-stage correlates analysis. Stage 1 consists of analyses of Day 35 markers as correlates of risk and of protection of the primary endpoint and potentially also of some secondary endpoints, and includes antibody marker data from all COVID and SARS-CoV-2 infection cases (COV-INF) through to the time of the data lock for the first correlates analyses. Stage 2 consists of analyses of Day 35 markers as correlates of risk and of protection of longer term endpoints and analyses of longitudinal markers as outcome-proximal correlates of risk and of protection, and includes antibody marker data from all subsequent COVID and COV-INF cases. Stage 1 measures Day 1, Day 35 antibody markers and COV-INF and COVID diagnosis time point markers; Stage 2 measures antibody markers from all sampling time points and COV-INF plus COVID diagnosis sampling time points not yet assayed. The same immunogenicity subcohort is used for both stages.

#### 15 Novavax Binary Principal Stratification Results

**list of variables to include in Baseline behavior risk**

Table 5: Novavax PREVENT-19: Correlates of Vaccine Efficacy Results by Gilbert et al. (2020) Method for High vs. Low Marker Subgroups Under No Early Harm Assumption with Sensitivity Analysis Scenarios*

| | Marker | Sens | Ilol | Ilou | Elol | Elou | Ihil | Ihiu | Ehil | Ehiu | Icnl | Icnu | Ecnl | Ecnu |
|---|---|---|---|---|---|---|---|---|---|---|---|---|---|---|
| 1 | D57S.4 | None | 0.79 | 0.79 | 0.64 | 0.88 | 0.95 | 0.95 | 0.92 | 0.97 | 4.22 | 4.22 | 2.17 | 8.21 |
| 2 | D57S.4 | Med | 0.72 | 0.84 | 0.56 | 0.90 | 0.95 | 0.95 | 0.92 | 0.97 | 3.05 | 5.80 | 1.73 | 10.05 |
| 3 | D57S.4 | High | 0.58 | 0.89 | 0.34 | 0.94 | 0.94 | 0.95 | 0.92 | 0.97 | 1.89 | 8.99 | 1.04 | 15.49 |
| 4 | D57RBD.4 | None | 0.77 | 0.77 | 0.60 | 0.86 | 0.95 | 0.95 | 0.93 | 0.97 | 4.68 | 4.68 | 2.41 | 9.11 |
| 5 | D57RBD.4 | Med | 0.69 | 0.83 | 0.52 | 0.89 | 0.95 | 0.95 | 0.93 | 0.97 | 3.38 | 6.42 | 1.91 | 11.14 |
| 6 | D57RBD.4 | High | 0.53 | 0.89 | 0.28 | 0.93 | 0.95 | 0.95 | 0.92 | 0.97 | 2.09 | 9.97 | 1.15 | 17.18 |
| 7 | D57ID50.4 | None | 0.75 | 0.75 | 0.58 | 0.84 | 0.88 | 0.88 | 0.80 | 0.93 | 2.12 | 2.12 | 1.06 | 4.24 |
| 8 | D57ID50.4 | Med | 0.66 | 0.81 | 0.48 | 0.88 | 0.87 | 0.89 | 0.80 | 0.93 | 1.46 | 3.03 | 0.81 | 5.40 |
| 9 | D57ID50.4 | High | 0.48 | 0.88 | 0.22 | 0.92 | 0.85 | 0.90 | 0.77 | 0.93 | 0.83 | 4.97 | 0.45 | 9.13 |
| 10 | D57ID80.4 | None | 0.73 | 0.73 | 0.55 | 0.84 | 0.89 | 0.89 | 0.81 | 0.93 | 2.35 | 2.35 | 1.16 | 4.74 |
| 11 | D57ID80.4 | Med | 0.64 | 0.80 | 0.45 | 0.87 | 0.88 | 0.89 | 0.81 | 0.93 | 1.62 | 3.36 | 0.89 | 6.04 |
| 12 | D57ID80.4 | High | 0.45 | 0.87 | 0.16 | 0.92 | 0.86 | 0.90 | 0.78 | 0.94 | 0.93 | 5.51 | 0.49 | 10.24 |
| 13 | D57S.5 | None | 0.87 | 0.87 | 0.79 | 0.92 | 0.95 | 0.95 | 0.93 | 0.97 | 2.87 | 2.87 | 1.52 | 5.42 |
| 14 | D57S.5 | Med | 0.83 | 0.90 | 0.75 | 0.93 | 0.95 | 0.96 | 0.92 | 0.97 | 2.07 | 3.93 | 1.21 | 6.68 |
| 15 | D57S.5 | High | 0.76 | 0.93 | 0.65 | 0.95 | 0.94 | 0.96 | 0.91 | 0.97 | 1.28 | 6.08 | 0.74 | 10.31 |
| 16 | D57RBD.5 | None | 0.87 | 0.87 | 0.80 | 0.92 | 0.95 | 0.95 | 0.92 | 0.97 | 2.65 | 2.65 | 1.40 | 5.00 |
| 17 | D57RBD.5 | Med | 0.84 | 0.90 | 0.76 | 0.93 | 0.95 | 0.96 | 0.92 | 0.97 | 1.91 | 3.63 | 1.12 | 6.17 |
| 18 | D57RBD.5 | High | 0.77 | 0.93 | 0.66 | 0.95 | 0.94 | 0.96 | 0.91 | 0.98 | 1.18 | 5.72 | 0.68 | 10.51 |
| 19 | D57ID50.5 | None | 0.75 | 0.75 | 0.62 | 0.83 | 0.91 | 0.91 | 0.82 | 0.95 | 2.72 | 2.72 | 1.26 | 5.84 |
| 20 | D57ID50.5 | Med | 0.67 | 0.81 | 0.54 | 0.86 | 0.90 | 0.92 | 0.82 | 0.95 | 1.87 | 3.88 | 0.98 | 7.36 |
| 21 | D57ID50.5 | High | 0.53 | 0.87 | 0.33 | 0.91 | 0.87 | 0.93 | 0.78 | 0.96 | 1.06 | 6.34 | 0.54 | 12.19 |
| 22 | D57ID80.5 | None | 0.78 | 0.78 | 0.65 | 0.86 | 0.88 | 0.88 | 0.80 | 0.93 | 1.90 | 1.90 | 0.93 | 3.87 |
| 23 | D57ID80.5 | Med | 0.72 | 0.83 | 0.58 | 0.89 | 0.87 | 0.90 | 0.79 | 0.94 | 1.31 | 2.72 | 0.72 | 4.93 |
| 24 | D57ID80.5 | High | 0.59 | 0.88 | 0.40 | 0.92 | 0.84 | 0.91 | 0.74 | 0.94 | 0.74 | 4.43 | 0.39 | 8.16 |
| 25 | D57S.6 | None | 0.88 | 0.88 | 0.82 | 0.92 | 0.97 | 0.97 | 0.94 | 0.98 | 3.82 | 3.82 | 1.87 | 7.80 |
| 26 | D57S.6 | Med | 0.85 | 0.90 | 0.80 | 0.93 | 0.96 | 0.97 | 0.94 | 0.98 | 2.76 | 5.24 | 1.51 | 9.51 |
| 27 | D57S.6 | High | 0.80 | 0.92 | 0.72 | 0.95 | 0.96 | 0.97 | 0.93 | 0.98 | 1.70 | 8.07 | 0.93 | 14.62 |
| 28 | D57RBD.6 | None | 0.88 | 0.88 | 0.83 | 0.92 | 0.97 | 0.97 | 0.94 | 0.98 | 3.42 | 3.42 | 1.71 | 6.84 |
| 29 | D57RBD.6 | Med | 0.85 | 0.90 | 0.80 | 0.93 | 0.96 | 0.97 | 0.93 | 0.98 | 2.47 | 4.68 | 1.37 | 8.36 |
| 30 | D57RBD.6 | High | 0.80 | 0.93 | 0.73 | 0.95 | 0.95 | 0.97 | 0.92 | 0.98 | 1.52 | 7.22 | 0.84 | 12.86 |
| 31 | D57ID50.6 | None | 0.78 | 0.78 | 0.67 | 0.85 | 0.92 | 0.92 | 0.81 | 0.96 | 2.65 | 2.65 | 1.08 | 6.47 |
| 32 | D57ID50.6 | Med | 0.72 | 0.82 | 0.62 | 0.87 | 0.90 | 0.93 | 0.80 | 0.96 | 1.82 | 3.78 | 0.86 | 8.00 |
| 33 | D57ID50.6 | High | 0.62 | 0.87 | 0.47 | 0.91 | 0.87 | 0.94 | 0.75 | 0.97 | 1.03 | 6.17 | 0.47 | 13.30 |
| 34 | D57ID80.6 | None | 0.79 | 0.79 | 0.69 | 0.86 | 0.90 | 0.90 | 0.80 | 0.95 | 2.07 | 2.07 | 0.91 | 4.69 |
| 35 | D57ID80.6 | Med | 0.74 | 0.84 | 0.64 | 0.88 | 0.88 | 0.91 | 0.78 | 0.95 | 1.42 | 2.95 | 0.71 | 5.86 |
| 36 | D57ID80.6 | High | 0.64 | 0.88 | 0.50 | 0.92 | 0.85 | 0.93 | 0.72 | 0.96 | 0.81 | 4.82 | 0.39 | 9.77 |

*None: beta sensitivity parameters log(1.0) and -log(1.0); Med: beta sensitivity parameters log(0.75) and -log(0.75); High: beta sensitivity parameters log(0.5) and -log(0.5)

Table 6: Novavax PREVENT-19: Correlates of Vaccine Efficacy Results by Gilbert et al. (2020) Method for High vs. Low Marker Subgroups Under No Early Harm Assumption with Sensitivity Analysis Scenarios*

| | Marker | Sens | Ilol | Ilou | Elol | Elou | Ihil | Ihiu | Ehil | Ehiu | Icnl | Icnu | Ecnl | Ecnu |
|---|---|---|---|---|---|---|---|---|---|---|---|---|---|---|
| 1 | D29S.3 | None | 0.78 | 0.78 | 0.54 | 0.89 | 0.94 | 0.94 | 0.92 | 0.96 | 3.62 | 3.62 | 1.63 | 8.04 |
| 2 | D29S.3 | Med | 0.70 | 0.84 | 0.45 | 0.92 | 0.94 | 0.94 | 0.92 | 0.96 | 2.61 | 4.97 | 1.30 | 9.62 |
| 3 | D29S.3 | High | 0.54 | 0.90 | 0.17 | 0.95 | 0.94 | 0.94 | 0.91 | 0.96 | 1.62 | 7.72 | 0.77 | 14.75 |
| 4 | D29RBD.3 | None | 0.85 | 0.85 | 0.65 | 0.93 | 0.94 | 0.94 | 0.91 | 0.95 | 2.36 | 2.36 | 0.97 | 5.72 |
| 5 | D29RBD.3 | Med | 0.79 | 0.89 | 0.59 | 0.95 | 0.93 | 0.94 | 0.91 | 0.95 | 1.70 | 3.24 | 0.79 | 6.78 |
| 6 | D29RBD.3 | High | 0.69 | 0.93 | 0.38 | 0.97 | 0.93 | 0.94 | 0.91 | 0.95 | 1.05 | 5.04 | 0.47 | 10.40 |
| 7 | D29ID50.3 | None | 0.58 | 0.58 | 0.34 | 0.73 | 0.90 | 0.90 | 0.84 | 0.94 | 4.23 | 4.23 | 2.25 | 7.95 |
| 8 | D29ID50.3 | Med | 0.43 | 0.69 | 0.17 | 0.79 | 0.89 | 0.91 | 0.84 | 0.94 | 2.92 | 6.05 | 1.71 | 10.24 |
| 9 | D29ID50.3 | High | 0.13 | 0.80 | -0.27 | 0.87 | 0.88 | 0.91 | 0.83 | 0.94 | 1.67 | 9.82 | 0.97 | 16.58 |
| 10 | D29S.4 | None | 0.86 | 0.86 | 0.76 | 0.91 | 0.95 | 0.95 | 0.92 | 0.96 | 2.60 | 2.60 | 1.40 | 4.83 |
| 11 | D29S.4 | Med | 0.81 | 0.89 | 0.71 | 0.93 | 0.94 | 0.95 | 0.92 | 0.96 | 1.88 | 3.58 | 1.11 | 5.97 |
| 12 | D29S.4 | High | 0.72 | 0.93 | 0.57 | 0.95 | 0.94 | 0.95 | 0.91 | 0.96 | 1.16 | 5.54 | 0.67 | 9.23 |
| 13 | D29RBD.4 | None | 0.87 | 0.87 | 0.78 | 0.92 | 0.94 | 0.94 | 0.92 | 0.96 | 2.29 | 2.29 | 1.23 | 4.29 |
| 14 | D29RBD.4 | Med | 0.83 | 0.90 | 0.73 | 0.94 | 0.94 | 0.95 | 0.92 | 0.96 | 1.65 | 3.15 | 0.97 | 5.31 |
| 15 | D29RBD.4 | High | 0.74 | 0.93 | 0.61 | 0.96 | 0.93 | 0.95 | 0.91 | 0.96 | 1.02 | 4.89 | 0.59 | 8.20 |
| 16 | D29ID50.4 | None | 0.66 | 0.66 | 0.49 | 0.77 | 0.91 | 0.91 | 0.85 | 0.95 | 3.88 | 3.88 | 2.01 | 7.47 |
| 17 | D29ID50.4 | Med | 0.55 | 0.74 | 0.37 | 0.82 | 0.90 | 0.92 | 0.85 | 0.95 | 2.67 | 5.54 | 1.53 | 9.60 |
| 18 | D29ID50.4 | High | 0.33 | 0.83 | 0.07 | 0.88 | 0.89 | 0.93 | 0.82 | 0.95 | 1.53 | 8.98 | 0.87 | 15.53 |
| 19 | D29S.5 | None | 0.87 | 0.87 | 0.81 | 0.92 | 0.95 | 0.95 | 0.93 | 0.97 | 2.72 | 2.72 | 1.52 | 4.86 |
| 20 | D29S.5 | Med | 0.84 | 0.90 | 0.77 | 0.93 | 0.95 | 0.96 | 0.93 | 0.97 | 1.96 | 3.74 | 1.20 | 6.06 |
| 21 | D29S.5 | High | 0.77 | 0.93 | 0.68 | 0.95 | 0.94 | 0.96 | 0.92 | 0.97 | 1.21 | 5.78 | 0.73 | 9.36 |
| 22 | D29RBD.5 | None | 0.88 | 0.88 | 0.81 | 0.92 | 0.95 | 0.95 | 0.93 | 0.97 | 2.68 | 2.68 | 1.50 | 4.78 |
| 23 | D29RBD.5 | Med | 0.84 | 0.90 | 0.78 | 0.93 | 0.95 | 0.96 | 0.93 | 0.97 | 1.93 | 3.67 | 1.18 | 5.96 |
| 24 | D29RBD.5 | High | 0.77 | 0.93 | 0.68 | 0.95 | 0.94 | 0.96 | 0.92 | 0.97 | 1.19 | 5.68 | 0.72 | 9.20 |
| 25 | D29ID50.5 | None | 0.75 | 0.75 | 0.63 | 0.83 | 0.90 | 0.90 | 0.83 | 0.94 | 2.62 | 2.62 | 1.33 | 5.17 |
| 26 | D29ID50.5 | Med | 0.68 | 0.80 | 0.55 | 0.86 | 0.89 | 0.91 | 0.82 | 0.95 | 1.81 | 3.75 | 1.02 | 6.62 |
| 27 | D29ID50.5 | High | 0.54 | 0.86 | 0.36 | 0.90 | 0.86 | 0.92 | 0.78 | 0.96 | 1.03 | 6.11 | 0.58 | 11.56 |
| 28 | D29S.6 | None | 0.89 | 0.89 | 0.84 | 0.92 | 0.97 | 0.97 | 0.94 | 0.98 | 3.29 | 3.29 | 1.74 | 6.23 |
| 29 | D29S.6 | Med | 0.86 | 0.91 | 0.81 | 0.93 | 0.96 | 0.97 | 0.94 | 0.98 | 2.37 | 4.52 | 1.39 | 7.70 |
| 30 | D29S.6 | High | 0.81 | 0.93 | 0.74 | 0.95 | 0.95 | 0.97 | 0.92 | 0.98 | 1.46 | 6.97 | 0.84 | 11.87 |
| 31 | D29RBD.6 | None | 0.89 | 0.89 | 0.84 | 0.92 | 0.96 | 0.96 | 0.94 | 0.98 | 3.01 | 3.01 | 1.61 | 5.62 |
| 32 | D29RBD.6 | Med | 0.86 | 0.91 | 0.82 | 0.93 | 0.96 | 0.97 | 0.93 | 0.98 | 2.17 | 4.13 | 1.28 | 6.96 |
| 33 | D29RBD.6 | High | 0.81 | 0.93 | 0.75 | 0.95 | 0.95 | 0.97 | 0.92 | 0.98 | 1.33 | 6.37 | 0.78 | 10.73 |

*None: beta sensitivity parameters log(1.0) and -log(1.0); Med: beta sensitivity parameters log(0.75) and -log(0.75); High: beta sensitivity parameters log(0.5) and -log(0.5)

Table 7: Novavax PREVENT-19: Cut-points Defining High and Low Marker Subgroups

| | Perc. 0.4 | Perc. 0.5 | Perc. 0.6 |
|---|---|---|---|
| D57Spike | 1380.38 | 2094.11 | 2851.67 |
| D57RBD | 1895.40 | 3087.45 | 4218.91 |
| D57ID50 | 157.98 | 223.46 | 297.17 |
| D57ID80 | 381.24 | 497.62 | 606.18 |

Table 8: Novavax PREVENT-19: Cut-points Defining High and Low Marker Subgroups

| | Perc. 0.3 | Perc. 0.4 | Perc. 0.5 |
|---|---|---|---|
| D29Spike | 55.89 | 140.02 | 225.48 |
| D29RBD | 52.35 | 139.38 | 225.79 |
| D29ID50 | 5.71 | 8.67 | 13.76 |
| D29ID80 | 16.43 | 21.77 | 27.56 |

Table 9: Novavax PREVENT-19: Cut-points Defining High and Low Marker Subgroups

| | Perc. 0.3 | Perc. 0.4 | Perc. 0.5 |
|---|---|---|---|
| D29Spike | 1.75 | 2.15 | 2.35 |
| D29RBD | 1.72 | 2.14 | 2.35 |
| D29ID50 | 0.76 | 0.94 | 1.14 |
| D29ID80 | 1.22 | 1.34 | 1.44 |
